# Methodological Adherence, Reporting Completeness, and Risk of Bias in AI-Assisted Systematic Reviews: A Matched Meta-Research Study

**DOI:** 10.64898/2026.11.06.26367388

**Authors:** Mohammad Jay, Maryam Esttaifo, Ali Samani, Emma Wilson, Sarah Esttaifo, Mary Morgan, Joanne Abi-Jaoude, Grace Reszetnik, Meera Chopra, Abdulrahman Alrashidi, Jonathan Karl Monteiro, Geoffrey Schemitsch, Vishwathsen Karthikeyan, Ogechi Obi, Fiona Emdin, David-Dan Nguyen, Naharin Sultana Anni, Rinku Sutradhar, Christoffer Dharma, Catherine Yu, Sharon E. Straus, Lorraine L. Lipscombe, Antoine Eskander

## Abstract

**Background:** Artificial intelligence (AI) is increasingly used to accelerate systematic reviews that inform clinical practice and policy. However, evidence on the methodological adherence, reporting completeness, and risk of bias of completed AI-assisted reviews is limited.

**Objective:** To compare methodological adherence, reporting completeness, and risk of bias between published systematic reviews with reported AI assistance and matched reviews with no reported AI use, and to assess AI-use transparency.

**Methods:** We conducted a matched-cohort meta-research study of English-language, peer-reviewed health systematic reviews. AI-assisted reviews published during 2023–2025 were matched 1:2 to reviews with no reported AI use using individualized MEDLINE searches targeting publication timing, clinical domain, review type, and meta-analysis status. When more than two eligible comparators were available, two were randomly selected. AI assistance was defined as reported use during searching, screening, data extraction, risk-of-bias assessment, evidence synthesis, or substantive drafting. Full texts and supplementary materials were examined to classify reported AI use. The primary outcome was a prespecified modified 13-item AMSTAR-2 methodological-adherence score (range 0–13; noninferiority margin −1 point). Secondary outcomes included PRISMA 2020 reporting completeness and ROBIS risk of bias; AI-use transparency was assessed descriptively. Adapted 13-item AMSTAR-2 confidence categories were summarized descriptively, while critical- and noncritical-domain adherence was examined in exploratory adjusted analyses. Multivariable regression models adjusted for journal impact tier, funding reporting, formal review-group affiliation, review-team size, and review-question type, using cluster-robust standard errors by matched set. Sensitivity analyses included matched-set fixed-effects models. The study was registered on the Open Science Framework (DOI: 10.17605/OSF.IO/Q5JRW).

**Results:** The cohort included 231 systematic reviews: 77 AI-assisted and 154 with no reported AI use. Mean 13-item AMSTAR-2 scores were 7.58 and 9.63, respectively. In multivariable-adjusted analyses, AI-assisted reviews scored 1.64 points lower (95% CI −2.37 to −0.91); noninferiority was not demonstrated. PRISMA adherence was estimated to be 4.7 percentage points lower among AI-assisted reviews in primary adjusted analyses (95% CI −8.9 to −0.6) and 6.4 points lower in matched-set fixed-effects analyses (95% CI −10.7 to −2.1). Among reviews with assessable ROBIS ratings, 75.0% of AI-assisted reviews and 65.6% of comparators were rated low risk (adjusted difference +17.8 percentage points; 95% CI 2.7 to 33.0). Among AI-assisted reviews, model version or date was reported in 42.9%, prompts or parameters in 55.8%, and data-governance or ethics considerations in 9.1%.

**Conclusions:** Reviews reporting AI assistance had lower methodological-adherence scores and had less complete reporting, despite more favorable risk-of-bias ratings. These findings provide an early warning that AI-assisted systematic reviews may require stronger safeguards, human verification, and transparency to support trust.

## Introduction

Systematic reviews inform clinical guidelines, policy, reimbursement, and practice; their credibility depends on prespecified, transparent, and reproducible methods.[1] Their resource demands create strong incentives to use artificial intelligence (AI) to accelerate evidence synthesis.[2]

Early platforms such as Rayyan introduced semi-automated support for title-and-abstract screening, while newer automated and generative tools now support search development, citation screening and prioritization, data extraction, risk-of-bias assessment, synthesis, and manuscript drafting.[3–8] Task-level evaluations have demonstrated potential efficiency gains but variable accuracy, including missed studies during screening and errors in data extraction and evidence appraisal.[9–15] However, these evaluations do not establish whether completed AI-assisted reviews preserve methodological adherence, reporting completeness, and transparency across the full review workflow. The limited studies evaluating completed or AI-generated reviews have generally been conducted in narrow clinical or methodological settings.[16,17] These task-level evaluations provide important evidence about the benefits and limitations of AI-assisted methods, but leave a complementary question unanswered: whether completed reviews reporting AI assistance differ in methodological adherence, reporting completeness, and risk of bias, irrespective of whether any observed differences are attributable to AI use itself.

Established instruments assess different aspects of review trustworthiness: AMSTAR-2 appraises methodological weaknesses, PRISMA 2020 promotes transparent reporting of systematic reviews, and ROBIS assesses risk of bias in review processes.[18–21] However, these frameworks were developed for conventional workflows and do not explicitly require AI-use details such as tool identity and version, prompts or parameters, human verification, or data-governance safeguards.[18–21] Emerging guidance, including PRISMA-trAIce, GAMER, and TRIPOD-LLM, is beginning to define expectations for AI disclosure, but empirical evidence on the methodological adherence, standard reporting completeness, and AI-use transparency of completed AI-assisted reviews remains limited.[22–24] As AI-assisted systematic reviews become more common, clinicians, guideline developers, and policymakers need empirical evidence on whether these reviews adhere to established methods and report AI-mediated steps sufficiently for appraisal.

We therefore compared methodological adherence, reporting completeness, and risk of bias between published systematic reviews with reported AI assistance and matched reviews with no reported AI use. Among AI-assisted reviews, we also assessed AI-specific transparency using the Artificial Intelligence Transparency and Disclosure Index (AITDI), a preliminary meta-research instrument specified in the published protocol.[25]

## Methods

### Study Design

We conducted a matched-cohort meta-research study comparing peer-reviewed systematic reviews with reported AI assistance with systematic reviews with no reported AI use. The study was preregistered on the Open Science Framework (DOI: 10.17605/OSF.IO/Q5JRW), and its protocol was published in JMIR Research Protocols.[25] Detailed methods are provided in Supplementary Appendix.

### Information Sources and Search Strategy

An information specialist searched MEDLINE (Ovid), CINAHL (EBSCOhost), and the Cochrane Database of Systematic Reviews on December 4–5, 2025. The Ovid MEDLINE search strategy was peer reviewed by a second information specialist using the PRESS 2015 Checklist.[26] Complete strategies and retrieval counts are provided in Section S2 and Appendix 2 of Supplementary Appendix.

### Eligibility Criteria and Exposure Classification

Eligible articles were English-language, peer-reviewed systematic reviews, with or without meta-analysis, addressing human health topics. To maintain comparability, we excluded protocols, conference abstracts, commentaries, editorials, narrative reviews, scoping reviews, rapid reviews, evidence maps, umbrella reviews, and reviews focused exclusively on preclinical or animal research. Title-and-abstract and full-text screening were performed independently by two reviewers in Covidence (disagreements resolved through discussion or third-reviewer adjudication).

Reviews published from 2023 through 2025, an early period of widespread large language model adoption, were classified as AI-assisted when the article or supplement explicitly reported AI use during a substantive review stage, including search or retrieval, screening or study selection, data extraction, risk-of-bias assessment, synthesis or analysis, or substantive drafting or interpretation.[25] Use of conventional review-management software (e.g., Covidence, Rayyan, or RevMan) alone was not classified as AI assistance unless authors explicitly reported using an AI-enabled feature for a substantive review task. AI use limited to language editing or readability support was not considered substantive review conduct. Comparators met the same criteria but had no reported AI use; this does not establish AI nonuse (Supplementary Methods S3–S4).

### Comparator Identification and Selection

After each AI-assisted review was confirmed eligible, an information specialist constructed an individualized Ovid MEDLINE search for candidate comparators, targeting publication within 3 months and similarity in broad clinical domain, review type, and meta-analysis status. These characteristics guided comparator identification rather than requiring exact matching. Reviews were matched 1:2, a ratio selected to improve precision while preserving feasibility.[25,27] When more than two potentially eligible comparators were available, two were selected by reproducible random sampling in R (seed 42). Selected comparators underwent independent duplicate full-text assessment; ineligible records were replaced using the next randomized record (Supplementary Methods S5). Detailed procedures and exceptions are described in Supplementary Methods S5.

### Data Extraction and Appraisal

Review characteristics and AMSTAR-2, PRISMA 2020, ROBIS, and AI-transparency assessments were extracted independently by two reviewers using piloted forms. Discrepancies were resolved by discussion or third-reviewer adjudication, and the completed dataset underwent consistency checks (Supplementary Methods S6).

### Outcomes

The primary outcome was methodological adherence, measured using a prespecified modified 13-item AMSTAR-2 score applicable to reviews with or without meta-analysis (range, 0–13; higher scores indicate greater adherence).[18,25] Although AMSTAR-2 was not designed as a formal additive quality score for individual reviews, we used the modified score to compare the average number of criteria fully satisfied across groups rather than to assign overall methodological quality to individual reviews. Because individual criteria differ in methodological importance, we also examined critical- and noncritical-domain adherence and adapted 13-item AMSTAR-2 confidence categories. Only a full “Yes” counted as adherent. The noninferiority margin of −1.0 point was selected a priori as the largest acceptable average decrement in methodological adherence, approximately equivalent to one fewer criterion satisfied on the 13-item scale.[25] This pragmatic threshold was not based on a validated minimally important difference.

Secondary outcomes were the full 16-item AMSTAR-2 score among reviews with meta-analysis; adapted 13-item AMSTAR-2 confidence categories; PRISMA 2020 adherence (range, 0 to 1, with partial reporting receiving half credit); and ROBIS overall risk of bias.[18,19,21] For comparative ROBIS analyses, low risk was coded as favorable and high or unclear risk as not low risk.

Among AI-assisted reviews, AI-specific transparency was assessed using the AITDI, a preliminary, not-yet-validated meta-research instrument comprising six scored items representing distinct transparency domains (total score, 0 to 6; higher scores indicate more complete reporting) and one additional nonscored item addressing AI-related limitations.[25] AITDI was used descriptively and was not intended as a formal reporting guideline or measure of review quality. Exploratory outcomes included AMSTAR-2 critical- and noncritical-domain scores, AMSTAR-2 and PRISMA item-level adherence, PRISMA domain-level adherence, ROBIS domain-level ratings, receipt-to-publication interval, 12-month citation counts, and Altmetric Attention Score (Supplementary Methods S7).

### Covariates

Adjusted models included journal impact tier, funding reporting, formal review-group affiliation, review-team size operationalized as number of authors, and review-question type. These covariates were selected based on their potential associations with reported AI use and review methodological quality, reflecting differences in review resources, reporting infrastructure, and methodological context. Covariates were not screened for statistical significance before inclusion in the adjusted models. No studies were excluded from the primary adjusted analysis because of missing covariate data. Unavailable categorical information was retained using explicitly defined categories rather than excluding observations. No covariate imputation was performed (Supplementary Methods S8).

### Statistical Analysis

The primary estimand was the adjusted mean difference in the 13-item AMSTAR-2 score, calculated as AI-assisted reviews minus reviews with no reported AI use. Ordinary least-squares regression used CR2 cluster-robust variance estimation by matched set with Satterthwaite corrections.[28] Noninferiority required the lower limit of the two-sided 95% confidence interval to exceed −1.0 point.

The AMSTAR-2 16-item score and PRISMA adherence were analyzed using linear regression; ROBIS outcomes were analyzed with linear probability models. Adapted 13-item AMSTAR-2 confidence categories and AITDI item and total scores were summarized descriptively. Receipt-to-publication interval was modeled as an absolute difference in days; 12-month citation counts and Altmetric Attention Scores were modeled after log(1 + outcome) transformation, with estimates expressed as relative percentage differences. Analyses used available observations; receipt-to-publication, citation, and Altmetric models adjusted for journal tier and publication year.

Sensitivity analyses included paired matched-set comparisons, restriction to standard systematic reviews, matched-set fixed-effects models, leave-one-journal-out analyses, and overlap weighting.[29] Exploratory analyses included AMSTAR-2, PRISMA, and ROBIS domain- and item-level models; review-group-stratified models of factors associated with AMSTAR-2 and PRISMA scores; and associations of AITDI scores with AMSTAR-2 and PRISMA among AI-assisted reviews. Analyses were performed in R. ChatGPT (OpenAI) was used for limited troubleshooting, minor revisions, and documentation of the R analysis script. AI-assisted changes were reviewed and verified by the authors. Study-level data and appraisal scores are provided in Multimedia Appendix 2, the R analysis script in Multimedia Appendix 3, and protocol deviations in Section S12 of Supplementary Appendix. The complete study-level dataset and R analysis script were provided to the journal for peer review but are not publicly available with this preprint.

### Patient and Public Involvement

Patients and members of the public were not directly involved in this meta-research study. The study was designed and conducted by a multidisciplinary team of clinicians, researchers, and evidence-synthesis experts with experience producing, appraising, and using systematic reviews.

### Ethics approval

Research ethics board approval and informed consent were not required because this meta-research study evaluated publicly available published systematic reviews and did not involve human participants or identifiable patient-level data.

## Results

### Study Sample

The AI-assisted and comparator searches yielded 5967 and 267 screened records, respectively. After full-text assessment and matching, the final analytic cohort comprised 231 reviews in 77 matched sets, including 77 AI-assisted reviews and 154 comparator reviews with no reported AI use (Figure 1; Appendix 3 of Supplementary Appendix).

**Figure 1.**
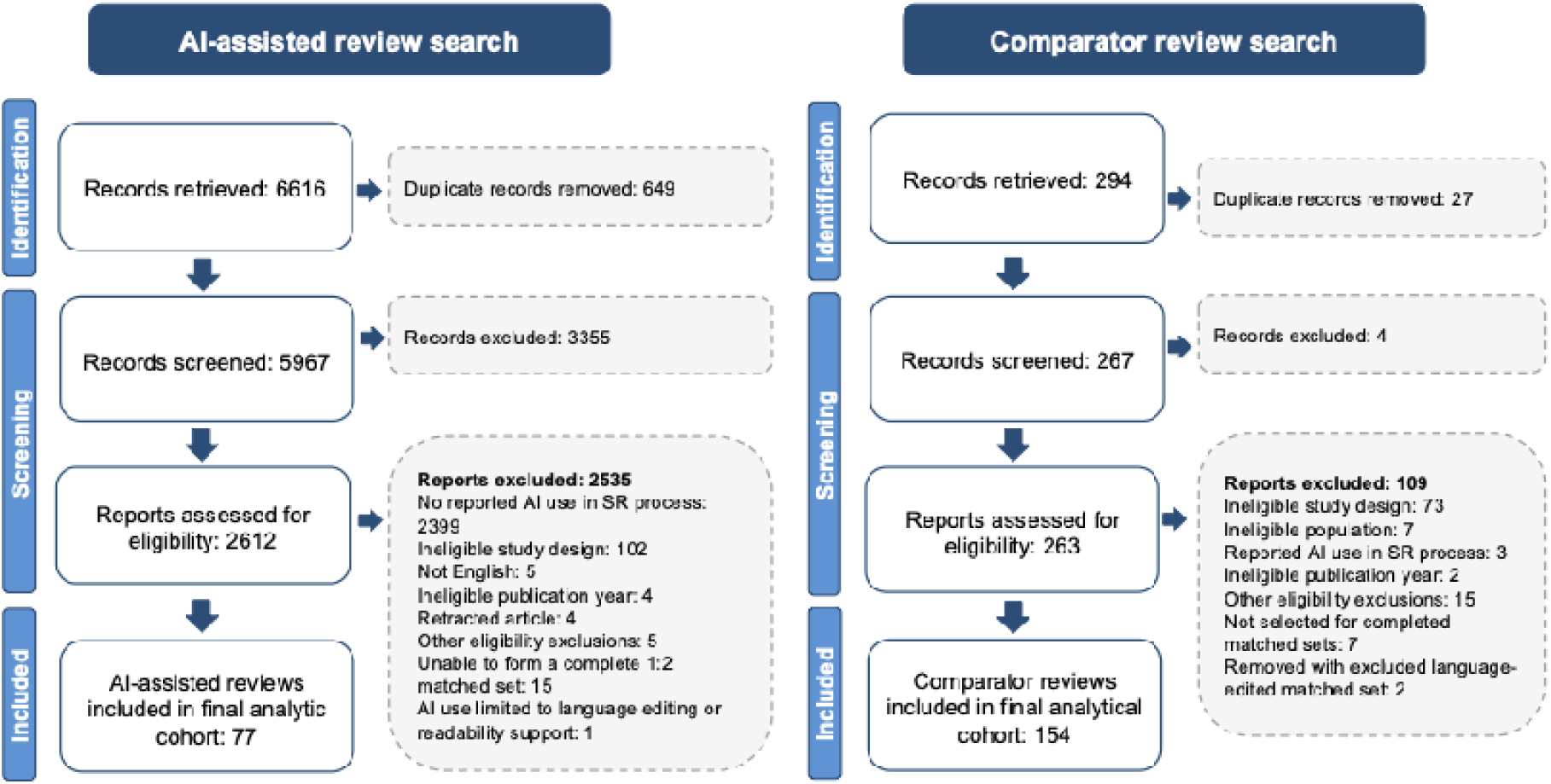
Study Selection Flow Diagram. AI denotes artificial intelligence; SR, systematic review. Comparator reviews were reviews with no reported AI use.

Most reviews were published in 2024 or 2025, appeared in journals in the highest impact quartile, and included a funding disclosure statement. Both groups searched a median of 4 databases (IQR, 3–5) and had a median of 6 authors. AI-assisted reviews less often included meta-analysis (40.3% vs. 55.8%), less often addressed intervention questions (19.5% vs. 44.2%), and more often addressed methodological questions (26.0% vs. 4.5%) than reviews with no reported AI use (Table 1; Table S1). Study-level characteristics and appraisal outcomes are provided in Tables S2–S3.

**Table 1.**
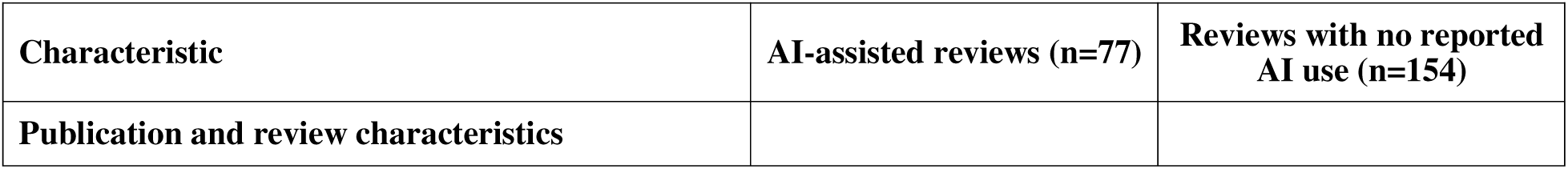
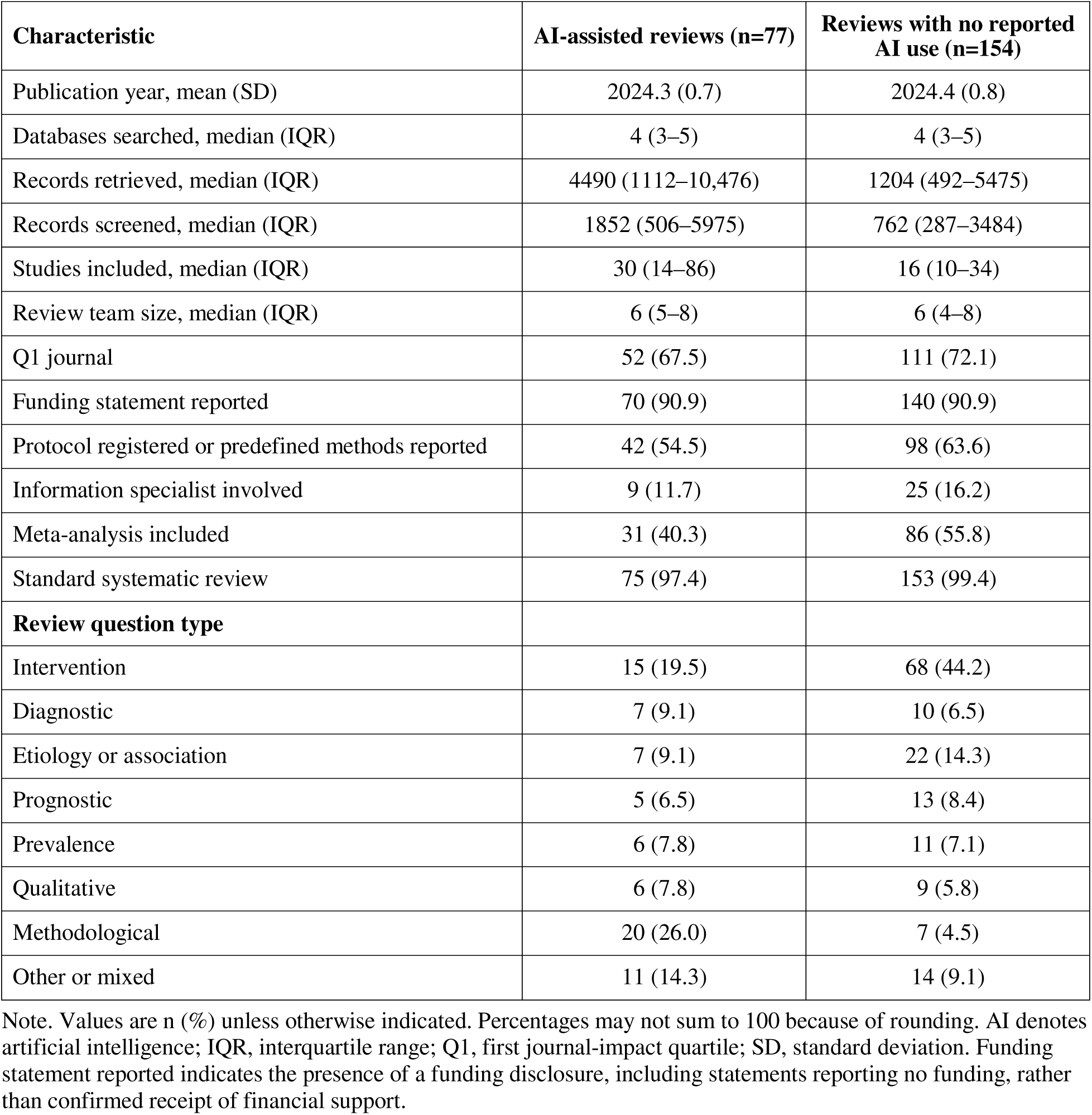
Selected Characteristics of AI-Assisted Reviews and Reviews with No Reported AI Use.

| **Characteristic** | **AI-assisted reviews (n=77)** | **Reviews with no reported AI use (n=154)** |
|---|---|---|
| **Publication and review characteristics** |  |  |
| **Characteristic** | **AI-assisted reviews (n=77)** | **Reviews with no reported AI use (n=154)** |
| Publication year, mean (SD) | 2024.3 (0.7) | 2024.4 (0.8) |
| Databases searched, median (IQR) | 4 (3–5) | 4 (3–5) |
| Records retrieved, median (IQR) | 4490 (1112–10,476) | 1204 (492–5475) |
| Records screened, median (IQR) | 1852 (506–5975) | 762 (287–3484) |
| Studies included, median (IQR) | 30 (14–86) | 16 (10–34) |
| Review team size, median (IQR) | 6 (5–8) | 6 (4–8) |
| Q1 journal | 52 (67.5) | 111 (72.1) |
| Funding statement reported | 70 (90.9) | 140 (90.9) |
| Protocol registered or predefined methods reported | 42 (54.5) | 98 (63.6) |
| Information specialist involved | 9 (11.7) | 25 (16.2) |
| Meta-analysis included | 31 (40.3) | 86 (55.8) |
| Standard systematic review | 75 (97.4) | 153 (99.4) |
| **Review question type** |  |  |
| Intervention | 15 (19.5) | 68 (44.2) |
| Diagnostic | 7 (9.1) | 10 (6.5) |
| Etiology or association | 7 (9.1) | 22 (14.3) |
| Prognostic | 5 (6.5) | 13 (8.4) |
| Prevalence | 6 (7.8) | 11 (7.1) |
| Qualitative | 6 (7.8) | 9 (5.8) |
| Methodological | 20 (26.0) | 7 (4.5) |
| Other or mixed | 11 (14.3) | 14 (9.1) |

Note. Values are n (%) unless otherwise indicated. Percentages may not sum to 100 because of rounding. AI denotes artificial intelligence; IQR, interquartile range; Q1, first journal-impact quartile; SD, standard deviation. Funding statement reported indicates the presence of a funding disclosure, including statements reporting no funding, rather than confirmed receipt of financial support.

### Methodological Adherence

Mean 13-item AMSTAR-2 scores were 7.58 for AI-assisted reviews and 9.63 for reviews with no reported AI use. The adjusted difference was −1.64 points (95% CI, −2.37 to −0.91; P<0.001), so noninferiority was not demonstrated. The matched-set fixed-effects estimate was −1.73 points (95% CI, −2.42 to −1.04). Among reviews with meta-analysis, the adjusted 16-item difference was −2.71 points (95% CI, −3.76 to −1.66) (Table 2; Figure 2; Figures S1-S2). Adapted 13-item confidence categories were high, moderate, low, and critically low in 2.6%, 0.0%, 27.3%, 70.1% of AI-assisted reviews, respectively; corresponding proportions among reviews with no reported AI use were 14.3%, 4.5%, 32.5%, 48.7% (Table S4).

**Figure 2.**
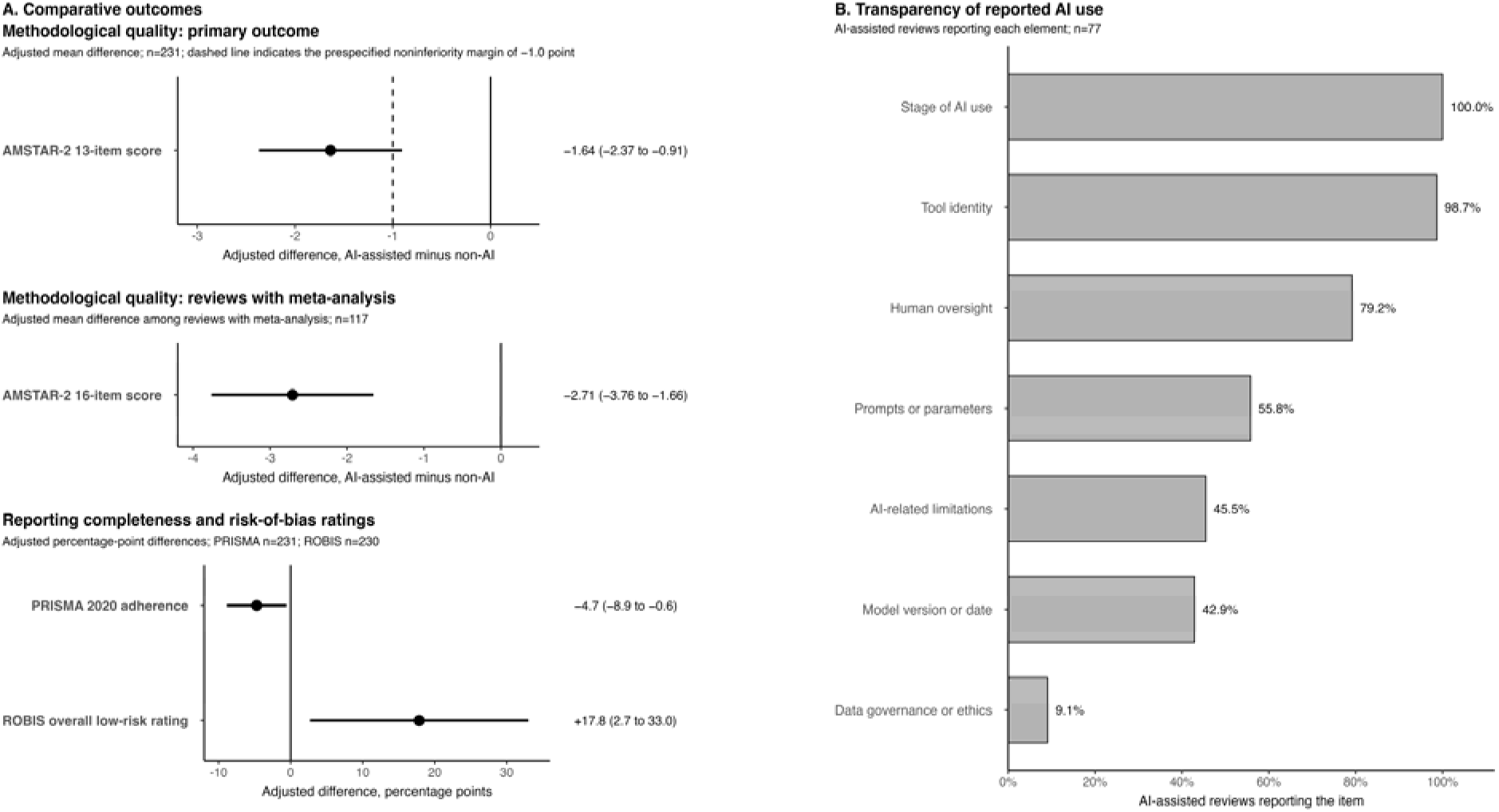
Comparative Outcomes and Transparency of Reported AI Use. Panel A shows adjusted between-group differences with 95% confidence intervals; negative values favor reviews with no reported AI use for AMSTAR-2 and PRISMA outcomes, whereas positive values favor AI-assisted reviews for the ROBIS low-risk outcome. The dashed vertical line indicates the prespecified noninferiority margin of −1.0 point for the primary AMSTAR-2 outcome. Adjusted models included journal impact tier, funding reporting, review-group affiliation, review-team size, and review-question type, with standard errors clustered by matched set. Panel B shows the proportion of AI-assisted reviews reporting each transparency element. AI denotes artificial intelligence; AMSTAR-2, A Measurement Tool to Assess Systematic Reviews, version 2; PRISMA, Preferred Reporting Items for Systematic Reviews and Meta-Analyses; ROBIS, Risk of Bias in Systematic Reviews.

**Table 2.**
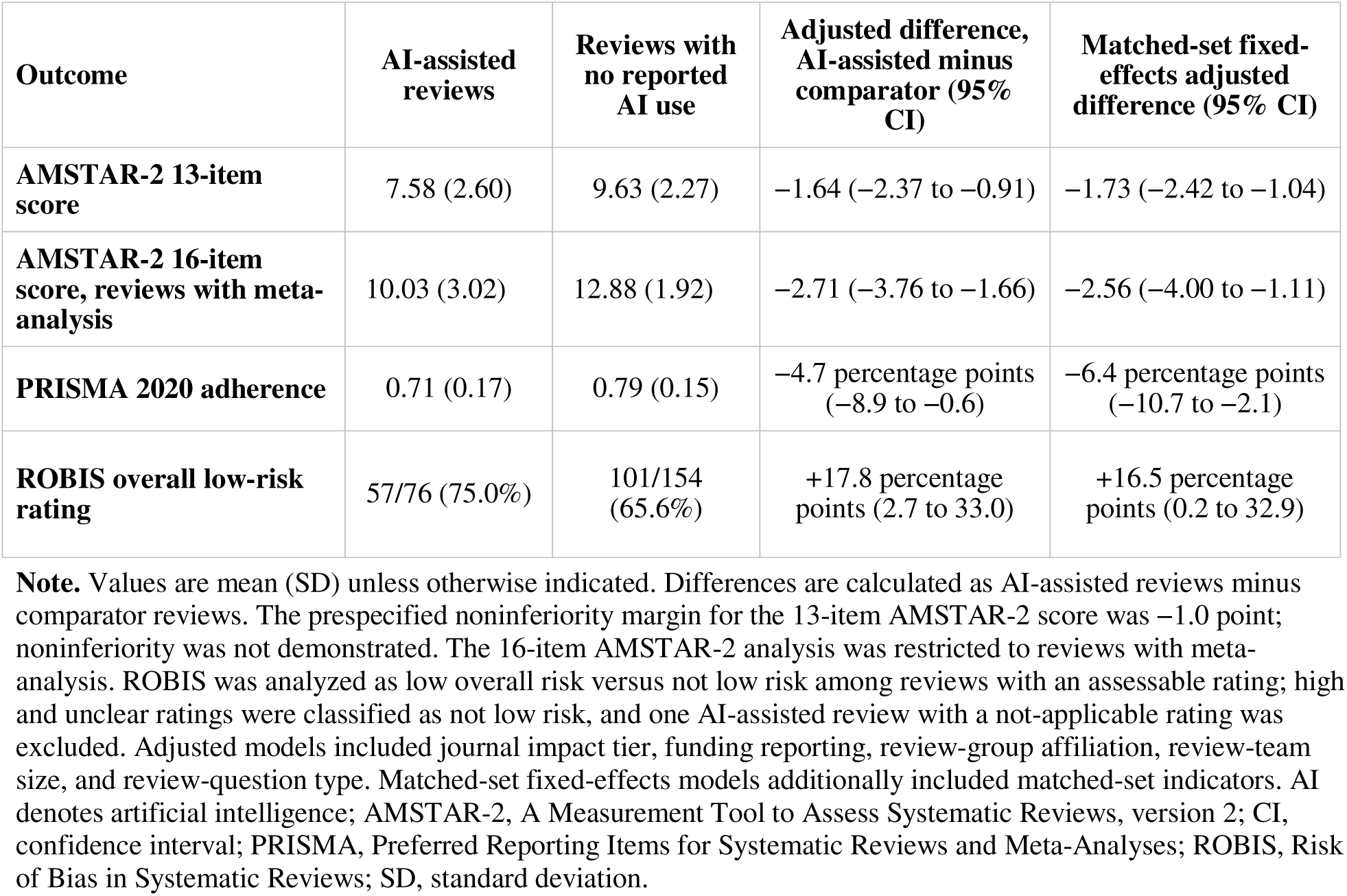
Primary and Secondary Outcomes Comparing AI-Assisted Reviews with Reviews with No Reported AI Use.

| **Outcome** | **AI-assisted reviews** | **Reviews with no reported AI use** | **Adjusted difference, AI-assisted minus comparator (95% CI)** | **Matched-set fixed-effects adjusted difference (95% CI)** |
|---|---|---|---|---|
| **AMSTAR-2 13-item score** | 7.58 (2.60) | 9.63 (2.27) | −1.64 (−2.37 to −0.91) | −1.73 (−2.42 to −1.04) |
| **AMSTAR-2 16-item score, reviews with meta-analysis** | 10.03 (3.02) | 12.88 (1.92) | −2.71 (−3.76 to −1.66) | −2.56 (−4.00 to −1.11) |
| **PRISMA 2020 adherence** | 0.71 (0.17) | 0.79 (0.15) | −4.7 percentage points (−8.9 to −0.6) | −6.4 percentage points (−10.7 to −2.1) |
| **ROBIS overall low-risk rating** | 57/76 (75.0%) | 101/154 (65.6%) | +17.8 percentage points (2.7 to 33.0) | +16.5 percentage points (0.2 to 32.9) |

**Note.** Values are mean (SD) unless otherwise indicated. Differences are calculated as AI-assisted reviews minus comparator reviews. The prespecified noninferiority margin for the 13-item AMSTAR-2 score was −1.0 point; noninferiority was not demonstrated. The 16-item AMSTAR-2 analysis was restricted to reviews with meta-analysis. ROBIS was analyzed as low overall risk versus not low risk among reviews with an assessable rating; high and unclear ratings were classified as not low risk, and one AI-assisted review with a not-applicable rating was excluded. Adjusted models included journal impact tier, funding reporting, review-group affiliation, review-team size, and review-question type. Matched-set fixed-effects models additionally included matched-set indicators. AI denotes artificial intelligence; AMSTAR-2, A Measurement Tool to Assess Systematic Reviews, version 2; CI, confidence interval; PRISMA, Preferred Reporting Items for Systematic Reviews and Meta-Analyses; ROBIS, Risk of Bias in Systematic Reviews; SD, standard deviation.

In adjusted analyses, AI-assisted reviews had lower critical-domain scores (−0.58 points; 95% CI, −0.96 to −0.19) and noncritical-domain scores (−1.06 points; 95% CI, −1.53 to −0.58), with consistent descriptive and meta-analysis-restricted findings (Tables S5-S7). The largest item-level differences were for defining review-question components (−25.7 percentage points; 95% CI, −39.2 to −12.2), justifying eligible study designs (−24.2; 95% CI, −37.7 to −10.7), and explaining heterogeneity (−21.9; 95% CI, −36.2 to −7.6). Among critical items, AI-assisted reviews were less likely to consider risk of bias when interpreting results (−16.3 percentage points; 95% CI, −31.7 to −0.9) or provide an excluded-studies list with justifications (−16.1; 95% CI, −28.8 to −3.4) (Figure S3; Table S8). Meta-analysis-restricted item-level results are reported separately (Figure S4; Table S9).

### Reporting Completeness

Mean PRISMA adherence was 0.71 (SD, 0.17) for AI-assisted reviews and 0.79 (SD, 0.15) for reviews with no reported AI use. The adjusted difference was −4.7 percentage points (95% CI, −8.9 to −0.6; P=0.027), and the matched-set fixed-effects difference was −6.4 percentage points (95% CI, −10.7 to −2.1; P=0.004) (Table 2; Figure 2; Figure S5). The largest adjusted domain-level difference was in Results (−7.99 percentage points; 95% CI, −14.56 to −1.43), with item-level findings reported separately (Tables S10-S11; Figure S6).

### Risk of Bias

Among reviews with assessable ROBIS ratings, 57 of 76 AI-assisted reviews (75.0%) and 101 of 154 reviews with no reported AI use (65.6%) were rated low risk. The adjusted difference was +17.8 percentage points (95% CI, 2.7 to 33.0; P=0.022), and the matched-set fixed-effects difference was +16.5 percentage points (95% CI, 0.2 to 32.9; P=0.048) (Table 2; Figure 2; Figure S7). The largest domain-level difference was for synthesis and findings (78.9% vs. 64.9%; adjusted difference, +20.1 percentage points; 95% CI, 6.2 to 34.0), whereas other domains showed no clear differences (Figure S8; Table S12).

### Transparency of AI Use

Among AI-assisted reviews, the mean AITDI score was 3.86 (SD, 1.06). Stage of AI use (100%) and tool identity (98.7%) were usually reported, whereas human oversight (79.2%), prompts or parameter settings (55.8%), AI-related limitations (45.5%), model version or date (42.9%), and data-governance or ethics considerations (9.1%) were less consistently reported (Figure 2; Figure S9; Table S13). AITDI scores were similar across journal tier, review-question type, and stage of AI use (Table S14), and were not associated with AMSTAR-2 scores (adjusted difference per 1-point higher AITDI score, −0.10; 95% CI, −0.64 to 0.43) or PRISMA adherence (−1.3 percentage points; 95% CI, −4.6 to 2.1) (Table S15).

### Timeliness and Dissemination

Receipt-to-publication intervals, 12-month citation counts, and Altmetric Attention Scores did not differ between groups in adjusted analyses; data were available for 193, 208, and 90 reviews, respectively (Table S16).

### Sensitivity and Exploratory Analyses

The primary finding was consistent across sensitivity analyses. Noninferiority was not demonstrated in paired comparisons (difference, −2.05 points; 95% CI, −2.69 to −1.40), analyses restricted to standard systematic reviews (adjusted difference, −1.75; 95% CI, −2.47 to −1.03), leave-one-journal-out analyses (range, −1.82 to −1.54), or overlap-weighted analyses (−2.09; 95% CI, −2.73 to −1.44), with good post-weighting covariate balance (Tables S17-S21). In exploratory stratified models, intervention reviews had higher AMSTAR-2 scores than methodological, qualitative, or other reviews among both AI-assisted reviews (adjusted difference, 2.20 points; 95% CI, 0.73 to 3.67) and reviews with no reported AI use (1.19 points; 95% CI, 0.06 to 2.32). PRISMA adherence showed a similar pattern (Table S22).

## Discussion

Reviews with reported AI assistance had lower adherence to established methodological criteria and less complete standard reporting than matched reviews with no reported AI use, although risk-of-bias ratings were more favorable. Many reviews disclosed that AI was used but did not provide the details needed to reproduce, audit, or interpret the AI-mediated workflow. For clinicians, guideline developers, and policymakers, these findings do not establish that AI use causes unreliable review findings; rather, they identify methodological and reporting deficiencies among published reviews reporting AI assistance. As AI-enabled workflows are adopted more widely, this should be treated as an early warning to preserve core systematic-review standards, including transparent reporting, human verification, reproducibility, and auditability.

These findings reflect current published AI-assisted review practice rather than the future potential of AI in evidence synthesis. AI may improve efficiency in citation screening, data extraction, appraisal, synthesis, and review updating.[3–15] The principal concern is implementation. The largest item-level differences involved defining the review question, justifying eligible study designs, and explaining heterogeneity, suggesting that lower adherence may have reflected weaknesses in foundational review design and reporting rather than AI use alone. The safeguards that make systematic reviews clinically useful, including prespecified methods, comprehensive searches, duplicate processes, transparent excluded-study lists, risk-of-bias assessment, reproducible synthesis, human verification, and detailed reporting, remain essential regardless of whether review tasks are performed by humans or AI-enabled tools.[1,10,11,14,15,18–21,30]

This study shifts evaluation from task-level performance to the quality of completed reviews entering the biomedical literature. That distinction matters because systematic reviews combine interdependent decisions across the full workflow. An AI tool may perform one task accurately yet contribute to an untrustworthy review if its outputs are not prespecified, verified, documented, and traceable to source material. Conversely, AI can be useful as a bounded adjunct within a transparent, human-supervised process.[7–17,30,31]

More broadly, AI-assisted evidence synthesis should be understood as part of the clinical evidence pathway rather than as an isolated technical application. Systematic reviews are not AI tools deployed at the bedside, but they shape guidelines, coverage decisions, and clinical care. The impact of medical AI therefore depends not only on tool performance but also on how AI-enabled workflows are evaluated, reported, integrated into human processes, and translated into practice.[32–35]

The discordance among AMSTAR-2, PRISMA, and ROBIS reflects their different constructs and decision rules.[18–21,36] AMSTAR-2 appraises methodological weaknesses, PRISMA 2020 addresses reporting completeness, and ROBIS evaluates whether review processes may have introduced bias. Lower AMSTAR-2 and PRISMA scores may partly reflect less complete reporting or disclosure, whereas the more favorable ROBIS ratings suggest that these deficiencies were not consistently judged to increase the risk that review processes biased the findings or conclusions.

The practical implications are immediate. When AI contributes substantively to the conduct of a systematic review, its use should be reported in the Methods and, when central to the review, in the abstract rather than only in an acknowledgement, footnote, or generic disclosure. Existing systematic-review reporting standards support transparent reporting of conventional methods but were not designed to capture model identity and version, prompts or parameters, human verification, or data-governance safeguards.[19,20] Emerging guidance, including PRISMA-trAIce, GAMER, and TRIPOD-LLM, is beginning to define expectations for standardized AI disclosure.[22–24] Journals should scrutinize the documentation, human verification, and auditability of substantive AI-assisted review steps, treating incomplete documentation as incomplete methods reporting rather than penalizing transparent disclosure of AI use. For clinicians and guideline panels, AI use alone should not reduce confidence in a review; confidence in reproducibility should depend on whether the human–AI workflow is transparent and auditable.

This study has limitations. Exposure classification depended on reported AI use, so comparators represented reviews with no reported AI use rather than confirmed nonuse. The extent of undisclosed substantive AI use and the direction of any resulting misclassification bias are uncertain. The search-guided matching procedure did not require exact agreement on all characteristics, and differences in review composition and scope remained. Compared with controls, AI-assisted reviews included fewer intervention reviews, more methodological reviews, fewer reviews with meta-analysis, and generally larger evidence bases, as reflected by greater numbers of records retrieved, records screened, and studies included. Although multivariable models adjusted for review-question type and the primary finding remained consistent across sensitivity analyses, including matched-set fixed effects and propensity-score overlap weighting, residual confounding related to review scope or complexity and differential applicability of appraisal criteria across review types may have contributed to the observed associations.

AMSTAR-2 was not designed as a formal additive quality score for individual reviews; the modified AMSTAR-2 total was therefore interpreted as a prespecified group-level methodological-adherence outcome, alongside overall confidence ratings. Appraisers could not be fully blinded to reported AI use, and AMSTAR-2, PRISMA, ROBIS, and AITDI require judgment. Although assessments used piloted forms and duplicate review, detailed item-level rules for classifying PRISMA reporting as complete or partial were not provided, which may limit reproducibility of these judgments. These instruments assess reported methods, reporting completeness, and review processes rather than the accuracy of every review decision or AI-mediated output; complete reporting does not guarantee correct conduct, and incomplete reporting may obscure rigorous conduct. AI assistance encompassed screening classifiers, extraction and appraisal tools, and generative language models, which raise different validity and reproducibility concerns; generative systems may also produce plausible but inaccurate or unverifiable content, including erroneous references.[5–8,31] AITDI is preliminary and requires further validation. Some secondary and exploratory analyses, particularly AITDI associations and meta-analysis-restricted models, had limited precision because of smaller sample sizes. The included reviews were English-language, peer-reviewed health systematic reviews from an early period of AI adoption, which may limit generalizability to future AI-assisted review practice. Descriptive confidence categories used an adaptation of the AMSTAR-2 decision framework to the modified 13-item assessment and should not be interpreted as standard overall confidence ratings.

Future work should link the quality of completed reviews to the specific human–AI workflows used to produce them. Priorities include assessing the reproducibility and source traceability of AI-assisted screening, data extraction, risk-of-bias assessment, and synthesis across repeated runs, models, versions, AI systems, and human reviewers.[15,24,30,31,35] AITDI was applied retrospectively in this study to measure how completely published reviews reported AI use; it is distinct from prospective reporting frameworks, such as PRISMA-trAIce and GAMER, which are intended to guide authors when designing and reporting future reviews.[22–25] Future work should map AITDI against these emerging frameworks and validate whether its domains capture reproducibility, auditability, stakeholder trust, and real-world review performance. Qualitative and consensus-based studies with authors, editors, clinicians, policymakers, and patients are also needed to define acceptable standards for disclosure, supervision, and accountability. These efforts should define how AI can accelerate evidence synthesis while preserving rigor, reproducibility, and clinical trust.

In conclusion, reviews with reported AI assistance had lower methodological and reporting adherence and incomplete AI-specific transparency, despite more favorable risk-of-bias ratings. AI-assisted evidence synthesis should be judged by whether its methods, human oversight, and chain of evidence are auditable and reproducible, because gains in speed are valuable only when the resulting evidence remains trustworthy.

## Use of Artificial Intelligence

During preparation of this manuscript, the authors used ChatGPT (GPT-5.6; OpenAI) to assist with language editing, organization, and minor rephrasing in the manuscript, tables, figure legends, and supplementary materials, as well as troubleshooting, minor code revisions, and improving the organization, commenting, and labeling of the R analysis script. The tool was not used to perform study selection, data extraction, or appraisal, or to independently conduct statistical analyses. Decisions regarding statistical methods and interpretation of study results remained the responsibility of the authors. All AI-assisted content was critically reviewed, revised, and verified by the authors, who retained responsibility for all scientific judgments, interpretations, and conclusions and take full responsibility for the accuracy, integrity, and originality of the final manuscript.

## Data availability statement

The detailed supplementary methods, tables, figures, and references are provided in the supplementary appendix accompanying this preprint. The complete machine-readable study-level dataset and R analysis script have been provided to the journal for peer review but are not publicly available with this preprint. These materials will be made publicly available upon peer-reviewed publication.

## Contributors

**Concept and design:** Jay, Dharma, Yu, Straus, Lipscombe, Eskander.

**Acquisition, analysis, or interpretation of data:** Jay, M. Esttaifo, Samani, Wilson, S. Esttaifo, Morgan, Abi-Jaoude, Reszetnik, Chopra, Alrashidi, Monteiro, Schemitsch, Karthikeyan, Obi, Emdin, Nguyen, Anni, Sutradhar, Dharma, Yu, Straus, Lipscombe, Eskander.

**Drafting of the manuscript:** Jay, M. Esttaifo.

**Critical revision of the manuscript for important intellectual content:** All authors.

**Statistical analysis:** Jay, Sutradhar.

**Administrative, technical, or material support:** Jay, M. Esttaifo, Samani, Wilson.

**Supervision:** Yu, Straus, Lipscombe, Eskander.

All authors approved the final manuscript and agree to be accountable for the work. MJ and AE are the guarantors and had access to the study data. Both take responsibility for the integrity of the data, accuracy of the analyses, and overall content of the manuscript. The use of ChatGPT is described in the Use of Artificial Intelligence section.

## Abbreviations

AITDI: Artificial Intelligence Transparency and Disclosure Index
AMSTAR-2: A Measurement Tool to Assess Systematic Reviews, version 2
PRISMA: Preferred Reporting Items for Systematic Reviews and Meta-Analyses
ROBIS: Risk of Bias in Systematic Reviews

---

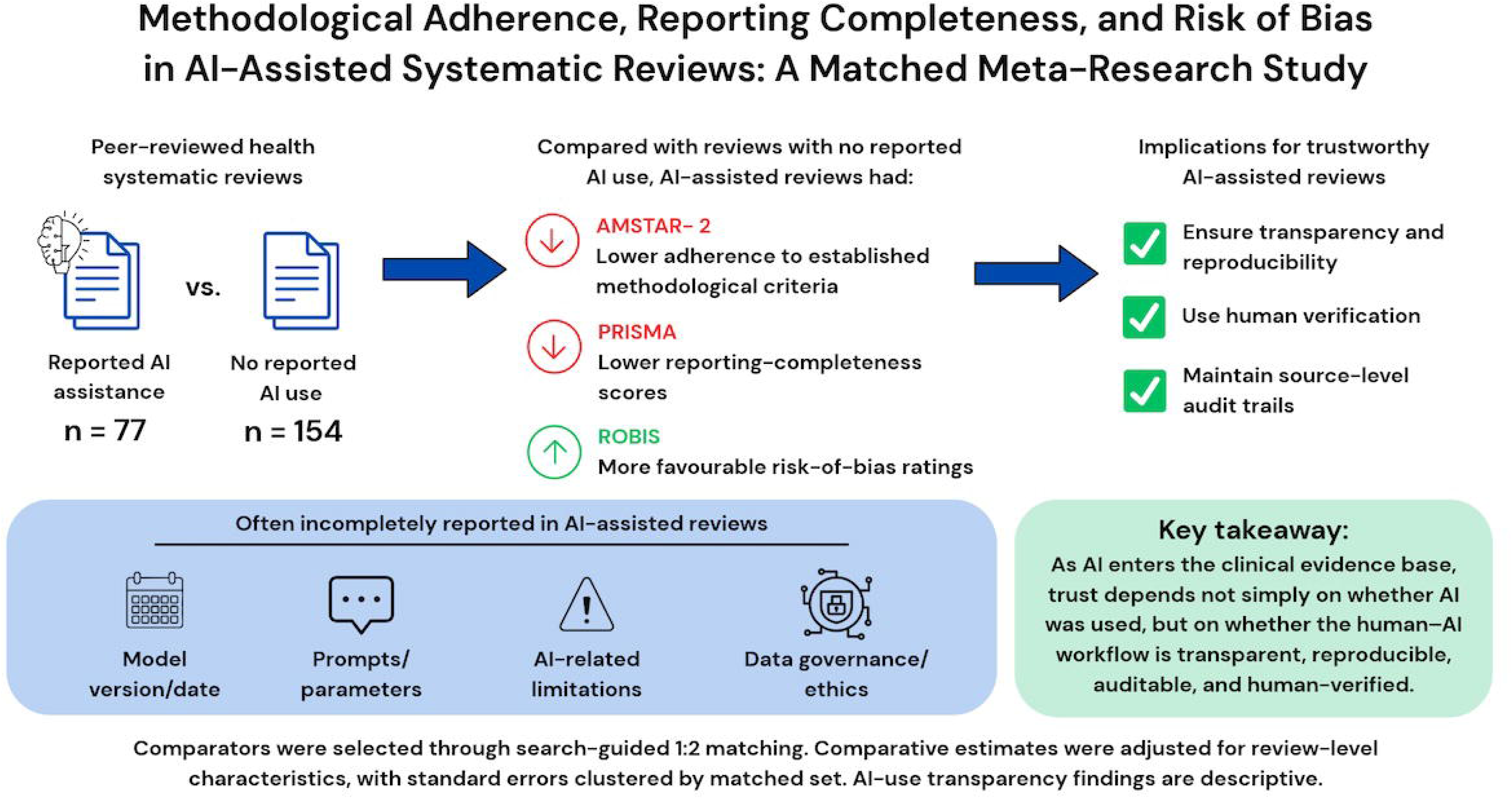

