## Supplementary Methods Tables and Figures for "Methodological Adherence, Reporting Completeness, and Risk of Bias in AI-Assisted Systematic Reviews: A Matched Meta-Research Study"

| **Supplementary Materials Overview** |
| --- |
| **Appendix 1. Supplementary Methods** |
| S1. Relationship to the Published Protocol |
| S2. Information Sources and Search Strategy |
| S3. Study Selection |
| S4. Exposure and Comparator Classification |
| S5. Comparator Identification, Matching, and Random Selection |
| S6. Data Extraction and Quality Control |
| S7. Outcome Definitions and Scoring Rules |
| S8. Covariates and Operational Coding |
| S9. Primary and Secondary Statistical Analyses |
| S10. Timeliness and Dissemination Analyses |
| S11. Sensitivity and Exploratory Analyses |
| S12. Deviations and Clarifications Relative to the Published Protocol |
| **Appendix 2. Detailed Search Strategies** |
| Table S1. Expanded characteristics of the included systematic reviews |
| Table S2. Individual-study characteristics of the included systematic reviews |
| Table S3. Individual-study methodological quality, reporting, risk-of-bias, and AI-transparency outcomes |
| Figure S1. Distribution of AMSTAR-2 13-item scores |
| Figure S2. Distribution of AMSTAR-2 16-item scores |
| Table S4. Adapted 13-item AMSTAR-2 confidence categories |
| Table S5. Descriptive AMSTAR-2 domain scores |
| Table S6. Adjusted AMSTAR-2 domain analyses |
| Table S7. AMSTAR-2 domain analyses among reviews with meta-analysis |
| Figure S3. AMSTAR-2 item-level adjusted differences |
| Table S8. Item-level AMSTAR-2 adherence |
| Figure S4. AMSTAR-2 item-level differences among reviews with meta-analysis |
| Table S9. Item-level AMSTAR-2 adherence among reviews with meta-analysis |
| Figure S5. Distribution of PRISMA adherence |
| Table S10. PRISMA domain-level analyses |
| Figure S6. PRISMA item-level adjusted differences |
| Table S11. PRISMA item-level analyses |
| Figure S7. ROBIS overall low-risk classifications |
| Figure S8. ROBIS domain-level differences |
| Table S12. ROBIS domain-level analyses |
| Figure S9. AITDI item-level reporting |
| Table S13. AITDI item-level reporting |
| Table S14. AITDI total scores by selected characteristics |
| Table S15. Associations between AITDI and quality or reporting |
| Table S16. Timeliness and dissemination analyses |
| Table S17. Matched-set paired sensitivity analysis |
| Table S18. Standard systematic review sensitivity analysis |
| Table S19. Leave-one-journal-out sensitivity analysis |
| Table S20. Overlap-weighted sensitivity analysis |
| Table S21. Balance diagnostics for overlap weighting |
| Table S22. Exploratory stratified predictor models |
| Table S23. Model-complexity assessment |
| **Appendix 3. References for Included Systematic Reviews**  **Appendix 4. Reporting Checklist for Meta-Epidemiological Methodology Research** |

### **Supplementary Materials Overview**

This Supplementary Appendix contains the supplemental methods, complete electronic search strategies, supplementary tables and figures, and a self-contained reference list. Items are arranged in the order in which they are first cited in the main manuscript.

Tables S2 and S3 provide a human-readable study-level listing of the included reviews and their principal appraisal outcomes. Multimedia Appendix 2 contains the complete machine-readable study-level dataset, including domain- and item-level appraisal scores. Multimedia Appendix 3 contains the complete R analysis script. Appendix 3 provides full references for all included reviews.

The complete Excel study-level dataset and R analysis script were supplied separately for journal peer review and are not publicly included with this preprint.

### **Appendix 1. Supplementary Methods**

#### **S1. Relationship to the Published Protocol**

This report presents Aim 1 of the previously published four-aim mixed-methods meta-research protocol.[1] The principal prespecified design features were retained: comparison of systematic reviews with reported artificial intelligence (AI) assistance with reviews with no reported AI use; selection of two comparator reviews for each AI-assisted review from a similar temporal, clinical, and methodological context; use of a modified 13-item AMSTAR-2 adherence score as the primary outcome; and a noninferiority margin of −1.0 point.

The comparator-identification process was operationalized through individualized Ovid MEDLINE searches constructed by an information specialist. These searches were designed to identify potentially eligible comparator reviews published within 3 months before or after the corresponding AI-assisted review and from a similar broad clinical and methodological context, considering clinical domain, review type, and presence or absence of meta-analysis. When more than two potentially eligible comparators were identified, two were selected using reproducible computer-generated random sampling with a fixed seed of 42.

Additional operational clarifications and analytic modifications included analysis of ROBIS as low risk versus not low risk; use of available-case analyses rather than multiple imputation; use of receipt-to-publication interval as the available measure of publication timeliness; and addition of paired matched-set, matched-set fixed-effects, domain-level, item-level, leave-one-journal-out, and overlap-weighted analyses. These modifications and their rationales are summarized in the protocol-deviation table.

#### **S2. Information Sources and Search Strategy**

An experienced information specialist developed and conducted searches in MEDLINE through Ovid, CINAHL through EBSCOhost, and the Cochrane Database of Systematic Reviews to identify potentially eligible AI-assisted systematic reviews. Searches were conducted on December 4, 2025, in MEDLINE and the Cochrane Database of Systematic Reviews and on December 5, 2025, in CINAHL. The strategies combined systematic-review terms with terms related to artificial intelligence, machine learning, natural language processing, large language models, and automated evidence-synthesis tools.

The Ovid MEDLINE strategy was peer reviewed by an independent information specialist using the Peer Review of Electronic Search Strategies 2015 Guideline.[2] Recommended revisions were incorporated before the strategy was finalized and translated for CINAHL and the Cochrane Database of Systematic Reviews. A separate systematic-review filter was not applied in the Cochrane Database of Systematic Reviews because the database consists of systematic reviews.

AI-assisted reviews were eligible if published from January 1, 2023, through December 31, 2025. Comparator reviews could be published from October 1, 2022, through March 31, 2026, to allow selection within 3 months before or after the corresponding AI-assisted review. Complete database-specific strategies, platforms, coverage dates, search dates, and line-level retrieval counts are provided in Appendix 2.

#### **S3. Study Selection**

Records identified through the AI-targeted search underwent duplicate title-and-abstract screening followed by duplicate full-text assessment in Covidence. Before formal screening, reviewers completed calibration exercises using approximately 10–20 records for title-and-abstract screening and five records for full-text screening. Reviewers compared decisions and discussed discrepancies, with a target agreement of at least 80% before proceeding to independent screening. Screening guidance was refined when disagreements identified differences in the interpretation of the eligibility criteria.

During title-and-abstract screening, reviewers assessed whether each record represented a potentially eligible systematic review addressing a human-health, public-health, health-care-delivery, or health-system topic. At full-text assessment, reviewers applied all eligibility criteria and examined the main article, supplementary materials, acknowledgements, disclosures, author-contribution statements, and footnotes for reported AI use. Disagreements were resolved through discussion, with unresolved disagreements adjudicated by a third reviewer. AI-use classification was based on explicit reporting in the available article and supplementary materials. No cases were sufficiently ambiguous to require contact with the corresponding author.

After an AI-assisted review was confirmed as eligible, an information specialist constructed an individualized Ovid MEDLINE search to generate a candidate pool of potentially eligible comparator reviews. Two records were selected from each candidate pool using reproducible computer-generated random sampling and underwent duplicate full-text assessment. Reviewers confirmed general eligibility and the absence of reported AI use in the article and supplementary materials. When a selected record was ineligible, it was replaced using the next record in the randomized candidate order. The final analytic cohort included 77 AI-assisted reviews and 154 comparator reviews in 77 matched sets.

#### **S4. Exposure and Comparator Classification**

The exposure was author-reported AI assistance. A review was classified as AI-assisted when the available published materials explicitly reported use of artificial intelligence, machine learning, natural language processing, a large language model, or an AI-enabled automation tool at a systematic-review stage. Eligible stages included search development or information retrieval; title, abstract, or full-text screening; data extraction; risk-of-bias or quality appraisal; qualitative or quantitative synthesis or analysis; substantive manuscript drafting or interpretation; or multiple stages of the systematic-review workflow.

Use of conventional software without reported AI-enabled functionality, including Covidence, RevMan, reference-management software, spreadsheet software, and conventional statistical software, was not classified as AI assistance. Application of an AI or machine-learning method solely to a separate analytic component outside the systematic-review workflow was also not classified as AI-assisted review conduct.

Comparator reviews met the same general eligibility criteria but contained no reported AI, machine-learning, natural-language-processing, or large-language-model use in the main article or supplementary materials. Comparator reviews are therefore interpreted as reviews with no reported AI use rather than reviews in which AI use was definitively absent.

AI use was additionally classified by workflow stage and broad tool type for descriptive analyses. Broad tool categories included large-language-model-based tools, screening or classification tools, combined large-language-model and screening tools, and other or mixed AI tools.

#### **S5. Comparator Identification, Matching, and Random Selection**

#### Each AI-assisted review was matched to two reviews with no reported AI use. For each AI-assisted review, an information specialist constructed an individualized Ovid MEDLINE search to identify potentially eligible comparators from a similar publication period and broad clinical and methodological context.

#### Candidate comparator reviews were intended to have been published within 3 months before or after the corresponding AI-assisted review. Clinical domain was defined broadly rather than according to the precise review question. Searches incorporated relevant Medical Subject Headings, indexed terms, title and abstract terminology, and journal specialty when useful. Review type and presence or absence of meta-analysis were considered when constructing or assessing the candidate pools in accordance with the prespecified matching framework.

#### The individualized searches were intended to generate sufficiently broad candidate pools for comparator selection rather than to identify all systematic reviews within each clinical domain. Under the standard procedure, when more than two potentially eligible comparators were identified, two were selected using computer-generated random sampling with a fixed seed of 42.

#### Selected comparator records underwent duplicate full-text assessment to confirm the general eligibility criteria and absence of reported AI use in the article or supplementary materials. Under the standard procedure, an ineligible randomly selected record was replaced using the next record in the reproducibly randomized candidate order until two eligible comparators were identified. Each AI-assisted review and its two comparator reviews were assigned a matched-set identifier. Matched-set identifiers were retained for cluster-robust variance estimation, paired analyses, and matched-set fixed-effects sensitivity analyses.

The publication-date window was intended as a standard restriction, subject to the exceptions described below. Clinical domain, review type, and meta-analysis status informed candidate identification rather than requiring exact agreement or a prespecified minimum number of concordant characteristics. Consequently, the selection procedure did not ensure balance across these characteristics.

Two replacement comparators were selected through additional topic-based PubMed searches outside the original candidate pools. These searches used the PubMed publication type “Systematic Review,” excluded records tagged “Meta-Analysis” or “Scoping Review,” and required the “Humans” MeSH heading. Candidate reviews were additionally checked manually for relevance to a human clinical topic and availability of full text. The intended publication-date window was not applied.

For Rakhshandehroo (study 4580), one comparator required replacement, and the two remaining unused candidates in the original pool did not meet these criteria. Narimani (published December 24, 2025, approximately 2.5 years after the AI-assisted review) was selected through the additional PubMed search. For Canfell (study 3464), the original candidate file was incorrectly labelled “3465,” preventing its identification by the automated selection notebook. Miake-Lye (published October 5, 2023) was therefore selected through an additional PubMed search. Subsequent checking established that the original candidate file contained eligible unused comparators within the intended publication window. Both replacement comparators fell outside their intended publication windows.

A retained summary note documented the additional-search approach and filters, but the exact search terms, result lists, and rationale for selecting these particular reviews were unavailable. Selection could therefore not be fully reconstructed or verified as random. Both comparators were retained in the final analysis and are documented as deviations from the intended selection procedure. A subsequent audit confirmed that the other 152 comparators originated from the original candidate pools for their corresponding AI-assisted reviews.

#### **S6. Data Extraction and Quality Control**

Data were extracted using standardized and piloted forms. Review characteristics, AI-use characteristics, AMSTAR-2 assessments, PRISMA 2020 assessments, and ROBIS ratings were extracted independently by two reviewers. Reviewer pairs compared their initial extractions and discussed differences to establish a common interpretation of the coding guidance before completing their remaining assignments. Discrepancies were resolved through discussion, with unresolved disagreements adjudicated by a third reviewer. The completed dataset was additionally reviewed for completeness and consistency before analysis.

Review-level characteristics included publication year, journal impact tier, funding reporting, protocol registration or prespecified methods, information-specialist involvement, meta-analysis status, review type, review-question type, included study designs, corresponding-author country, clinical domain, number of databases searched, numbers of records retrieved and screened, number of included studies, and review-team size.

For AI-assisted reviews, additional fields captured tool identity, tool type, model version or date, stage of AI use, prompts or parameter settings, reported human oversight, data-governance or ethics considerations, and acknowledgement of AI-related limitations. The main article, supplementary materials, and any cited or linked review protocol were considered part of the review report when extracting characteristics and applying the appraisal tools.

#### **S7. Outcome Definitions and Scoring Rules**

***AMSTAR-2 Adherence Scores***

Methodological adherence was assessed using AMSTAR-2,[3] a 16-item instrument evaluating the conduct and reporting of systematic reviews. The primary outcome was a modified 13-item AMSTAR-2 adherence score applicable to reviews with and without meta-analysis. It included AMSTAR-2 items 1 through 10, 13, 14, and 16. Items 11, 12, and 15 were excluded because they apply specifically to reviews conducting quantitative synthesis. The score ranged from 0 to 13, with higher scores indicating greater methodological adherence. AMSTAR-2 was not designed to generate a formal additive quality score for individual reviews; therefore, the modified 13-item and full 16-item totals were used as prespecified group-level methodological-adherence outcomes for comparative meta-research, not as replacements for standard AMSTAR-2 overall confidence ratings.

AMSTAR-2 permits “Partial Yes” ratings for some items. For the modified 13-item and full 16-item adherence totals used in this meta-research study, only a full “Yes” response contributed one point; “Partial Yes” and “No” responses were coded as nonadherent. Item-level adherence analyses similarly classified only a full “Yes” response as adherent. This approach avoided assigning subjective partial credit across heterogeneous items and preserved a transparent item-level adherence metric for group-level comparison.

For analyses of the 13-item score, the five critical items were items 2, 4, 7, 9, and 13. The eight noncritical items were items 1, 3, 5, 6, 8, 10, 14, and 16. Critical- and noncritical-domain outcomes were expressed both as the number of items satisfied and as the proportion of included items satisfied.

Among reviews with meta-analysis, a secondary 16-item adherence score included all AMSTAR-2 items. For these analyses, the seven critical items were items 2, 4, 7, 9, 11, 13, and 15, and the nine noncritical items were items 1, 3, 5, 6, 8, 10, 12, 14, and 16. AMSTAR-2 domain- and item-level analyses were exploratory and were not adjusted for multiple comparisons.

***AMSTAR-2 Overall Confidence Ratings***

Confidence categories were recalculated descriptively from the reconciled modified 13-item ratings using an adapted AMSTAR-2 framework. Critical items were 2, 4, 7, 9, and 13; items 11, 12, and 15 were excluded. Only a full Yes satisfied an item; Partial Yes and No counted as weaknesses. High indicated no critical weaknesses and at most one noncritical weakness; moderate, no critical weaknesses and more than one noncritical weakness; low, one critical weakness; and critically low, more than one critical weakness. These adapted categories are not original AMSTAR-2 overall confidence ratings.

***PRISMA 2020 Adherence***

Reporting completeness was assessed using the 27-item PRISMA 2020 checklist.[4] Each item was scored as 1 for complete reporting, 0.5 for partial reporting, and 0 for absent reporting. The total numeric score was divided by 27 to produce an adherence proportion ranging from 0 to 1. Unlike the AMSTAR-2 adherence score, which counted only fully satisfied methodological criteria, the PRISMA score was intended to capture degrees of reporting completeness; partial credit distinguished incomplete reporting from complete omission.

PRISMA domains comprised title and abstract, introduction, methods, results, discussion, and other information. Domain-level adherence was calculated as the sum of item scores within the domain divided by the maximum possible score for that domain. Primary PRISMA item-level analyses used the numeric item scores and therefore preserved partial credit. Domain- and item-level analyses were exploratory and were not adjusted for multiple comparisons.

***ROBIS***

Risk of bias in the review process was assessed using ROBIS,[5] which evaluates four domains: study-eligibility criteria; identification and selection of studies; data collection and study appraisal; and synthesis and findings. Overall and domain-level judgments were classified as low, high, unclear, not applicable, or missing. For comparative analyses, low risk was coded as 1, and high or unclear risk was coded as 0. Overall and domain-level outcomes were analyzed separately using the available judgment for each outcome. One AI-assisted review had an overall ROBIS judgment of not applicable and was excluded from the overall binary ROBIS analysis. The overall ROBIS analysis therefore included 76 AI-assisted reviews and 154 comparator reviews.

***Artificial Intelligence Transparency and Disclosure Index***

The preliminary Artificial Intelligence Transparency and Disclosure Index was developed to assess the completeness of AI-specific reporting in AI-assisted systematic reviews.[1] It was developed through an iterative, concept-driven evidence-mapping and analytic synthesis process drawing on three inputs: existing AI-related reporting standards, extensions, and policy-level guidance relevant to research transparency and evidence synthesis; recurrent transparency gaps observed in published AI-assisted systematic reviews; and foundational principles of reproducibility in systematic reviews, including the need for sufficient methodological detail to permit independent appraisal and repetition of the review process. These inputs were mapped across the systematic-review workflow to identify common transparency gaps and informed six scored AITDI domains and one nonscored contextual domain. The AITDI was used in this study as a preliminary analytic framework to support meta-research and comparative evaluation of current reporting practice. It was not intended to serve as a formal reporting guideline, a validated quality measure, or a replacement for established systematic-review appraisal tools. The six scored items were tool identity; model version or date; stage of AI use; prompts, parameters, thresholds, or configuration details; human oversight or verification; and data-governance or ethics considerations. Each item was coded as 1 when the minimum reporting criterion was satisfied and 0 when it was not satisfied, producing a total score ranging from 0 to 6. Reporting of AI-related limitations was summarized as an additional nonscored item and was not incorporated into the 6-item total.

#### **S8. Covariates and Operational Coding**

The primary adjusted models included journal impact tier, funding reporting, review-group affiliation, review-team size, and review-question type. These variables were selected as measurable indicators of review resources, reporting infrastructure, methodological context, and the likelihood of AI adoption.

Journal impact tier was obtained from the SCImago Journal & Country Rank database at the time of extraction and categorized as Q1, Q2, Q3, Q4, or unknown or not ranked. Funding reporting indicated whether the review included a funding statement and was categorized as reported or not reported. This variable represented the presence of funding reporting rather than classification of the source as industry or nonindustry.

Review-group affiliation was coded as present when the review was explicitly identified as affiliated with Cochrane or another formally named evidence-synthesis or review group. Reviews without a reported formal affiliation were coded as not affiliated. Review-team size was defined as the number of listed manuscript authors. No covariate imputation was performed. Unavailable or unclear values for categorical adjustment covariates were retained as an “Unknown” category. Cochrane reviews were coded as affiliated regardless of whether a listed author had a separate Cochrane institutional affiliation.

Review-question type was categorized as intervention, diagnostic, etiology or association, prognostic, prevalence, qualitative, methodological, or other or mixed. Intervention reviews constituted the reference category in adjusted models. Publication timing, clinical domain, review type, and meta-analysis status were considered during comparator identification and were also summarized descriptively. They were not included as covariates in the primary adjusted model. Information-specialist involvement was summarized descriptively but was not included in the primary adjusted model because it was incompletely or inconsistently reported.

#### **S9. Primary and Secondary Statistical Analyses**

#### Continuous variables were summarized using means with standard deviations or medians with interquartile ranges, as appropriate. Categorical variables were summarized using counts and percentages.

#### The primary estimand was the adjusted mean difference in the AMSTAR-2 13-item score between AI-assisted reviews and comparator reviews, expressed as AI-assisted minus comparator. Ordinary least-squares regression was used. Confidence intervals and hypothesis tests used CR2 cluster-robust variance estimation, with matched set as the clustering unit and Satterthwaite small-sample degrees-of-freedom corrections. The adjusted model included AI exposure, journal impact tier, funding reporting, review-group affiliation, review-team size, and review-question type.

#### Noninferiority was considered demonstrated only if the lower limit of the two-sided 95% confidence interval for the AI-minus-comparator mean difference was greater than the prespecified margin of −1.0 AMSTAR-2 point. The same criterion was applied to unadjusted and sensitivity estimates of the 13-item outcome. No noninferiority margin was applied to secondary outcomes.

#### The 16-item AMSTAR-2 adherence score was analyzed using the same regression approach among reviews with meta-analysis. Adapted 13-item AMSTAR-2 confidence categories were summarized descriptively by review group. PRISMA adherence was analyzed using linear regression, with estimates reported as absolute percentage-point differences. Overall and domain-level ROBIS outcomes were analyzed using linear probability models, with low risk as the favorable outcome. Estimates were reported as absolute percentage-point differences. Matched-set fixed-effects models included indicators for matched set in addition to the adjustment covariates. These models were treated as sensitivity analyses because the number of matched-set parameters was large relative to the cohort size.

#### For each principal adjusted model, we counted the number of estimated regression parameters, excluding the intercept, and calculated the number of observations per parameter. These values are reported in Table S23. We did not apply this calculation to matched-set fixed-effects models because those models include separate indicators for matched sets, which substantially increase the number of parameters. Analyses used available observations for the outcome under analysis. Missing outcomes were not imputed. Analyses were conducted in R. CR2 cluster-robust variance estimators and Satterthwaite small-sample corrections were implemented using the clubSandwich package.

#### **S10. Timeliness and Dissemination Analyses**

Because dates reflecting protocol registration or initiation of review conduct were not consistently available, publication timeliness was operationalized as the number of days from manuscript receipt by the journal to publication. This outcome represents the journal submission-to-publication process and was not interpreted as the time required to conduct the systematic review.

Reviews with missing receipt-to-publication intervals were excluded. Absolute differences in days were estimated using linear regression. A log-linear model of the natural logarithm of the interval was also fitted, with coefficients transformed as 100 × [exp(β) − 1] to obtain relative percentage differences. Models adjusted for journal impact tier and publication year and used cluster-robust variance estimation by matched set.

Early dissemination was assessed using 12-month citation counts and Altmetric Attention Scores among reviews with available data. Each outcome was transformed as log (1 + outcome) and modeled using linear regression adjusted for journal impact tier and publication year. Coefficients were exponentiated and reported as relative percentage differences using 100 × [exp(β) − 1]. Timeliness and dissemination analyses were exploratory. Reviews with unavailable outcome data were excluded from the corresponding analysis.

#### **S11. Sensitivity and Exploratory Analyses**

For the paired matched-set analysis, the AMSTAR-2 13-item score of each AI-assisted review was compared with the mean score of its two matched comparators. The resulting 77 matched-set differences were summarized, and one-sample t tests evaluated the mean difference against both 0 and the noninferiority margin of −1.0.

The standard-review sensitivity analysis excluded reviews not classified as standard systematic reviews. Adjusted and matched-set fixed-effects models otherwise used the primary model specification. The number of retained reviews and matched sets with within-set exposure contrasts was reported. The restriction followed recorded review-type labels; one AI-assisted review labelled only “Systematic” was not included in this stricter restriction. Leave-one-journal-out analyses sequentially removed all matched sets containing each journal that contributed at least three reviews to the cohort and refitted the adjusted primary model.

For the overlap-weighted analysis, a logistic propensity-score model estimated the probability of reported AI assistance using Q1 journal status, funding reporting, and review-team size. AI-assisted reviews received a weight of 1 minus the propensity score, and comparator reviews received a weight equal to the propensity score. Covariate balance was assessed using standardized mean differences before and after weighting. The weighted AMSTAR-2 model used CR2 cluster-robust variance estimation with matched set as the clustering unit. Effective sample sizes and the distribution of weights were also examined.

AMSTAR-2 critical- and noncritical-domain analyses, item-level AMSTAR-2 analyses, PRISMA domain- and item-level analyses, and ROBIS domain-level analyses used the same adjustment covariates as the primary model unless otherwise specified. These analyses were exploratory and were not adjusted for multiple comparisons.

Exploratory models stratified by review group assessed associations of review-question type, review-team size, and Q1 journal status with AMSTAR-2 and PRISMA scores. Conventional model-based standard errors were used for models restricted to AI-assisted reviews because each matched set contributed only one AI-assisted review. For models restricted to non-AI reviews, CR2 cluster-robust standard errors were estimated using matched set as the clustering unit, with Satterthwaite small-sample corrections. Among AI-assisted reviews, additional models examined associations between the 6-item AITDI total and AMSTAR-2 or PRISMA adherence. These analyses were exploratory and intended to identify potential factors associated with methodological or reporting quality rather than to establish causal relationships.

#### **S12. Deviations and Clarifications Relative to the Published Protocol**

The principal study design and primary outcome remained consistent with the published protocol. The following operational clarifications and modifications were made during completion of the study.

| **Protocol specification** | **Final approach** | **Rationale and interpretation** |
| --- | --- | --- |
| Comparator reviews were to be selected according to publication timing, clinical domain, review type, and meta-analysis status. | For each AI-assisted review, an information specialist constructed an individualized Ovid MEDLINE candidate pool considering the prespecified matching characteristics. Candidate reviews were restricted to a publication window of 3 months before or after the corresponding AI-assisted review and were selected from a similar broad clinical and methodological context. When multiple potentially eligible comparators were available, two were selected using reproducible computer-generated random sampling with a fixed seed of 42. | The prespecified matching framework was retained but operationalized through individualized candidate-pool searches followed by random selection rather than exact algorithmic matching on every characteristic. This approach reduced investigator discretion when multiple candidate comparators were available. |
| AI use included both methodological and supporting uses, including language editing. | Reviews were required to report AI involvement in a substantive stage of systematic-review conduct, analysis, synthesis, or interpretation. | The exposure definition was refined to distinguish AI involvement that could plausibly influence review methods, evidence selection, analysis, interpretation, or conclusions from language-only assistance that did not alter the underlying review process. Including language-editing-only use would have grouped minimally exposed reviews with reviews using AI for core methodological tasks, increasing exposure heterogeneity and reducing the interpretability of between-group comparisons. |
| Up to 150 AI-assisted reviews were anticipated, with possible expansion of the search if fewer reviews were identified. | The final cohort included all 77 AI-assisted reviews meeting the final exposure definition and eligibility criteria for which complete 1:2 comparator matching was achieved. The search was not expanded beyond the prespecified databases, publication period, or eligibility criteria. | The protocol target represented an anticipated maximum rather than a required minimum. All eligible reviews identified through the prespecified search were included. Expanding the databases or publication period would have changed the planned study scope and was not feasible within available study resources. The smaller-than-anticipated cohort is considered when interpreting the precision of secondary and subgroup analyses. |
| Funding source was to be extracted and classified as industry versus nonindustry for inclusion in adjusted analyses. | Detailed funding information was initially extracted from published reports. During data cleaning, the analytic variable was simplified to whether a funding statement was reported. | Funding disclosures varied widely in form and often included mixed, institutional, governmental, charitable, author-level, or no-funding statements, making a consistent binary industry-versus-nonindustry classification difficult. Presence of a funding statement could be applied consistently across reviews and was therefore used as the adjustment covariate. |
| Cochrane affiliation was specified as an adjustment variable. | Affiliation with Cochrane or another explicitly named formal review group was coded as present or absent. | The coding was broadened to capture formal review infrastructure beyond Cochrane while preserving the intended construct of organized evidence-synthesis support and oversight. |
| ROBIS was to be analyzed using ordinal or multinomial regression. | ROBIS was analyzed as low risk versus high or unclear risk using linear probability models. Ratings of not applicable or missing were excluded from the corresponding analysis. | Some ROBIS categories contained relatively few reviews, limiting stable ordinal or multinomial modeling. The binary low-risk contrast provided a directly interpretable absolute percentage-point difference. |
| Multiple imputation was proposed for remaining missing data. | Analyses used available observations for each outcome. The principal adjustment covariates were complete and required no imputation. | Missingness was primarily outcome-specific, including unavailable dissemination measures and not-applicable appraisal ratings. Imputation of methodological-quality or reporting outcomes was not considered appropriate. |
| Publication timeliness was to be measured from protocol registration or the earliest available indication of review initiation to publication. | Timeliness was measured as the interval from journal receipt to publication. | Dates reflecting initiation of review conduct were not consistently available. Receipt-to-publication time was therefore used as an available journal-process measure and was not interpreted as the duration required to conduct the systematic review. |
| Negative-binomial or linear models were contemplated for citation and Altmetric outcomes. | Citation counts and Altmetric Attention Scores were transformed using log(1 + outcome) and analyzed using linear regression, with coefficients converted to relative percentage differences. | The transformation accommodated zero values and reduced the influence of strongly right-skewed distributions while permitting interpretable relative estimates. These analyses were exploratory. |
| A sensitivity analysis restricted to reviews using AI for a core methodological stage was planned. | A separate core-stage sensitivity analysis was not performed. | The final exposure definition already excluded reviews in which AI use was limited to non-substantive language editing or readability support. The language-editing-only matched set was removed before the primary analysis, making a further restriction largely redundant (see second row of this table) |
| A sensitivity analysis restricted to comparator reviews explicitly reporting that no AI was used was planned. | This sensitivity analysis was not performed. | Explicit statements of AI nonuse were not captured as a structured variable during data extraction. The analysis would therefore have required retrospective re-extraction from all comparator reports and was not undertaken. Comparator classification remained based on the absence of reported AI use in the article and supplementary materials. |
| Paired matched-set, matched-set fixed-effects, AMSTAR-2 domain- and item-level, PRISMA domain- and item-level, leave-one-journal-out, overlap-weighted, and stratified analyses were not all fully specified in the published protocol. | These analyses were added as sensitivity or exploratory analyses. | The additional analyses evaluated the robustness, distribution, and potential sources of the primary findings. They were interpreted as secondary and were not adjusted for multiple comparisons. |

### **Appendix 2. Detailed Search Strategies**

Searches were conducted on December 4, 2025, in Ovid MEDLINE and the Cochrane Database of Systematic Reviews and on December 5, 2025, in CINAHL. Searches were limited to records published from 2023 onward. The number of records retrieved at each line is shown before deduplication.

Search Strategy A. MEDLINE

Database and platform: Ovid MEDLINE(R) ALL <1946 to December 04, 2025>. Search date: December 4, 2025.

| **Line** | **Search terms** | **Results** |
| --- | --- | --- |
| 1 | Artificial Intelligence/ or Generative Artificial Intelligence/ or Machine Learning/ or Deep Learning/ or Natural Language Processing/ | 155490 |
| 2 | Large Language Models/ | 1431 |
| 3 | ("artificial intelligence" adj5 ("research question*" or search* or criteria or select* or evaluat* or synthesiz* or writing or write* or summariz* or screen* or duplicat* or analy*)).mp. | 8813 |
| 4 | (("AI" or "GenAI" or "generative artificial intelligence" or "machine learning" or "deep learning" or "natural language processing" or "large language model*" or LLM*) adj5 ("research question*" or search* or criteria or select* or evaluat* or synthesi#* or writing or write* or summari#* or screen* or duplicat* or analy*)).mp. | 56759 |
| 5 | (automat* adj3 (search* or extract* or select* or synthesi#* or interpret* or screen* or duplicat* or analy*)).mp. | 52361 |
| 6 | ("machine-assisted" adj5 ("research question*" or search* or criteria or select* or evaluat* or synthesiz* or writing or write* or summariz* or screen* or duplicat* or analy*)).mp. | 32 |
| 7 | (chatgpt* or OpenAI or GPT3 or GPT-3 or GPT4 or GPT-4 or gemini or claude or claude2 or "claude 2" or "microsoft copilot" or DeepSeek or mistral or mixtral or "BLOOM ai" or Llama or LLama2 or "perplexity ai" or distillerAI or "rayyan AI" or "elicit AI" or "research rabbit" or "undermind AI" or "litmaps" or "consensus AI" or "open evidence" or "Nvivo AI").mp. | 16204 |
| 8 | 1 or 2 or 3 or 4 or 5 or 6 or 7 | 247497 |
| 9 | (systematic review or meta-analysis).pt. | 400048 |
| 10 | meta-analysis/ or systematic review/ or systematic reviews as topic/ or meta-analysis as topic/ or "meta analysis (topic)"/ or "systematic review (topic)"/ or exp technology assessment, biomedical/ or network meta-analysis/ | 447355 |
| 11 | ((systematic* adj3 (review* or overview*)) or (methodologic* adj3 (review* or overview*))).ti,ab,kf. | 452417 |
| 12 | ((quantitative adj3 (review* or overview* or synthes*)) or (research adj3 (integrati* or overview*))).ti,ab,kf. | 21607 |
| 13 | ((integrative adj3 (review* or overview*)) or (collaborative adj3 (review* or overview*)) or (pool* adj3 analy*)).ti,ab,kf. | 49601 |
| 14 | (met analy* or metanaly* or technology assessment* or HTA or HTAs or technology overview* or technology appraisal*).ti,ab,kf. | 14855 |
| 15 | (meta-analy* or metaanaly* or systematic review* or biomedical technology assessment* or bio-medical technology assessment*).mp,hw. | 613602 |
| 16 | (cochrane or (health adj2 technology assessment) or evidence report).jw. | 23026 |
| 17 | (meta-analysis or systematic review).mp. | 573306 |
| 18 | (mixed adj3 treatment adj3 (meta-analy* or metaanaly*)).ti,ab,kf. | 189 |
| 19 | umbrella review*.ti,ab,kf. | 3261 |
| 20 | (multi* adj2 paramet* adj2 evidence adj2 synthesis).ti,ab,kf. | 16 |
| 21 | (multiparamet* adj2 evidence adj2 synthesis).ti,ab,kf. | 20 |
| 22 | (multi-paramet* adj2 evidence adj2 synthesis).ti,ab,kf. | 14 |
| 23 | 9 or 10 or 11 or 12 or 13 or 14 or 15 or 16 or 17 or 18 or 19 or 20 or 21 or 22 | 697048 |
| 24 | 8 and 23 | 7542 |
| 25 | limit 24 to yr="2023 -Current" | 5007 |

Search Strategy B. Cochrane Database of Systematic Reviews

Database and platform: EBM Reviews - Cochrane Database of Systematic Reviews <2005 to December 3, 2025>. Search date: December 4, 2025. A separate systematic-review filter was not applied because the database consists of systematic reviews.

| **Line** | **Search terms** | **Results** |
| --- | --- | --- |
| 1 | ("Artificial Intelligence" or "Generative Artificial Intelligence" or "Machine Learning" or "Deep Learning" or "Natural Language Processing").kw. | 3 |
| 2 | Large Language Models.kw. | 0 |
| 3 | ("artificial intelligence" adj5 ("research question*" or search* or criteria or select* or evaluat* or synthesiz* or writing or write* or summariz* or screen* or duplicat* or analy*)).af. | 24 |
| 4 | (("AI" or "GenAI" or "generative artificial intelligence" or "machine learning" or "deep learning" or "natural language processing" or "large language model*" or LLM*) adj5 ("research question*" or search* or criteria or select* or evaluat* or synthesi#* or writing or write* or summari#* or screen* or duplicat* or analy*)).af. | 132 |
| 5 | (automat* adj3 (search* or extract* or select* or synthesi#* or interpret* or screen* or duplicat* or analy*)).af. | 171 |
| 6 | ("machine-assisted" adj5 ("research question*" or search* or criteria or select* or evaluat* or synthesiz* or writing or write* or summariz* or screen* or duplicat* or analy*)).af. | 0 |
| 7 | (chatgpt* or OpenAI or GPT3 or GPT-3 or GPT4 or GPT-4 or gemini or claude or claude2 or "claude 2" or "microsoft copilot" or DeepSeek or mistral or mixtral or "BLOOM ai" or Llama or LLama2 or "perplexity ai" or distillerAI or "rayyan AI" or "elicit AI" or "research rabbit" or "undermind AI" or "litmaps" or "consensus AI" or "open evidence" or "Nvivo AI").af. | 113 |
| 8 | 1 or 2 or 3 or 4 or 5 or 6 or 7 | 400 |
| 9 | limit 8 to yr="2023 -Current" | 133 |

Search Strategy C. CINAHL

Database and platform: CINAHL via EBSCOhost. Search date: December 5, 2025.

| **Line** | **Search terms** | **Results** |
| --- | --- | --- |
| 1 | (MH "Artificial Intelligence") OR (MH "Artificial Intelligence, Generative") OR (MH "Machine Learning") OR (MH "Deep Learning") OR (MH "Natural Language Processing") | 34976 |
| 2 | XB (("artificial intelligence" N5 ("research question*" or search* or criteria or select* or evaluat* or synthesiz* or writing or write* or summariz* or screen* or duplicat* or analy*))) | 1993 |
| 3 | XB ((("AI" or "GenAI" or "generative artificial intelligence" or "machine learning" or "deep learning" or "natural language processing" or "large language model*" or LLM*) N5 ("research question*" or search* or criteria or select* or evaluat* or synthesi#* or writing or write* or summari#* or screen* or duplicat* or analy*))) | 8713 |
| 4 | XB ((automat* N3 (search* or extract* or select* or synthesi#* or interpret* or screen* or duplicat* or analy*))) | 7084 |
| 5 | XB (("machine-assisted" N5 ("research question*" or search* or criteria or select* or evaluat* or synthesiz* or writing or write* or summariz* or screen* or duplicat* or analy*))) | 5 |
| 6 | XB ((chatgpt* or OpenAI or GPT3 or GPT-3 or GPT4 or GPT-4 or gemini or claude or claude2 or "claude 2" or "microsoft copilot" or DeepSeek or mistral or mixtral or "BLOOM ai" or Llama or LLama2 or "perplexity ai" or distillerAI or "rayyan AI" or "elicit AI" or "research rabbit" or "undermind AI" or "litmaps" or "consensus AI" or "open evidence" or "Nvivo AI")) | 3125 |
| 7 | S1 OR S2 OR S3 OR S4 OR S5 OR S6 | 47409 |
| 8 | (MH "meta analysis" OR MH "systematic review" OR MH "Technology, Medical/EV" OR PT "systematic review" OR PT "meta analysis" OR (((TI systematic* OR AB systematic*) N3 ((TI review* OR AB review*) OR (TI overview* OR AB overview*))) OR ((TI methodologic* OR AB methodologic*) N3 ((TI review* OR AB review*) OR (TI overview* OR AB overview*)))) OR (((TI quantitative OR AB quantitative) N3 ((TI review* OR AB review*) OR (TI overview* OR AB overview*) OR (TI synthes* OR AB synthes*))) OR ((TI research OR AB research) N3 ((TI integrati* OR AB integrati*) OR (TI overview* OR AB overview*)))) OR (((TI integrative OR AB integrative) N3 ((TI review* OR AB review*) OR (TI overview* OR AB overview*))) OR ((TI collaborative OR AB collaborative) N3 ((TI review* OR AB review*) OR (TI overview* OR AB overview*))) OR ((TI pool* OR AB pool*) N3 (TI analy* OR AB analy*))) OR ((TI "met analy*" OR AB "met analy*") OR (TI metanaly* OR AB metanaly*) OR (TI "technology assessment*" OR AB "technology assessment*") OR (TI HTA OR AB HTA) OR (TI HTAs OR AB HTAs) OR (TI "technology overview*" OR AB "technology overview*") OR (TI "technology appraisal*" OR AB "technology appraisal*")) OR (TI meta-analy* OR TI metaanaly* OR TI "systematic review*" OR TI "biomedical technology assessment*" OR TI "bio-medical technology assessment*" OR AB meta-analy* OR AB metaanaly* OR AB "systematic review*" OR AB "biomedical technology assessment*" OR AB "bio-medical technology assessment*" OR MW meta-analy* OR MW metaanaly* OR MW "systematic review*" OR MW "biomedical technology assessment*" OR MW "bio-medical technology assessment*") OR (SO Cochrane OR SO health technology assessment OR SO evidence report) OR (TI metaanaly* OR AB metaanaly*))) OR (TI "umbrella review*" OR AB "umbrella review*") OR ((TI multi* OR AB multi*) N2 (TI paramet* OR AB paramet*) N2 (TI evidence OR AB evidence) N2 (TI synthesis OR AB synthesis)) OR ((TI multiparamet* OR AB multiparamet*) N2 (TI evidence OR AB evidence) N2 (TI synthesis OR AB synthesis)) OR ((TI multi-paramet* OR AB multi-paramet*) N2 (TI evidence OR AB evidence) N2 (TI synthesis OR AB synthesis)) | 335762 |
| 9 | S7 AND S8; Limiters - Publication Date: 20230101-20260131 | 1476 |

Across the three databases, 6,616 records were retrieved before deduplication: 5,007 from MEDLINE, 133 from the Cochrane Database of Systematic Reviews, and 1,476 from CINAHL. Abbreviations and syntax: ab, abstract; adjN, terms within N words of one another in Ovid; af, all fields; hw, heading word; jw, journal word; kf, keyword heading word; kw, keyword; MH, CINAHL subject heading; mp, multipurpose field; Nn, terms within n words of one another in CINAHL; pt, publication type; ti, title; XB, CINAHL all-text field.

Table S1. Expanded Characteristics of AI-Assisted and Non-AI Systematic Reviews

| **Characteristic** | **AI-assisted reviews (n=77)** | **Non-AI reviews (n=154)** |
| --- | --- | --- |
| **Continuous characteristics** |  |  |
| Publication year, mean (SD) | 2024.3 (0.7) (n=77) | 2024.4 (0.8) (n=154) |
| Databases searched, median [IQR] | 4 [3-5] (n=76) | 4 [3-5] (n=154) |
| Records retrieved, median [IQR] | 4490 [1112-10476] (n=74) | 1204 [492-5475] (n=148) |
| Records screened, median [IQR] | 1852 [506-5975] (n=74) | 762 [287-3484] (n=147) |
| Studies included, median [IQR] | 30 [14-86] (n=74) | 16 [10-34] (n=151) |
| Review team size, median [IQR] | 6 [5-8] (n=77) | 6 [4-8] (n=154) |
| Altmetric Attention Score, median [IQR] | 6 [2.2-12.5] (n=34) | 2 [1-8] (n=56) |
| 12-month citations, median [IQR] | 5 [1.2-12] (n=74) | 3 [2-8.8] (n=134) |
| Receipt-to-publication interval, days, median [IQR] | 127.5 [70.2-204.8] (n=64) | 140 [76-228] (n=129) |
| **Categorical characteristics** |  |  |
| Journal impact tier |  |  |
| NR / not ranked | 4 (5.2%) | 4 (2.6%) |
| Q1 | 52 (67.5%) | 111 (72.1%) |
| Q2 | 21 (27.3%) | 34 (22.1%) |
| Q3 | 0 (0.0%) | 5 (3.2%) |
| Q4 | 0 (0.0%) | 0 (0.0%) |
| Funding statement reported |  |  |
| Not reported | 7 (9.1%) | 14 (9.1%) |
| Reported | 70 (90.9%) | 140 (90.9%) |
| Protocol registered |  |  |
| No | 35 (45.5%) | 56 (36.4%) |
| Yes | 42 (54.5%) | 98 (63.6%) |
| Information specialist involved |  |  |
| No | 68 (88.3%) | 129 (83.8%) |
| Yes | 9 (11.7%) | 25 (16.2%) |
| Meta-analysis |  |  |
| No | 46 (59.7%) | 68 (44.2%) |
| Yes | 31 (40.3%) | 86 (55.8%) |
| Review type |  |  |
| Network meta-analysis | 0 (0.0%) | 1 (0.6%) |
| Rapid/living/descriptive/other | 4 (5.2%) | 0 (0.0%) |
| Standard systematic review | 73 (94.8%) | 153 (99.4%) |
| Review question type |  |  |
| Diagnostic | 7 (9.1%) | 10 (6.5%) |
| Etiology/Association | 7 (9.1%) | 22 (14.3%) |
| Intervention | 15 (19.5%) | 68 (44.2%) |
| Methodological | 20 (26.0%) | 7 (4.5%) |
| Other/Mixed | 11 (14.3%) | 14 (9.1%) |
| Prevalence | 6 (7.8%) | 11 (7.1%) |
| Prognostic | 5 (6.5%) | 13 (8.4%) |
| Qualitative | 6 (7.8%) | 9 (5.8%) |
| Included study designs |  |  |
| Mixed RCT + observational | 34 (44.2%) | 58 (37.7%) |
| Observational only | 13 (16.9%) | 24 (15.6%) |
| Other | 18 (23.4%) | 32 (20.8%) |
| RCT only/RCT | 9 (11.7%) | 39 (25.3%) |
| Unclear/missing | 3 (3.9%) | 1 (0.6%) |
| Corresponding author country |  |  |
| Asia excluding China | 13 (16.9%) | 30 (19.5%) |
| Australia/New Zealand | 8 (10.4%) | 11 (7.1%) |
| Canada | 3 (3.9%) | 7 (4.5%) |
| China | 4 (5.2%) | 31 (20.1%) |
| Europe | 23 (29.9%) | 27 (17.5%) |
| Other | 3 (3.9%) | 17 (11.0%) |
| UK | 5 (6.5%) | 7 (4.5%) |
| USA | 18 (23.4%) | 24 (15.6%) |
| First author degree |  |  |
| MD | 15 (19.5%) | 27 (17.5%) |
| MD-PhD | 4 (5.2%) | 6 (3.9%) |
| Missing | 13 (16.9%) | 29 (18.8%) |
| Other health/science degree | 11 (14.3%) | 5 (3.2%) |
| Other/unclear | 10 (13.0%) | 33 (21.4%) |
| PhD | 24 (31.2%) | 54 (35.1%) |
| Last author degree |  |  |
| MD | 15 (19.5%) | 21 (13.6%) |
| MD-PhD | 9 (11.7%) | 16 (10.4%) |
| Missing | 6 (7.8%) | 20 (13.0%) |
| Other health/science degree | 0 (0.0%) | 8 (5.2%) |
| Other/unclear | 6 (7.8%) | 18 (11.7%) |
| PhD | 41 (53.2%) | 71 (46.1%) |
| Review group affiliation |  |  |
| No | 74 (96.1%) | 153 (99.4%) |
| Yes | 3 (3.9%) | 1 (0.6%) |
| Clinical domain |  |  |
| Medicine/clinical specialties | 57 (74.0%) | 118 (76.6%) |
| Neurology/Neuroscience | 10 (13.0%) | 23 (14.9%) |
| Oncology | 4 (5.2%) | 16 (10.4%) |
| Psychiatry/Mental health | 6 (7.8%) | 6 (3.9%) |
| Cardiology/Cardiovascular | 3 (3.9%) | 10 (6.5%) |
| Pediatrics/Neonatology | 8 (10.4%) | 9 (5.8%) |
| Infectious disease/COVID | 3 (3.9%) | 7 (4.5%) |
| Rheumatology/MSK/Pain | 2 (2.6%) | 6 (3.9%) |
| Obstetrics/Gynecology/Reproductive health | 1 (1.3%) | 8 (5.2%) |
| Renal/Nephrology | 2 (2.6%) | 5 (3.2%) |
| Gastroenterology/Hepatology | 2 (2.6%) | 2 (1.3%) |
| Pulmonology/Respiratory | 0 (0.0%) | 4 (2.6%) |
| Endocrine/Metabolic/Nutrition | 1 (1.3%) | 6 (3.9%) |
| Hematology | 1 (1.3%) | 2 (1.3%) |
| Other clinical specialty | 14 (18.2%) | 14 (9.1%) |
| Surgery/procedural/oral health | 4 (5.2%) | 15 (9.7%) |
| - Of which: Surgery/procedural specialties | 3 (3.9%) | 8 (5.2%) |
| - Of which: Dentistry/Oral health | 1 (1.3%) | 7 (4.5%) |
| Public health/health services/digital health | 11 (14.3%) | 16 (10.4%) |
| Methodological/non-clinical/unclear | 5 (6.5%) | 5 (3.2%) |

Note. Values are n (%) unless otherwise indicated. Continuous variables are summarized among reviews with available data. AI denotes artificial intelligence; IQR, interquartile range; RCT, randomized controlled trial; SD, standard deviation.Funding statement reported indicates the presence of a funding disclosure, including statements reporting no funding, rather than confirmed receipt of financial support. Indented rows are subsets of the parent category and should not be added to the total.

Table S2. Individual-Study Characteristics of the Included Systematic Reviews

| **Manuscript ID** | **Matched set ID** | **Review group** | **First author** | **Year** | **Article title** | **Journal** | **Clinical domain** | **Review question type** | **Meta-analysis** | **AI tool/category (AI-assisted only)** | **Stage of AI use (AI-assisted only)** |
| --- | --- | --- | --- | --- | --- | --- | --- | --- | --- | --- | --- |
| 1 | 1 | AI-assisted | Aceves-Martins | 2024 | What Environmental Metrics Are Used in Scientific Research to Estimate the Impact of Human Diets?. [6] | Nutrition and Dietetics | Nutrition | Methodological | No | LLM | Title/Abstract Screening |
| 2 | 2 | AI-assisted | Adu | 2024 | Psychology of nerve injury, repair, and recovery: a systematic review. [7] | Frontiers in Rehabilitation Sciences | Neurology | Prognostic | No | Screening Classifier | Title/Abstract Screening |
| 3 | 3 | AI-assisted | Ali | 2025 | Ignorance Is Not Bliss: A Systematic Review on the Impact of Ignoring UK Medical Students During Clinical Placements. [8] | Cureus | Education | Qualitative | No | LLM | Search & Synthesis |
| 4 | 4 | AI-assisted | Allen | 2024 | The Promise of Explainable AI in Digital Health for Precision Medicine: A Systematic Review. [9] | Journal of Personalized Medicine | Biomedical | Methodological | No | LLM | Search |
| 5 | 5 | AI-assisted | Alonso-Sanchez | 2024 | Aberrant brain language network in schizophrenia spectrum disorder: a systematic review of its relation to language signs beyond symptoms. [10] | Frontiers in Psychiatry | Psychiatry | Etiology | Yes | LLM | Data Extraction |
| 6 | 6 | AI-assisted | Anghelescu | 2025 | Standardized clinical assessments and advanced AI-driven instruments used to evaluate neurofunctional deficits, including within biomarker based framework, in Parkinson's disease - human intelligence made vs. AI models - systematic review. [11] | Frontiers in Medicine | Parkinson's Disease | Other | Yes | LLM | Search/retrieval, title/abstract screening, full-text screening |
| 7 | 7 | AI-assisted | Arzani | 2025 | Detection of carotid artery calcifications using artificial intelligence in dental radiographs: a systematic review and meta-analysis. [12] | BMC Medical Imaging | Carotid Artery Diseases | Diagnostic | Yes | LLM | Screening |
| 8 | 8 | AI-assisted | Beecham | 2025 | Systematic review of patient safety incident reporting practices in maternity care. [13] | BMJ Open Quality | Patient Safety | Qualitative | No | LLM | Data Extraction |
| 9 | 9 | AI-assisted | Bell-Aldeghi | 2023 | Determinants of the Cost-Effectiveness of Telemedicine: Systematic Screening and Quantitative Analysis of the Literature. [14] | Telemedicine and e-Health | Telemedicine | Intervention | Yes | Classifier | Screening |
| 10 | 10 | AI-assisted | Bennett | 2024 | Screening for breast cancer: a systematic review update to inform the Canadian Task Force on Preventive Health Care guideline. [15] | BMC Systematic Reviews | Breast Cancer | Diagnostic | No | LLM | Screening |
| 11 | 11 | AI-assisted | Bourke | 2025 | Children's and Adolescents' Actual Motor Competence, Perceived Physical Competence and Physical Activity: A Structural Equation Modelling Meta-Analysis. [16] | Sports Medicine | Child Exercise | Other | Yes | Screening Classifier | Title/abstract screening |
| 12 | 12 | AI-assisted | Bourke | 2023 | Adherence to the World Health Organization's physical activity recommendation in preschool-aged children: a systematic review and meta-analysis of accelerometer studies. [17] | International Journal of Behavioral Nutrition and Physical Activity | Child Exercise | Prevalence | Yes | Screening Classifier | Title/abstract screening |
| 13 | 13 | AI-assisted | Brozek | 2024 | Patients' values and preferences for health states in allergic rhinitis-An artificial intelligence supported systematic review. [18] | Allergy | Allergy | Other | No | LLM | Extraction |
| 14 | 14 | AI-assisted | Burger | 2024 | Sleep disturbances in anorexia nervosa. [19] | European Eating Disorders Review | Psychiatry | Prognostic | Yes | LLM | Title/Abstract Screening |
| 15 | 15 | AI-assisted | Cale | 2025 | Analyzing Molecular Determinants of Nanodrugs' Cytotoxic Effects. [20] | International Journal of Molecular Sciences | Toxicology | Etiology | No | LLM | Search, Screening, Extraction |
| 16 | 16 | AI-assisted | Canfell | 2024 | The Impact of Digital Hospitals on Patient and Clinician Experience: Systematic Review and Qualitative Evidence Synthesis. [21] | Journal of Medical Internet research | Digital Health | Qualitative | Yes | LLM | Synthesis |
| 17 | 17 | AI-assisted | De Silva | 2024 | Development of a pharmaceutical science systematic review process using a semi-automated machine learning tool: Intravenous drug compatibility in the neonatal intensive care setting. [22] | Pharmacology Research & Perspectives | Neonatology / Pharmacy | Methodological | No | Screening Classifier | Title/abstract screening |
| 18 | 18 | AI-assisted | Deering | 2025 | Artificial Intelligence Versus Human Systematic Literature Review Into Negative-pressure Wound Therapy in Plastic Surgery. [23] | Plastic and Reconstructive Surgery - Global Open | Surgery / wound care | Methodological | No | LLM | Full-text screening, Data extraction, and GRADE Assessment |
| 19 | 19 | AI-assisted | Demirtas Yilmaz | 2025 | Prediction of Auditory Performance in Cochlear Implants Using Machine Learning Methods: A Systematic Review [24] | Audiology Research | Cochlear Implantation | Methodological | No | Screening Classifier | Title/Abstract Screening; Study Selection/Classification |
| 20 | 20 | AI-assisted | Ducreux | 2025 | Systematic review on the DNA methylation role in endometriosis: current evidence and perspectives. [25] | Clinical Epigenetics | Reproductive Medicine/Epigenetics | Etiology | No | Screening Classifier | Title and abstract screening |
| 21 | 21 | AI-assisted | Fajkis-Zajaczkowska | 2025 | Comprehensive analysis of frameworks for evaluating artificial intelligence solutions in Healthcare: A descriptive review. [26] | Computers in Biology and Medicine | Artificial intelligence / delivery of health care | Methodological | No | LLM | Synthesis (defining criteria and framework) |
| 22 | 22 | AI-assisted | Feil | 2023 | The intention-behaviour gap in physical activity: a systematic review and meta-analysis of the action control framework. [27] | British Journal of Sports Medicine | Exercise / Health Behavior | Prevalence | Yes | Screening Classifier | Title/Abstract Screening |
| 23 | 23 | AI-assisted | Feng | 2025 | Efficacy and safety of adenosine for supraventricular tachycardia: A meta-analysis utilizing BioMedGPT-LM-7B. [28] | BMC Cardiovascular Disorders | Cardiology/Tachycardia/Anti-Arrhythmia Agents | Intervention | Yes | LLM | Search Strategy generation and retrieval, risk of bias assessment, comparison of performance with other LLMs |
| 24 | 24 | AI-assisted | Franzoi | 2023 | Which individual, social, and urban factors in early childhood predict psychopathology in later childhood, adolescence and young adulthood? A systematic review [29] | SSM - Population Health | Child Development; Mental Health | Etiology | No | Screening Classifier | Title/Abstract Screening |
| 25 | 25 | AI-assisted | Gabriel | 2023 | Mobile and wearable technologies for the analysis of Ten Meter Walk Test: A concise systematic review. [30] | Helyon | Gait | Methodological | Yes | LLM | Title/Abstract Screening |
| 26 | 26 | AI-assisted | Garcia Valencia | 2025 | Health insurance and kidney transplantation outcomes in the United States: a systematic review and AI-driven analysis of disparities in access and survival [31] | Renal Failure | Kidney Transplantation | Etiology | No | LLM | Policy synthesis and recommendation generation |
| 27 | 27 | AI-assisted | Garcia-Torres | 2024 | Enhancing Clinical Reasoning with Virtual Patients: A Hybrid Systematic Review Combining Human Reviewers and ChatGPT [32] | Healthcare | Education, Medical | Intervention | No | LLM | Data Extraction |
| 28 | 28 | AI-assisted | Garg | 2023 | Exploring the role of ChatGPT in patient care (diagnosis and treatment) and medical research: A systematic review [33] | Health Promotion Perspectives | Biomedical Research | Other | No | LLM | Data Extraction; Data Analysis; Writing |
| 29 | 29 | AI-assisted | Holland | 2025 | Validity of using a semi-automated screening tool in a systematic review assessing non-specific effects of respiratory vaccines [34] | BMC Medical Research Methodology | Respiratory Tract Infections; Vaccines | Methodological | No | Screening Classifier | Title/Abstract Screening |
| 30 | 30 | AI-assisted | Hsieh | 2024 | Diagnosis and Treatment of Tethered Spinal Cord [35] | Pediatrics | Neural Tube Defects; Spinal Cord Diseases | Intervention | No | Screening Classifier | Title/Abstract Screening |
| 31 | 31 | AI-assisted | Iaconisi | 2024 | Targeting mitochondria in Cancer therapy: Machine learning analysis of hyaluronic acid-based drug delivery systems. [36] | International Journal of Biological Macromolecules | Cancer / Neoplasms | Other | No | Screening Classifier | Title Abstract Screening, Full -text screening, synthesis/meta-analysis |
| 32 | 32 | AI-assisted | Isern-de-Val | 2025 | Analysis of clinical experience with angiotensin II: A meta-analysis and 4 AI [37] | Medicina Intensiva | Shock; Angiotensin II; Vasoconstrictor Agents | Intervention | Yes | LLM | Synthesis/Meta-Analysis |
| 33 | 33 | AI-assisted | Kelani | 2024 | Early antiplatelet therapy after intravenous thrombolysis for acute ischemic stroke: a systematic review and meta-analysis. [38] | Neurological Sciences | Neurology | Intervention | Yes | Screening Classifier | Title/abstract screening |
| 34 | 34 | AI-assisted | Ketelaars | 2025 | Impact of parental adverse childhood experiences on children's healthcare utilisation: a systematic review. [39] | Journal of Epidemiology and Community Health (JECH | Pediatrics / Public Health | Etiology / Association | No | Screening classifier and LLM | Title/abstract screening (ASReview); Manuscript drafting/editing (ChatGPT used for "enhancing readability") |
| 35 | 35 | AI-assisted | Kumar | 2025 | Impact of social factors on outcomes following pediatric neuro-oncology surgery in the United States: a systematic review and meta-analysis. [40] | Neurosurgical Review | Pediatric Neurosurgery | Etiology / Association | Yes | LLM, Screening classifier | Search/retrieval, Title/abstract screening |
| 36 | 36 | AI-assisted | Kwarah | 2025 | Evaluating predictive performance, validity, and applicability of machine learning models for predicting HIV treatment interruption: a systematic review. [41] | BMC Global and Public Health | Infectious Disease | Prognostic | No | Screening Classifier / Systematic Review Software | Search/retrieval |
| 37 | 37 | AI-assisted | Landau | 2024 | Characterizing the untapped potential of virtual reality in plastic and reconstructive surgical training: A systematic review on skill transferability. [42] | JPRAS Open | Plastic Surgery / Reconstructive Surgery | Methodological | No | LLM | Data extraction |
| 38 | 38 | AI-assisted | Lauria | 2023 | A Machine Learning Approach for Highlighting microRNAs as Biomarkers Linked to Amyotrophic Lateral Sclerosis Diagnosis and Progression. [43] | Biomolecules | ALS | Diagnostic | No | Other | Data extraction, Synthesis |
| 39 | 39 | AI-assisted | Lee | 2024 | Using meta-analysis and CNN-NLP to review and classify the medical literature for normal tissue complication probability in head and neck cancer. [44] | Radiation Oncology | Otolaryngology / Radiotherapy | Methodological | Yes | Other | Title/abstract screening |
| 40 | 40 | AI-assisted | Li | 2023 | Artificial Intelligence-Assisted Meta-Analysis of the Frequency of ACE I/D Polymorphisms in Centenarians and Other Long-Lived Individuals. [45] | International Journal of Molecular Sciences | Genetics | Prognostic | Yes | Screening classifier | Title/abstract screening |
| 41 | 41 | AI-assisted | Liu | 2025 | Within-person associations between psychological and contextual factors and dietary lapse in adults: a machine learning-assisted systematic review and meta-analysis. [46] | Annals of Behavioral Medicine | Behavioral Medicine / Diet / Healthy eating | Etiology | Yes | Screening Classifier | Title/abstract screening |
| 42 | 42 | AI-assisted | Matsui | 2024 | Human-Comparable Sensitivity of Large Language Models in Identifying Eligible Studies Through Title and Abstract Screening: 3-Layer Strategy Using GPT-3.5 and GPT-4 for Systematic Reviews. [47] | JMIR | Psychiatry / Bipolar Disorder | Methodological | No | LLM | Title/abstract screening |
| 43 | 43 | AI-assisted | Miao | 2024 | Characterisation of digital therapeutic clinical trials: a systematic review with natural language processing. [48] | Lancet Digital Health | Digital Therapeutics | Methodological / Characterisation | No | LLM | Title/abstract screening |
| 44 | 44 | AI-assisted | Mohamed | 2025 | The use of artificial intelligence in the prevention and management of bleeding disorders: a systematic review. [49] | Frontiers in Medicine | Bleeding disorder | Intervention / Diagnostic / Prognostic / Management | No | Screening Classifier / Management Software | Search/retrieval |
| 45 | 45 | AI-assisted | Molan | 2025 | Health Literacy and Interventions on Antibiotics Use and AMR in Younger Generations in High-Income Countries-A Systematic Review. [50] | Antibiotics | Antimicrobial Resistance / Health Literacy | Qualitative | No | LLM | Title/abstract screening |
| 46 | 46 | AI-assisted | Mondal | 2024 | Deciphering seizure semiology in corpus callosum injuries: A comprehensive systematic review with machine learning insights. [51] | Clinical Neurology and Neurosurgery | Neurology | Prognostic | Yes | Screening Classifier | Synthesis |
| 47 | 47 | AI-assisted | Mushcab | 2025 | Application of Artificial Intelligence in Cardio-Oncology Imaging for Cancer Therapy-Related Cardiovascular Toxicity: Systematic Review. [52] | JMIR cancer | Oncology | Diagnostic | No | Other (Literature Discover) | Search/retrieval |
| 48 | 48 | AI-assisted | Noe-Steinmuller | 2024 | Defining suffering in pain: a systematic review on pain-related suffering using natural language processing. [53] | PAIN The Journal of the International Association for the Study of Pain (IASP) | Anesthesiology | Qualitative | No | LLM; Other (Topic modelling) | Synthesis |
| 49 | 49 | AI-assisted | Nuvoli | 2025 | Digital screen exposure in infants, children and adolescents: a systematic review of existing recommendations. [54] | Public Health in Practice | Behavioral / Pediatrics | Other | No | LLM and Screening Classifier | Search/retrieval (ChatGPT) and Title/abstract screening (Rayyan) |
| 50 | 50 | AI-assisted | Park | 2025 | Smart Home Advancements for Health Care and Beyond: Systematic Review of Two Decades of User-Centric Innovation. [55] | JMIR Medical Internet research | Health Technology | Methodological | Yes | Text analysis | Abstract analysis |
| 51 | 51 | AI-assisted | Perronne | 2025 | Algorithmic approaches in hand imaging for rheumatic musculoskeletal diseases: A systematic literature review. [56] | Seminars in Arthritis and Rheumatism | Machine learning, Arthritis | Methodological | No | Screening Classifier | Title/Abstract Screening, Full text screening |
| 52 | 52 | AI-assisted | Peterson | 2024 | ADHD Diagnosis and Treatment in Children and Adolescents [57] | Agency for Healthcare Research and Quality | Mental Health / Pediatrics | Diagnostic and Intervention | Yes | Classifier | Data extraction |
| 53 | 53 | AI-assisted | Polin | 2025 | Enhancing literature reviews through AI integration: A case study on cognitive efficiency. [58] | Acta Psychologica | Research Methodology | Methodological | No | LLM | Data extraction |
| 54 | 54 | AI-assisted | PoshtMashhadi | 2025 | The Impact of Mass Gatherings and Events on Emergency Department: A Systematic Literature Review. [59] | Journal of Emergency Medicine | Public Health | Prevelance | No | LLM | Title/Abstract screening |
| 55 | 55 | AI-assisted | Purewal | 2025 | Human versus artificial intelligence: evaluating ChatGPT's performance in conducting published systematic reviews with meta-analysis in chronic pain research. [60] | Regional Anaesthesia&Pain Medicine | Chronic Pain | Methodological | Yes | LLM | Search, screening of title and full text and data extraction |
| 56 | 56 | AI-assisted | Rakhshandehroo | 2023 | Psychopathology in female offenders of terrorism and violent extremism: a systematic review. [61] | Frontiers in Psychiatry | Psychopathology | Prevelance | No | Screening | Search, title and full text screening |
| 57 | 57 | AI-assisted | Rich | 2025 | Sex, Sexual Orientation, and Gender Identity Measurement in Health Research: A Systematic Review and Narrative Synthesis. [62] | LGBT Health | Sexual Health | Methodological | Yes | Screening | Screening |
| 58 | 58 | AI-assisted | Robinson | 2024 | A machine-learning assisted review of the use of habit formation in medication adherence interventions for long-term conditions. [63] | Health Psychology Review | Pharmacology | Methodological | No | Classifier | Title and abstract screening |
| 59 | 59 | AI-assisted | Ruksakulpiwat | 2024 | Assessing the Efficacy of ChatGPT Versus Human Researchers in Identifying Relevant on mHealth Interventions for Improving Medication Adherence in Patients With Ischemic Stroke When Conducting Systematic Reviews: Comparative Analysis. [64] | JMIR Mhealth Uhealth | Neurology / Pharmacology | Methodological | No | LLM | Title and Full text screening |
| 60 | 60 | AI-assisted | Schopow | 2023 | Applications of the Natural Language Processing Tool ChatGPT in Clinical Practice: Comparative Study and Augmented Systematic Review. [65] | JMIR Med Inform | Clinical Practice | Intervention | No | LLM | Title and Full text screening, data extraction, drafting |
| 61 | 61 | AI-assisted | Shaffer | 2025 | Lead exposure and antisocial behavior: A systematic review of human and animal evidence. [66] | Environment Journal | Behavioural | Prevelance | No | Screening | Title and abstract screening |
| 62 | 62 | AI-assisted | Srinivasan | 2025 | Large Language Model Analysis of Reporting Quality of Randomized Clinical Trial Articles: A Systematic Review. [67] | JAMA Newtw Open | Research Methodology | Intervention | Yes | LLM | Analysis, screening |
| 63 | 63 | AI-assisted | Stafford | 2023 | Automated oxygen delivery for preterm infants with respiratory dysfunction [68] | Cochrane Databse of Systematic Reviews | Neonatology | Intervention | Yes | Screening Classifier | Title and abstract screening |
| 64 | 64 | AI-assisted | Steele | 2025 | Treatment of Obsessive-Compulsive Disorder in Children and Youth: A Meta-Analysis. [69] | Pediatrics | Pediatrics | Intervention | Yes | Screening Classifier | Title/abstract screening |
| 65 | 65 | AI-assisted | Talukdar | 2025 | Chronic Kidney Disease Prevalence in India: A Systematic Review and Meta-Analysis From Community-Based Representative Evidence Between 2011 to 2023. [70] | Nephrology | Nephrology | Prevalence | Yes | Screening Classifier | Search/retrieval |
| 66 | 66 | AI-assisted | Teo | 2024 | Federated machine learning in healthcare: A systematic review on clinical applications and technical architecture. [71] | Cell Reports Medicine | Machine Learning | Methodological | No | Screening Classifier | Title/abstract screening (Deduplication) |
| 67 | 67 | AI-assisted | Teperikidis | 2025 | Cardiac Remodeling in Preeclampsia: A Large-Language-Model-Assisted Meta-Analysis and Meta-Regression. [72] | Journal of Cardiovascular Pharmacology | Cardiovascular | Etiology | Yes | LLM | Screening, data extraction, RoB |
| 68 | 68 | AI-assisted | Trivedi | 2024 | Early versus late administration of amino acids in preterm infants receiving parenteral nutrition [73] | Cochrane Database Systematic Reviews | Neonatology | Intervention | Yes | Screening Classifier | Title/abstract screening |
| 69 | 69 | AI-assisted | Urru | 2024 | Strategies for cessation of caffeine administration in preterm infants [74] | Cochrane Database Systematic Reviews | Premature Infanat | Intervention | Yes | Screening Classifier | Title/abstract screening |
| 70 | 70 | AI-assisted | Visser | 2025 | From suspicion of cognitive decline to dementia diagnosis: a systematic review of healthcare professionals' considerations and attitudes. [75] | Age and Ageing | Neurology | Qualitative | No | Screening Classifier | Title/abstract screening |
| 71 | 71 | AI-assisted | Vizcarra | 2024 | Artificial Intelligence in the Diagnosis and Quantitative Phenotyping of Hyperkinetic Movement Disorders: A Systematic Review. [76] | Journal of Clinical Medicine | Digital Health | Diagnostic | No | LLM | Title/abstract screening |
| 72 | 72 | AI-assisted | Warren | 2023 | Functional magnetic resonance imaging, deep learning, and Alzheimer's disease: A systematic review. [77] | Journal of Neuroimaging | Neurology / Neurological disorder | Diagnostic | No | Screening Classifier | Title/abstract screening/Full text screening |
| 73 | 73 | AI-assisted | Weerakoon | 2025 | Microbiota-Based Therapies for Recurrent Clostridium difficile Infection: A Systematic Review of Their Efficacy and Safety. [78] | Cureus Journal of Medical Science | Gastroenterology | Intervention | No | Other | Search/retrieval |
| 74 | 74 | AI-assisted | Wen | 2025 | Bayesian Network Meta-Analysis of Randomized Controlled Trials Comparing the Efficacy of Diverse Interventions in Preventing Portal Venous System Thrombosis Following Splenectomy in Patients with Liver Cirrhosis. [79] | Annals of Vascular Surgery | Liver cirrhosis / thrombosis | Intervention | Yes | Screening Classifier | Title/abstract screening |
| 75 | 75 | AI-assisted | Zhu | 2025 | A systematic review and Bayesian network meta-analysis on the efficacy and potential of mobile interventions for stress management. [80] | Nature Human Behaviour | Digital health / Telemedicine / Psychology | Intervention | Yes | LLM | Title/abstract screening, data extraction |
| 76 | 76 | AI-assisted | de Zwart | 2024 | An update on tests used for intraoperative monitoring of cognition during awake craniotomy. [81] | Acta Neurochirurgica | Neurosurgery | Methodological | No | Screening Classifier | Title/abstract screening |
| 77 | 77 | AI-assisted | van Dijk | 2024 | Lack of Evidence Regarding Markers Identifying Acute Heart Failure in Patients with COPD: An AI-Supported Systematic Review. [82] | International Journal of Chronic Obstructive Pulmonary Disease | Cardiovascular | Diagnostic | No | Screening Classifier | Title/abstract screening |
| 78 | 1 | Non-AI | Riley | 2024 | Effects of saturated fatty acid consumption on lipoprotein (a): a systematic review and meta-analysis of randomized controlled trials. [83] | American Journal of Clinical Nutrition | Nutrition/CVD | Intervention | Yes |  |  |
| 79 | 1 | Non-AI | Bakhshimoghaddam | 2024 | Dietary inflammatory index and its association with risk of metabolic syndrome and its components: a systematic review and Meta-analysis of Observational studies. [84] | Journal of Health, Population and Nutrition | Nutrition / dietary inflammatory index and metabolic syndrome | Etiology | Yes |  |  |
| 80 | 2 | Non-AI | Khawaja | 2025 | Charting the Neurotropism of Human Parvovirus B19 and Encephalitis: A Systematic Review of Existing Literature of 25 Years. [85] | Cureus Journal of Medical Science | Neurology | Etiology | No |  |  |
| 81 | 2 | Non-AI | Alizadeh | 2024 | Human studies of the efficacy and safety of stem cells in the treatment of diabetic peripheral neuropathy: a systematic review and meta-analysis. [86] | Stem Cell Research & Therapy | Diabetic neuropathy | Intervention | Yes |  |  |
| 82 | 3 | Non-AI | Ngo | 2025 | A systematic review identifying effective teaching methods and their combinations for increasing empathy in physicians: pairwise and network meta-analysis. [87] | BMC Med Educ | Medical education | Intervention | Yes |  |  |
| 83 | 3 | Non-AI | Ramah | 2025 | High-Fidelity Simulators in Undergraduate Medical Education: A Systematic Review. [88] | Cureus | Medical education | Intervention | No |  |  |
| 84 | 4 | Non-AI | Akdemir | 2024 | Guided versus non-guided digital psychological interventions for cancer patients: A systematic review and meta-analysis of engagement and efficacy. [89] | Psycho-Oncology | Neoplasms/psychology | Intervention | Yes |  |  |
| 85 | 4 | Non-AI | Mestre-Ferrandiz | 2024 | Expert-based collaborative analysis of the situation and prospects of biomarker test implementation in oncology in Spain. [90] | Clinical and Translational Oncology | Oncology / Precision Medicine / Biomarkers | Other | No |  |  |
| 86 | 5 | Non-AI | Arnautovska | 2024 | Efficacy of User Self-Led and Human-Supported Digital Health Interventions for People With Schizophrenia: A Systematic Review and Meta-Analysis. [91] | Schizophrenia Bulletin | Psychiatry | Intervention | Yes |  |  |
| 87 | 5 | Non-AI | Mitra | 2024 | "My sister wants to kill me": A case report and systematic review of literature of co-occurring multiple sclerosis and psychosis. [92] | Indian Journal of Psychiatry | Multiple Sclerosis; Psychotic Disorders (no MeSH indexed in PubMed record PMID 39100118; author keywords: hallucinations, multiple sclerosis, psychosis, schizophrenia) | Intervention | No |  |  |
| 88 | 6 | Non-AI | Contreras-Ruston | 2025 | Sensory Feedback in Parkinson Disease Voice Production: A Systematic Review. [93] | Journal of Voice | Neurology | Other | No |  |  |
| 89 | 6 | Non-AI | Triolo | 2025 | Use of Smartphones and Wrist-Worn Devices for Motor Symptoms in Parkinson's Disease: A Systematic Review. [94] | Sensors | Parkinson's | Intervention | No |  |  |
| 90 | 7 | Non-AI | Adoberg | 2025 | Vascular calcification inhibitors in chronic kidney disease. [95] | Clinica Chimica Acta | Chronic kidney disease | Etiology | No |  |  |
| 91 | 7 | Non-AI | Kyriakoulis | 2025 | CCTA and CACS for preoperative cardiovascular risk stratification in patients undergoing noncardiac surgery: A systematic review and meta-analysis. [96] | Journal of Cardiovascular Computed Tomography | Coronary Angiography, Vascular Calcification, Coronary Artery Disease, Computed Tomography Angiography, Surgical Procedures, Operative, Coronary Stenosis | Prognostic | Yes |  |  |
| 92 | 8 | Non-AI | Alexandrou | 2025 | Enhancing Patient Safety in Refractory Ventricular Fibrillation: A Systematic Review of Double Sequential and Vector Change Defibrillation Barriers. [97] | Healthcare | Emergency Medicine/ Cardiology/ Resuscitation | Intervention/Implementation Barrier | Yes |  |  |
| 93 | 8 | Non-AI | Zhang | 2025 | Effectiveness of Interventions for Reducing Interruptions During Medication Administration: A Systematic Review. [98] | Worldviews on Evidence-Based Nursing | Nursing/Medication | Intervention | No |  |  |
| 94 | 9 | Non-AI | Zhang | 2024 | Effect of internet-based pulmonary rehabilitation on physical capacity and health-related life quality in patients with chronic obstructive pulmonary disease-a systematic review and meta-analysis. [99] | Disability and Rehabilitation | Pulmonary Disease, Chronic Obstructive; Quality of Life | Intervention | Yes |  |  |
| 95 | 9 | Non-AI | Sangalli | 2023 | Effectiveness of dental monitoring system in orthodontics: A systematic review. [100] | Jourmal of Orthodontics | Orthodontics | Intervention/Methodological | No |  |  |
| 96 | 10 | Non-AI | MacLehose | 2025 | CYP2D6 Phenotype and Breast Cancer Outcomes: A Bias Analysis and Meta-Analysis. [101] | Cancer Epidemiology, Biomarkers & Prevention | Breast cancer | Prognostic | Yes |  |  |
| 97 | 10 | Non-AI | Zhang | 2024 | Comparative efficacy and safety of eribulin versus paclitaxel in breast cancer: a systematic review and meta-analysis. [102] | Future Oncology | Breast cancer | Intervention | Yes |  |  |
| 98 | 11 | Non-AI | Lee | 2025 | The Effectiveness and Safety of Tai Chi on Knee Pain: A Systematic Review and Meta-Analysis. [103] | Healthcare | Knee pain | Intervention | Yes |  |  |
| 99 | 11 | Non-AI | Rostami Haji Abadi | 2025 | Psychometric Properties of Physical Activity Questionnaires for Children and Adolescents: An Updated Systematic Review. [104] | Sports Medicine | Sports Medicine | Methodological | No |  |  |
| 100 | 12 | Non-AI | Larouche | 2023 | Determinants of Outdoor Time in Children and Youth: A Systematic Review of Longitudinal and Intervention Studies. [105] | International Journal of Environmental Research and Public Health | COVID-19, Child, Adolescent, Exercise | Qualitative | No |  |  |
| 101 | 12 | Non-AI | Weng | 2023 | Comparative efficacy of exercise therapy and oral non-steroidal anti-inflammatory drugs and paracetamol for knee or hip osteoarthritis: a network meta-analysis of randomised controlled trials. [106] | British Journal of Sports Medicine | Orthopaedics / Rheumatology | Intervention | Yes |  |  |
| 102 | 13 | Non-AI | He | 2024 | Meta-analysis of randomized controlled trials assessing the efficacy of nigella sativa supplementation for allergic rhinitis treatment. [107] | Frontiers in Pharmacology | Ethnopharmacology | Intervention | Yes |  |  |
| 103 | 13 | Non-AI | Shao | 2024 | Efficacy and safety of montelukast-levocetirizine combination therapy in combined allergic rhinitis and asthma syndrome: a systematic review and meta-analysis. [108] | Journal of Asthma | Allergic rhinitis | Intervention | Yes |  |  |
| 104 | 14 | Non-AI | Holanda da Cunha | 2025 | Auriculotherapy for people with sleep disorders: A systematic review. [109] | Complementary Therapies in Clinical Practice | Auriculotherapy; Sleep initiation and maintenance | Intervention | No |  |  |
| 105 | 14 | Non-AI | Cassioli | 2024 | Leptin Levels in Acute and Recovered Eating Disorders: An Arm-Based Network Meta-Analysis. [110] | European Eating Disorders Review | Leptin, Feeding and Eating Disorders | Intervention | Yes |  |  |
| 106 | 15 | Non-AI | Firouzabadi | 2025 | The Impact of Cadmium Telluride Quantum Dots on Male Reproductive Health: A Systematic Review of Toxicological Effects and Mechanisms. [111] | World Journal of Men’s Health | Male reproductive health / Reproductive toxicology / Nanotoxicology | Etiology-Toxicology | No |  |  |
| 107 | 15 | Non-AI | Oyowvi | 2025 | Emerging nanotechnologies and their role in early ovarian cancer detection, diagnosis and interventions. [112] | Journal of Ovarian Research | Oncology (ovarian cancer diagnosis) / Nanomedicine | Diagnostic | No |  |  |
| 108 | 16 | Non-AI | Miake-Lye | 2023 | Transitioning from One Electronic Health Record to Another: A Systematic Review. [113] | Journal of General Internal Medicine | Electronic Health Records; Outcome Assessment, Health Care (PubMed major MeSH headings for PMID 37798580) | Other | No |  |  |
| 109 | 16 | Non-AI | Bertocchi | 2024 | Exploring the nexus between the standardized nursing terminologies and the unfinished nursing care phenomenon: An empty systematic review. [114] | International Journal of Nursing Knowledge | Nursing | Other | No |  |  |
| 110 | 17 | Non-AI | Elfios | 2024 | Parental satisfaction with neonatal intensive care unit services and associated factors in Ethiopia: systematic review and meta-analysis. [115] | BMC Nursing | Patient care | Qualitative | Yes |  |  |
| 111 | 17 | Non-AI | Shetty | 2023 | Prevalence of anxiety, depression, and stress among parents of neonates admitted to neonatal intensive care unit: a systematic review and meta-analysis. [116] | Clinical and Experimental Pediatrics | Anxiety; depression; stress; NICU | Prevalence | Yes |  |  |
| 112 | 18 | Non-AI | Karamitros | 2025 | Steal Syndrome in Free Flap Microvascular Reconstruction of the Lower Extremity: Systematic Review of Incidence, Risk Factors, and Surgical Management. [117] | Bioengineering | Reconstructive surgery | Etiology | No |  |  |
| 113 | 18 | Non-AI | Acosta | 2025 | Effectiveness of Negative Pressure Wound Therapy With Instillation and Dwell in Removing Nonviable Tissue, Promoting Granulation Tissue, and Reducing Surgical Debridements: A Systematic Literature Review. [118] | Wound Repair and Regeneration | Wound care / wound therapy | Intervention | No |  |  |
| 114 | 19 | Non-AI | Nair | 2025 | A systematic review of machine learning approaches in cochlear implant outcomes. [119] | NPJ Digital Medicine | Otology | Methodological | No |  |  |
| 115 | 19 | Non-AI | Padurean | 2025 | Music Therapy Outcomes in Older Adults Using Cochlear Implants, Hearing Aids, or Combined Bimodal Devices: A Systematic Review. [120] | Healthcare | Music therapy | Intervention | No |  |  |
| 116 | 20 | Non-AI | de Souza Gaio | 2025 | Clinical effectiveness of progestogens compared to combined oral contraceptive pills in the treatment of endometriosis: A systematic review and meta-analysis. [121] | European journal of obstetrics & gynecology and reproductive biology | Endometriosis; gynecology | Intervention | Yes |  |  |
| 117 | 20 | Non-AI | Maguire | 2025 | A quality-of-life meta-analysis comparing pre- and postoperative symptoms in women undergoing colorectal resection for deep infiltrating endometriosis. [122] | Colorectal Disease | Endometriosis, Colectomy, Colonic Diseases, Rectal Diseases, Quality of Life | Prognostic | Yes |  |  |
| 118 | 21 | Non-AI | Chiang | 2025 | How accurate is AI in detecting marginal jaw bone loss? A systematic review and meta-analysis. [123] | Journal of Dentistry | Dentistry / Periodontology / Dental radiology / Artificial intelligence | Diagnostic | Yes |  |  |
| 119 | 21 | Non-AI | Stockdale | 2025 | Service delivery models and care cascade outcomes for people living with chronic hepatitis B: a global systematic review and meta-analysis. [124] | The Lancet Gastroenterology and Hepatology | Hepatology | Intervention | Yes |  |  |
| 120 | 22 | Non-AI | Guimaraes | 2023 | Water-Based Exercises on Peak Oxygen Consumption, Exercise Time, and Muscle Strength in Patients with Coronary Artery Disease: A Systematic Review with Meta-Analysis. [125] | Cardiovascular therapeutics | Coronary Artery Disease | Intervention | Yes |  |  |
| 121 | 22 | Non-AI | Wang | 2023 | Effectiveness of different exercises in improving postural balance among Parkinson's disease patients: a systematic review and network meta-analysis. [126] | Frontiers in Aging Neuroscience | Exercise intervention | Intervention | Yes |  |  |
| 122 | 23 | Non-AI | Meszaros | 2025 | Decreased peak atrial longitudinal strain and left atrial contraction strain rate values may predict atrial fibrillation in patients in sinus rhythm at baseline: a systematic review and meta-analysis. [127] | European Heart Journal Open | Cardiology / cardiac imaging | Prognostic | Yes |  |  |
| 123 | 23 | Non-AI | Chen | 2025 | Does atrial fibrillation increase the risk of fractures? A systematic review and meta-analysis. [128] | Frontiers in Medicine | Bone fracture, arrhythmia | Etiology | Yes |  |  |
| 124 | 24 | Non-AI | Bali | 2024 | Exploring data sources and mathematical approaches for estimating human mobility rates and implications for understanding COVID-19 dynamics: a systematic literature review. [129] | Journal of Mathematical Biology | COVID-19 | Methodological | No |  |  |
| 125 | 24 | Non-AI | Sutarsa | 2024 | Multisectoral interventions and health system performance: a systematic review. [130] | Bull World Health Organ. | Healthcare services & delivery | Intervention | No |  |  |
| 126 | 25 | Non-AI | Diaz | 2023 | Mining Sensor Data to Assess Changes in Physical Activity Behaviors in Health Interventions: Systematic Review. [131] | JMIR Medical Informatics | Digital Health | Methodological | No |  |  |
| 127 | 25 | Non-AI | Seffah | 2023 | Exploring the Role of Wearable Electronic Medical Devices in Improving Cardiovascular Risk Factors and Outcomes Among Adults: A Systematic Review. [132] | Cureus Journal of Medical Science | Cardiovascular | Intervention | No |  |  |
| 128 | 26 | Non-AI | Muxunov | 2026 | Health-related quality of life in chronic kidney disease patients in low- and lower-middle income countries: a systematic review and meta-analysis. [133] | Quality of Life Research | Nephrology (CKD) | Prevalence | Yes |  |  |
| 129 | 26 | Non-AI | Chen | 2025 | Comparison of intraoperative and postoperative outcomes between open and robotic-assisted kidney transplantation: a meta-analysis and systematic review. [134] | Journal of Robotic Surgery | Robotic-assisted / Transplantation | Intervention | Yes |  |  |
| 130 | 27 | Non-AI | Hou | 2025 | The impact of virtual reality applied in nurse education on students' clinical thinking: A systematic review. [135] | Nurse Education Today | Nurse education; virtual reality | Intervention | No |  |  |
| 131 | 27 | Non-AI | Rohani | 2024 | Identifying Learning Preferences and Strategies in Health Data Science Courses: Systematic Review. [136] | JMIR Medical Education | Learning | Qualitative | No |  |  |
| 132 | 28 | Non-AI | Casey | 2023 | Application of a Comprehensive Evaluation Framework to COVID-19 Studies: Systematic Review of Translational Aspects of Artificial Intelligence in Health Care. [137] | JMIR | Coronavirus | Methodological | No |  |  |
| 133 | 28 | Non-AI | Ahmed | 2023 | An appraisal of the performance of AI tools for chronic stroke lesion segmentation. [138] | Computers in Biology and Medicine | Neuroradiology | Diagnostic | No |  |  |
| 134 | 29 | Non-AI | Li | 2024 | Willingness to pay for vaccines in China: A systematic review and single-arm Bayesian analysis. [139] | Human Vaccines & Immunotherapeutics | Vaccination | Prevelance | Yes |  |  |
| 135 | 29 | Non-AI | Wang | 2025 | Effectiveness of nirsevimab immunization against RSV infection in preterm infants: a systematic review and meta-analysis. [140] | Frontiers in Immunology | Neonatology; infectious diseases | Intervention | Yes |  |  |
| 136 | 30 | Non-AI | Gandy | 2024 | Cognitive behavioral interventions for depression and anxiety in adults with neurological disorders: a systematic review and meta-analysis. [141] | Psychological Medicine | Neurology | Intervention | Yes |  |  |
| 137 | 30 | Non-AI | Felisbino | 2025 | A Systematic Review of Parental Occupational Pesticide Exposure and Geographical Proximity to Agricultural Fields in Association with Neural Tube Defects. [142] | Toxics | Neural Tube Defects | Etiology | No |  |  |
| 138 | 31 | Non-AI | Rehim | 2024 | Mirvetuximab Soravtansine in solid tumors: A systematic review and meta-analysis. [143] | PLOS ONE | Oncology | Intervention | Yes |  |  |
| 139 | 31 | Non-AI | Wang | 2024 | Traditional Chinese Medicine in post-resection liver cancer therapy: a meta-analysis of efficacy. [144] | Am J Transl Res | Oncology | Intervention | Yes |  |  |
| 140 | 32 | Non-AI | Couppoussamy | 2026 | Effect of tranexamic acid in the treatment of angiotensin-converting enzyme inhibitor-induced angioedema: A systematic review and meta-analysis. [145] | Turkish Journal of Emergency Medicine | Intensive care | Intervention | Yes |  |  |
| 141 | 32 | Non-AI | Tejada | 2025 | Continuous versus intermittent infusion of beta-lactams in patients with sepsis and septic shock: a systematic review and meta-analysis. [146] | BMC Infectious Diseases | Infectious Disease | Intervention | Yes |  |  |
| 142 | 33 | Non-AI | Geravandi | 2024 | A comparison of the effects of ticagrelor and clopidogrel in patients with acute ST-segment elevation myocardial infarction: a systematic review and meta-analysis of randomized clinical trials. [147] | BMC Pharmacology and Toxicology | Cardiology | Intervention | Yes |  |  |
| 143 | 33 | Non-AI | Ghozy | 2025 | Leukoaraiosis severity and outcomes of endovascular thrombectomy for acute ischemic stroke: a systematic review and meta-analysis. [148] | European Radiology | Thrombectomy; Ischemic Stroke; Endovascular Procedures; Leukoaraiosis (major MeSH headings per PubMed PMID 40358674) | Etiology | Yes |  |  |
| 144 | 34 | Non-AI | Karimi | 2025 | The association of direct-acting antiviral therapy with health care utilization and costs in patients with chronic hepatitis C virus infection: A systematic review of real-world studies in the United States. [149] | Journal of Managed Care & Specialty Pharmacy | Gastroenterology | Intervention | No |  |  |
| 145 | 34 | Non-AI | Kumar | 2025 | Social Determinants of Human Papillomavirus Vaccine Uptake Among Adolescent Girls in Low-Middle-Income Countries: A Systematic Review. [150] | INQUIRY: The Journal of Health Care Organization, Provision, and Financing | Pulpotomy | Prevelance | Yes |  |  |
| 146 | 35 | Non-AI | Jayawardane | 2025 | Long-term morbidity of peripartum hysterectomy: A systematic review. [151] | International Journal of Gynecology & Obstetrics | Gynecology | Prognostic | No |  |  |
| 147 | 35 | Non-AI | Topala | 2025 | To cut or not to cut? Extended mesenteric excision during intestinal resection does not impact the postoperative recurrence nor the postoperative complications in Crohn's disease: a systematic review and meta-analysis. [152] | Techniques in Coloproctology | Gastroenterology / Colorectal Surgery / Inflammatory Bowel Disease / Crohn’s Disease | Intervention | Yes |  |  |
| 148 | 36 | Non-AI | Mukoka | 2025 | The key design features and effectiveness of social network interventions for HIV testing and linkage services in low- and middle-income countries: a systematic review and meta-analysis. [153] | Journal of the International AIDS Society | HIV / Infectious diseases | Intervention | Yes |  |  |
| 149 | 36 | Non-AI | Strauss | 2025 | HIV Infection and Antiretroviral Therapy Impair Liver Function in People Living with HIV: Systematic Review and Meta-Analysis. [154] | Pharmaceuticals | HIV/Hepatology | Etiology | Yes |  |  |
| 150 | 37 | Non-AI | Goh | 2025 | Effectiveness of Digital-Based Interventions on Physical and Psychological Outcomes Among Cancer Patients: A Systematic Review and Meta-Analysis. [155] | Seminars in Oncology Nursing | Oncology | Intervention | Yes |  |  |
| 151 | 37 | Non-AI | Valentine | 2024 | Impact of Medical Trainees on Efficiency and Productivity in the Emergency Department: Systematic Review. [156] | Western Journal of Emergency Medicine | Emergency Medicine | Intervention | No |  |  |
| 152 | 38 | Non-AI | Hong | 2023 | Meta analysis of the diagnostic value of circulating miRNA in benign and malignant pulmonary nodules. [157] | World Journal of Surgical Oncology | Oncology | Diagnostic | Yes |  |  |
| 153 | 38 | Non-AI | Lee | 2024 | Epigenetic MicroRNAs as Prognostic Markers of Postoperative Atrial Fibrillation: A Systematic Review. [158] | Current Problems in Cardiology | Atrial Fibrillation, Cardiac Surgical Procedures, MicroRNAs | Prognostic | No |  |  |
| 154 | 39 | Non-AI | Fonseca | 2024 | Global prevalence of human papillomavirus-related oral and oropharyngeal squamous cell carcinomas: a systematic review and meta-analysis. [159] | Clinical Oral Investigations | Dentistry / Oropharyngeal Cancer / HPV-related cancer | Prevalence | Yes |  |  |
| 155 | 39 | Non-AI | Lechien | 2023 | Epidemiological, clinical and oncological outcomes of laryngeal verrucous carcinomas: a systematic review. [160] | Journal of Otolaryngology - Head & Neck Surgery | Oncology / otolaryngology | Prognostic | No |  |  |
| 156 | 40 | Non-AI | Qu | 2023 | The Effects of Light Therapy for Depression in Dementia: A Systematic Review and Meta-Analysis. [161] | Journal of Alzheimer's Disease | Alzheimer's | Intervention | Yes |  |  |
| 157 | 40 | Non-AI | Lu | 2023 | The forkhead box protein P3 gene rs3761548 promoter polymorphism confers a genetic contribution to the risk of preeclampsia: A systematic review and meta-analysis. [162] | Cytokine | Pre-Eclampsia; Polymorphism, Single Nucleotide (major MeSH headings, PubMed PMID 36842371) | Etiology | Yes |  |  |
| 158 | 41 | Non-AI | Rostampour | 2025 | Dietary Flavonoids and Lung Cancer: A GRADE-Assessed Systematic Review and Meta-Analysis of Observational Studies. [163] | Nutrition and Cancer | Diet; lung neoplasm | Etiology | Yes |  |  |
| 159 | 41 | Non-AI | Flores-Hernández | 2024 | Efficacy of a High-Protein Diet to Lower Glycemic Levels in Type 2 Diabetes Mellitus: A Systematic Review. [164] | International Journal of Molecular Sciences | Endocrinology | Intervention | No |  |  |
| 160 | 42 | Non-AI | Wen | 2024 | Dimensional Neuroimaging Endophenotypes: Neurobiological Representations of Disease Heterogeneity Through Machine Learning. [165] | Biological Psychiatry | Psychiatry / Neurology / Neuroimaging / Machine Learning | Methodological | No |  |  |
| 161 | 42 | Non-AI | Khalid | 2025 | An Appraisal of the Quality of Development and Reporting of Predictive Models in Neurosurgery: A Systematic Review. [166] | Neurosurgery | Neurosurgery | Methodological | No |  |  |
| 162 | 43 | Non-AI | Gilbert | 2024 | The use of master protocols for efficient trial design to evaluate radiotherapy interventions: a systematic review. [167] | J Natl Cancer Inst | Neoplasms (radiotherapy); Research Design | Intervention | No |  |  |
| 163 | 43 | Non-AI | Dadon | 2024 | Vitamin D and Insulin-Dependent Diabetes: A Systematic Review of Clinical Trials. [168] | nutrients | Endocrinology | Intervention | No |  |  |
| 164 | 44 | Non-AI | Dumbuya | 2025 | Effectiveness of health-related quality of life assessment scales in patients with haemophilia after treatment: Systematic review. [169] | Thrombosis Research | Hematology | Qualitative | No |  |  |
| 165 | 44 | Non-AI | Han | 2025 | Heparin-Induced Thrombocytopenia and Thrombosis in Patients With or Without a Thrombophilia Background: A Systematic Review Involved 602 Cases. [170] | International Journal of Vascular Medicine | Hematology | Prognostic | No |  |  |
| 166 | 45 | Non-AI | Wan Mohd Johari | 2025 | Instruments Assessing Medication Literacy in Psychiatric Patients and the Caregivers: A Systematic Review. [171] | Pharmacopsychiatry | Psychiatry / mental health | Methodological / Other | No |  |  |
| 167 | 45 | Non-AI | Caut | 2025 | Health professionals' beliefs and attitudes towards preconception care: a systematic review. [172] | BMC Health Services Research | Health Services Research / Preconception Care / Reproductive Health / Primary Care | Other | No |  |  |
| 168 | 46 | Non-AI | Abdallah | 2024 | Systematic review of seizure-onset patterns in stereo-electroencephalography: Current state and future directions. [173] | Clinical Neurophysiology | Electroencephalography; Seizures (PubMed major MeSH: Electroencephalography, Seizures) | Other | Yes |  |  |
| 169 | 46 | Non-AI | Hwang | 2024 | Assistive technology on upper extremity function for stroke patients: A systematic review with meta-analysis. [174] | Journal of Hand Therapy | Neurology / Rehabilitation | Intervention | Yes |  |  |
| 170 | 47 | Non-AI | Hanafi | 2025 | Prediction of Emerging CVD Biomarkers in Population Exposed to Arsenic and Their Associated Risk Factors in Humans by Systematic Review and Meta-Analysis. [175] | Biological Trace Element Research | Cardiovascular diseases | Prevelance | Yes |  |  |
| 171 | 47 | Non-AI | Meillan | 2025 | Early Breast Cancer Treatment and Cardiac Events: A Systematic Review. [176] | Clinical Breast Cancer | Cardiology; breast cancer | Prevalence | No |  |  |
| 172 | 48 | Non-AI | Liu | 2024 | The effects of foot orthoses on radiological parameters and pain in children with flexible flat feet: a systematic review and meta-analysis. [177] | Frontiers in Pediatrics | Foot care | Intervention | Yes |  |  |
| 173 | 48 | Non-AI | Altuhafy | 2024 | Efficacy of computer-controlled local anesthesia delivery system on pain in dental anesthesia: a systematic review of randomized clinical trials. [178] | Journal of Dental Anesthesia and Pain Medicine | Dentistry / dental local anesthesia pain control | Intervention | No |  |  |
| 174 | 49 | Non-AI | Lima | 2025 | Impact of screen use on behavior and sleep in patients with autism spectrum disorder. [179] | Arquivos de Neuro-Psiquiatria | Autsism Spectrum Disorder | Etiology | No |  |  |
| 175 | 49 | Non-AI | Zgambo | 2025 | Effect of Digital Safety Interventions on Parental Practices in Safeguarding Children's Digital Activities: Systematic Review and Meta-Analysis. [180] | JMIR Pediatrics and Parenting | Pediatrics | Intervention | Yes |  |  |
| 176 | 50 | Non-AI | Clack | 2025 | Defining and Measuring Organizational Transformation in Health Care: A Systematic Literature Review. [181] | Medical Care Research and Review | Quality Improvement | Qualitative | No |  |  |
| 177 | 50 | Non-AI | Seddik | 2025 | A Comprehensive View of the Methods Used to Measure the Societal Impact of Healthcare Interventions: A Systematic Review. [182] | Value in Health | Delivery of healthcare | Qualitative | No |  |  |
| 178 | 51 | Non-AI | Xie | 2026 | Difficult-to-treat rheumatoid arthritis: systematic review and meta-analysis on global prevalence. [183] | Annals of the Rheumatic Diseases | Rheumatology | Prevalence | Yes |  |  |
| 179 | 51 | Non-AI | Liu | 2025 | Exposure to Occupational Inhalants and the Risk of Developing Rheumatoid Arthritis: A Systematic Review and Meta-analysis. [184] | Arthritis and Rheumatology | Rheumatology | Etiology | Yes |  |  |
| 180 | 52 | Non-AI | Long | 2024 | Shared and Distinct Neurobiological Bases of Bipolar Disorder and Attention-Deficit/Hyperactivity Disorder in Children and Adolescents: A Comparative Meta-Analysis of Structural Abnormalities. [185] | Journal of the American Academy of Child & Adolescent Psychiatry | Psychiatry / Neurology | Etiology | Yes |  |  |
| 181 | 52 | Non-AI | Frisira | 2025 | Systematic review and meta-analysis: relative age in attention-deficit/ hyperactivity disorder and autism spectrum disorder. [186] | European Child & Adolescent Psychiatry | Attention Deficit Disorder with Hyperactivity; Autism Spectrum Disorder | Etiology | Yes |  |  |
| 182 | 53 | Non-AI | Dejakaisaya | 2025 | Television watching and cognitive outcomes in adults and older adults: A systematic review and dose-response meta-analysis of observational studies. [187] | PLOS One | Cognition | Prognostic | Yes |  |  |
| 183 | 53 | Non-AI | Carollo | 2025 | Early Development in cross-country perspective: A systematic review of a quarter century of developmental research. [188] | Infant Behavior and Development | Child Development | Prevelance | No |  |  |
| 184 | 54 | Non-AI | N Alaqil | 2025 | The Role of Urgent Care Clinics in Alleviating Emergency Department Congestion: A Systematic Review of Patient Outcomes and Resource Utilization. [189] | Cureus | ED Congestion | Intervention | No |  |  |
| 185 | 54 | Non-AI | Abram | 2025 | Improving patient care experience in the pediatric emergency department: a systematic review. [190] | Canadian Journal of Emergency Medicine | Pediatric emergency medicine / patient experience | Qualtitative | No |  |  |
| 186 | 55 | Non-AI | Russo | 2025 | Systematic review and meta-analysis of conventional medical management in a patient population with refractory chronic pain suitable to receive a spinal cord stimulation system. [191] | Pain Medicine | Spinal Cord Stimulation; Chronic Pain; Pain Management; Intractable Pain | Intervention | Yes |  |  |
| 187 | 55 | Non-AI | Huygen | 2024 | Spinal Cord Stimulation vs Medical Management for Chronic Back and Leg Pain: A Systematic Review and Network Meta-Analysis. [192] | JAMA Network Open | Back pain | Intervention | Yes |  |  |
| 188 | 56 | Non-AI | Zhao | 2023 | Triage Algorithms for Mass-Casualty Bioterrorism: A Systematic Review. [193] | International Journal of Environmental and Public Health | Mass casualty incidents | Other | No |  |  |
| 189 | 56 | Non-AI | Narimani | 2025 | Comprehensive psychosocial support in CBRNE incidents: a systematic review of strategies for citizens and frontline personnel. [194] | BMC Emergency Medicine | Public Health | Other | No |  |  |
| 190 | 57 | Non-AI | Gonzalez | 2025 | Prevalence of Insomnia by Sex in U.S. Service Members: A Systematic Review and Meta-Analysis. [195] | Military Medicine | Sleep / insomnia in U.S. military service members | Prevalence | Yes |  |  |
| 191 | 57 | Non-AI | Lagden | 2025 | Chemsex Interventions for Men Who Have Sex With Men: A Systematic Review and Meta-Analysis. [196] | Cureus | Chemsex | Intervention | Yes |  |  |
| 192 | 58 | Non-AI | Collonnaz | 2024 | Lack of consistency in measurement methods and semantics used for network measures in adolescent health behaviour studies using social network analysis: a systematic review. [197] | Journal of Epidemiology and Community Health | Adolescent Behavior; Health Behavior; Peer Group | Etiology | No |  |  |
| 193 | 58 | Non-AI | Xu | 2023 | A Systematic Review of Dietary Interventions for Cancer Survivors and Their Families or Caregivers. [198] | Nutrients | Oncology | Intervention | No |  |  |
| 194 | 59 | Non-AI | Teti | 2025 | Telepractice Assessments for Individuals with Aphasia: A Systematic Review. [199] | Telemedicine and e-Health | Aphasia; telemedicine | Intervention | No |  |  |
| 195 | 59 | Non-AI | Pannunzio | 2024 | Patient and Staff Experience of Remote Patient Monitoring-What to Measure and How: Systematic Review. [200] | Journal of Medical Internet Research | Quality improvement | Qualitative | Yes |  |  |
| 196 | 60 | Non-AI | Jiao | 2024 | Systematic review of automatic post-stroke gait classification systems. [201] | Gait and Posture | Stroke/biomechanics | Methodological/ Other | No |  |  |
| 197 | 60 | Non-AI | Samartha | 2024 | AI-driven estimation of O6 methylguanine-DNA-methyltransferase (MGMT) promoter methylation in glioblastoma patients: a systematic review with bias analysis. [202] | Journal of Cancer Research and Clinical Oncology | Oncology / Neuro-oncology / Artificial intelligence | Diagnostic | No |  |  |
| 198 | 61 | Non-AI | Abdelkhader | 2025 | Partial pulpotomy for carious pulp exposure in adult mature permanent teeth: a systematic review and meta-analysis. [203] | BMC Oral Health | Pulpotomy; Dental Pulp Exposure; Dental Caries (major MeSH: Pulpotomy, Dental Caries, Dental Pulp Exposure — PubMed PMID 41339865) | Intervention | Yes |  |  |
| 199 | 61 | Non-AI | Ou | 2025 | Meta-analysis of maternal exposure to heavy metals (lead, cadmium, mercury, chromium) and adverse pregnancy outcomes. [204] | BMC Pregnancy and Childbirth | Pregnancy | Etiology | Yes |  |  |
| 200 | 62 | Non-AI | Erqou | 2025 | Re-evaluating Fasting Guidelines for Invasive Cardiac Procedures: A Systematic Review and Meta-analysis of Randomized Clinical Trials. [205] | Journal of the Society for Cardiovascular Angiography & Interventions | Cardiac Surgery, Anaesthesiology | Intervention | Yes |  |  |
| 201 | 62 | Non-AI | Soma | 2025 | Clinical and Radiographic Outcomes Following Pulpotomy Using Biodentine in Carious Exposed Mature Permanent Teeth: A Systematic Review and Meta-analysis. [206] | Journal of International Society of Preventative & Community Dentistry | Dentistry | Prognostic | Yes |  |  |
| 202 | 63 | Non-AI | Hay | 2023 | Systemic corticosteroids for the prevention of bronchopulmonary dysplasia, a network meta-analysis. [207] | Cochrane Database of Systematic Reviews | Neonatology | Intervention | Yes |  |  |
| 203 | 63 | Non-AI | Zhuang | 2024 | A Network Meta-Analysis of Success Rates following Low Dosage Anti-VEGF for Retinopathy of Prematurity. [208] | CURRENT EYE RESEARCH | Retinopathy | Intervention | Yes |  |  |
| 204 | 64 | Non-AI | Pastre | 2025 | Serotonergic underpinnings of obsessive-compulsive disorder: A systematic review and meta-analysis of neuroimaging findings. [209] | Psychiatry and Clinical Neurosciences | Obsessive-Compulsive Disorder; Serotonin Plasma Membrane Transport Proteins; Receptor, Serotonin, 5-HT2A; Serotonin; Neuroimaging; Brain (PubMed major MeSH headings, PMID 39511769) | Etiology | Yes |  |  |
| 205 | 64 | Non-AI | Pellegrini | 2025 | Meta-analysis of age at help-seeking and duration of untreated illness (DUI) in obsessive-compulsive disorder (OCD): The need for early interventions. [210] | Journal of Affective Disorders | Psychiatry | Prognostic | Yes |  |  |
| 206 | 65 | Non-AI | Chen | 2025 | Kidney and Cardiovascular Outcomes Among Patients With CKD Receiving GLP-1 Receptor Agonists: A Systematic Review and Meta-Analysis of Randomized Trials. [211] | American Journal of Kidney Diseases | Nephrology | Intervention | Yes |  |  |
| 207 | 65 | Non-AI | Korsa | 2025 | Risk Factor-Based Screening for Early Detection of Chronic Kidney Disease in Primary Care Settings: A Systematic Review. [212] | Kidney Medicine | Nephrology | Etiology | No |  |  |
| 208 | 66 | Non-AI | Pigoni | 2024 | Machine learning and the prediction of suicide in psychiatric populations: a systematic review. [213] | Translational Psychiatry | Psychiatry | Prognostic | No |  |  |
| 209 | 66 | Non-AI | Issaiy | 2023 | Artificial Intelligence and Acute Appendicitis: A Systematic Review of Diagnostic and Prognostic Models. [214] | World Journal of Emergency Surgery | AI/machine-learning models for diagnosis and prognosis | Diagnostic | No |  |  |
| 210 | 67 | Non-AI | Akankwasa | 2025 | Burden and determinants of preeclampsia and eclampsia in Ugandan tertiary healthcare facilities: a systematic review. [215] | Pregnancy Hypertension | Pre-Eclampsia; Eclampsia | Prevalence | No |  |  |
| 211 | 67 | Non-AI | Siargkas | 2025 | Uterine Artery Pulsatility Index in Singleton Pregnancies Conceived via Assisted Reproductive Technology Versus Spontaneous Conception: A Systematic Review and Meta-Analysis. [216] | Diagnostics | Pregnancy | Etiology | Yes |  |  |
| 212 | 68 | Non-AI | Seidler | 2023 | Short, medium, and long deferral of umbilical cord clamping compared with umbilical cord milking and immediate clamping at preterm birth: a systematic review and network meta-analysis with individual participant data. [217] | The Lancet | Neonatology | Intervention | Yes |  |  |
| 213 | 68 | Non-AI | Zeng | 2023 | Comparing different scoring systems for predicting mortality risk in preterm infants: a systematic review and network meta-analysis. [218] | Frontiers in Pediatrics | Neonatology | Prognostic | Yes |  |  |
| 214 | 69 | Non-AI | Poon | 2024 | Exploring the ergogenic potential of carbohydrate-caffeine combined mouth rinse on exercise and cognitive performance: a systematic review. [219] | Applied Physiology, Nutrition, and Metabolism | Sports Nutrition | Intervention | No |  |  |
| 215 | 69 | Non-AI | Hu | 2024 | Maternal smoking, consumption of alcohol, and caffeinated beverages during pregnancy and the risk of childhood brain tumors: a meta-analysis of observational studies. [220] | BMC Public Health | Cancer epidemiology | Etiology | Yes |  |  |
| 216 | 70 | Non-AI | Zhao | 2025 | The prevalence and influencing factors of reversion from mild cognitive impairment to normal cognition: A systemic review and meta-analysis. [221] | Geriatric Nursing | Geriatrics / Cognitive impairment / Alzheimer disease / Nursing | Prevalence/Prognostic Factors | Yes |  |  |
| 217 | 70 | Non-AI | Liu | 2025 | Alzheimer disease is (sometimes) highly heritable: Drivers of variation in heritability estimates for binary traits, a systematic review. [222] | PLOS Genetics | Neurology | Diagnostics | No |  |  |
| 218 | 71 | Non-AI | Davydow | 2025 | Potential Risk Factors for Psychosis in Parkinson's Disease: A Review of Cohort and Case-Control Studies. [223] | Journal of Geriatric Psychiatry and Neurology | Risk factors for psychosis in Parkinson's disease | Etiology | No |  |  |
| 219 | 71 | Non-AI | Panda | 2025 | Safety and Efficacy of Ecopipam in Patients with Tourette Syndrome: A Systematic Review and Meta-analysis. [224] | CNS Drugs | Tourette Syndrome; Dopamine Antagonists (major MeSH headings from PubMed PMID 39730854) | Intervention | Yes |  |  |
| 220 | 72 | Non-AI | Mendelsohn | 2022 | Commercial volumetric MRI reporting tools in multiple sclerosis: a systematic review of the evidence. [225] | Neuroradiology | Neuroradiology | Diagnostic | No |  |  |
| 221 | 72 | Non-AI | Huang | 2023 | Diagnostic value of magnetic resonance imaging and magnetic resonance arthrography for assessing acetabular labral tears: A systematic review and meta-analysis. [226] | Medicine | idiopathic pulmonary fibrosis | Diagnostic | Yes |  |  |
| 222 | 73 | Non-AI | Olufadewa | 2025 | Quantitative summarization of high-touch surfaces and epidemiological parameters of Clostridioides difficile acquisition and transmission for mathematical modeling: a systematic review. [227] | Infection Control and Hospital Epidemiology | Healthcare epidemiology / Infection control | Other | No |  |  |
| 223 | 73 | Non-AI | Almutawif | 2025 | Evaluation of probiotics efficiency for the prevention of Clostridioides difficile infection in hospitalized patients: a systematic review and meta-analysis. [228] | Infection | Infectious disease | Intervention | Yes |  |  |
| 224 | 74 | Non-AI | Yang | 2025 | Efficacy and Safety of Anticoagulants in Patients With Idiopathic Pulmonary Fibrosis: A Meta-Analysis. [229] | Canadian Respiratory Journal | Idiopathic Pulmonary Fibrosis; Anticoagulants (PubMed major MeSH: Anticoagulants [Y], Idiopathic Pulmonary Fibrosis [Y]) | Intervention | Yes |  |  |
| 225 | 74 | Non-AI | He | 2025 | Rivaroxaban vs. Enoxaparin for Preventing Venous Thromboembolism and Wound Complications after Knee Surgery: A Meta-Analysis. [230] | Journal of the College of Physicians and Surgeons Pakistan | Orthopedic surgery | Intervention | Yes |  |  |
| 226 | 75 | Non-AI | Tomescu-Stachie | 2025 | The impact of psychosocial interventions on women with gynaecological cancers: a systematic review and meta-analysis. [231] | Health Psychology Review | Psychology | Intervention | Yes |  |  |
| 227 | 75 | Non-AI | Dewi | 2025 | Effectiveness of parenting interventions on self-efficacy, anxiety, stress, and depression among parents of preterm infants: A systematic review and meta-analysis of randomized control trials. [232] | International Journal of Nursing Studies | Therapy | Intervention | Yes |  |  |
| 228 | 76 | Non-AI | Sanchez-Escudero | 2025 | Virtual Reality and Serious Videogame-Based Instruments for Assessing Spatial Navigation in Alzheimer's Disease: A Systematic Review of Psychometric Properties. [233] | Neuropsychology Review | Neuropsychology / Alzheimer's disease | Diagnostic | No |  |  |
| 229 | 76 | Non-AI | Bao | 2024 | Is there a bilingual advantage in auditory attention among children? A systematic review and meta-analysis of standardized auditory attention tests. [234] | Plos One | Multilingualism, Attention | Qualitative | Yes |  |  |
| 230 | 77 | Non-AI | de Macedo | 2024 | The Impact of Positive Expiratory Pressure Therapy on Hyperinflation in Patients With COPD. [235] | Respiratory Care | Pulmonary Disease, Chronic Obstructive; Positive-Pressure Respiration | Intervention | Yes |  |  |
| 231 | 77 | Non-AI | Oriecuia | 2023 | Sodium glucose co-transporter 2 inhibitors and quality of life in patients with heart failure: a comprehensive systematic review and meta-analysis of randomized controlled trials. [236] | European Heart Journal Cardiovascular Pharmacotherapy | Cardiovascular | Intervention | Yes |  |  |

Note. AI denotes artificial intelligence; LLM, large language model. AI tool and stage-of-use fields apply only to AI-assisted reviews. Supplementary reference numbers are shown in brackets after article titles. Complete machine-readable study-level data, including domain- and item-level appraisal scores, are provided in Multimedia Appendix 2.

Table S3. Individual-Study Methodological Quality, Reporting, Risk-of-Bias, and AI-Transparency Outcomes

| **Manuscript ID** | **Matched set ID** | **Review group** | **First author** | **Year** | **AMSTAR-2 13-item score** | **Adapted 13-item AMSTAR-2 confidence category** | **AMSTAR-2 16-item score (meta-analysis reviews only)** | **PRISMA adherence proportion** | **ROBIS overall rating** | **AITDI total score (AI-assisted only)** |
| --- | --- | --- | --- | --- | --- | --- | --- | --- | --- | --- |
| 1 | 1 | AI-assisted | Aceves-Martins | 2024 | 6 | Critically low | NA | 72.2% | Low | 3 |
| 2 | 2 | AI-assisted | Adu | 2024 | 5 | Critically low | NA | 53.7% | Low | 2 |
| 3 | 3 | AI-assisted | Ali | 2025 | 5 | Critically low | NA | 63.0% | High | 5 |
| 4 | 4 | AI-assisted | Allen | 2024 | 6 | Critically low | NA | 63.0% | Low | 5 |
| 5 | 5 | AI-assisted | Alonso-Sanchez | 2024 | 6 | Critically low | 7 | 74.1% | Low | 3 |
| 6 | 6 | AI-assisted | Anghelescu | 2025 | 2 | Critically low | 3 | 37.0% | High | 5 |
| 7 | 7 | AI-assisted | Arzani | 2025 | 7 | Critically low | 10 | 92.6% | Low | 4 |
| 8 | 8 | AI-assisted | Beecham | 2025 | 8 | Low | NA | 70.4% | Low | 5 |
| 9 | 9 | AI-assisted | Bell-Aldeghi | 2023 | 6 | Critically low | 9 | 75.9% | Low | 5 |
| 10 | 10 | AI-assisted | Bennett | 2024 | 10 | Low | NA | 100.0% | Low | 4 |
| 11 | 11 | AI-assisted | Bourke | 2025 | 9 | Critically low | 9 | 77.8% | Low | 2 |
| 12 | 12 | AI-assisted | Bourke | 2023 | 11 | Low | 12 | 79.6% | Low | 4 |
| 13 | 13 | AI-assisted | Brozek | 2024 | 6 | Low | NA | 81.5% | Low | 3 |
| 14 | 14 | AI-assisted | Burger | 2024 | 8 | Critically low | 11 | 83.3% | Low | 4 |
| 15 | 15 | AI-assisted | Cale | 2025 | 5 | Critically low | NA | 70.4% | Low | 3 |
| 16 | 16 | AI-assisted | Canfell | 2024 | 10 | Low | 10 | 74.1% | Low | 5 |
| 17 | 17 | AI-assisted | De Silva | 2024 | 6 | Critically low | NA | 51.9% | Not applicable | 4 |
| 18 | 18 | AI-assisted | Deering | 2025 | 4 | Critically low | NA | 50.0% | High | 5 |
| 19 | 19 | AI-assisted | Demirtas Yilmaz | 2025 | 6 | Critically low | NA | 59.3% | High | 4 |
| 20 | 20 | AI-assisted | Ducreux | 2025 | 12 | High | NA | 92.6% | Low | 5 |
| 21 | 21 | AI-assisted | Fajkis-Zajaczkowska | 2025 | 8 | Critically low | NA | 51.9% | High | 5 |
| 22 | 22 | AI-assisted | Feil | 2023 | 9 | Low | 12 | 88.9% | Low | 4 |
| 23 | 23 | AI-assisted | Feng | 2025 | 10 | Critically low | 11 | 88.9% | Low | 5 |
| 24 | 24 | AI-assisted | Franzoi | 2023 | 9 | Critically low | NA | 77.8% | High | 5 |
| 25 | 25 | AI-assisted | Gabriel | 2023 | 7 | Critically low | 8 | 64.8% | Low | 4 |
| 26 | 26 | AI-assisted | Garcia Valencia | 2025 | 11 | Low | NA | 81.5% | Low | 5 |
| 27 | 27 | AI-assisted | Garcia-Torres | 2024 | 8 | Critically low | NA | 68.5% | High | 6 |
| 28 | 28 | AI-assisted | Garg | 2023 | 7 | Critically low | NA | 46.3% | High | 3 |
| 29 | 29 | AI-assisted | Holland | 2025 | 6 | Critically low | NA | 44.4% | High | 3 |
| 30 | 30 | AI-assisted | Hsieh | 2024 | 11 | Low | NA | 94.4% | Low | 3 |
| 31 | 31 | AI-assisted | Iaconisi | 2024 | 4 | Critically low | NA | 40.7% | High | 3 |
| 32 | 32 | AI-assisted | Isern-de-Val | 2025 | 7 | Critically low | 9 | 72.2% | Low | 4 |
| 33 | 33 | AI-assisted | Kelani | 2024 | 7 | Critically low | 8 | 75.9% | Low | 3 |
| 34 | 34 | AI-assisted | Ketelaars | 2025 | 11 | Low | NA | 79.6% | Low | 4 |
| 35 | 35 | AI-assisted | Kumar | 2025 | 9 | Critically low | 10 | 87.0% | Low | 3 |
| 36 | 36 | AI-assisted | Kwarah | 2025 | 11 | Low | NA | 88.9% | Low | 3 |
| 37 | 37 | AI-assisted | Landau | 2024 | 8 | Critically low | NA | 70.4% | High | 4 |
| 38 | 38 | AI-assisted | Lauria | 2023 | 4 | Critically low | NA | 37.0% | High | 3 |
| 39 | 39 | AI-assisted | Lee | 2024 | 10 | Low | 12 | 87.0% | Low | 5 |
| 40 | 40 | AI-assisted | Li | 2023 | 9 | Critically low | 11 | 81.5% | Low | 3 |
| 41 | 41 | AI-assisted | Liu | 2025 | 11 | Low | 13 | 92.6% | Low | 5 |
| 42 | 42 | AI-assisted | Matsui | 2024 | 7 | Critically low | NA | 70.4% | Low | 5 |
| 43 | 43 | AI-assisted | Miao | 2024 | 7 | Critically low | NA | 74.1% | High | 5 |
| 44 | 44 | AI-assisted | Mohamed | 2025 | 11 | Low | NA | 79.6% | Low | 3 |
| 45 | 45 | AI-assisted | Molan | 2025 | 8 | Critically low | NA | 72.2% | High | 6 |
| 46 | 46 | AI-assisted | Mondal | 2024 | 6 | Critically low | 6 | 63.0% | Low | 3 |
| 47 | 47 | AI-assisted | Mushcab | 2025 | 9 | Critically low | NA | 81.5% | Low | 3 |
| 48 | 48 | AI-assisted | Noe-Steinmuller | 2024 | 3 | Critically low | NA | 59.3% | Low | 5 |
| 49 | 49 | AI-assisted | Nuvoli | 2025 | 6 | Critically low | NA | 63.0% | Low | 3 |
| 50 | 50 | AI-assisted | Park | 2025 | 2 | Critically low | 2 | 63.0% | High | 3 |
| 51 | 51 | AI-assisted | Perronne | 2025 | 8 | Critically low | NA | 57.4% | Low | 3 |
| 52 | 52 | AI-assisted | Peterson | 2024 | 9 | Low | 12 | 92.6% | Low | 3 |
| 53 | 53 | AI-assisted | Polin | 2025 | 2 | Critically low | NA | 18.5% | High | 4 |
| 54 | 54 | AI-assisted | PoshtMashhadi | 2025 | 9 | Low | NA | 100.0% | Low | 3 |
| 55 | 55 | AI-assisted | Purewal | 2025 | 8 | Critically low | 9 | 70.4% | Low | 5 |
| 56 | 56 | AI-assisted | Rakhshandehroo | 2023 | 10 | Low | NA | 88.9% | Low | 4 |
| 57 | 57 | AI-assisted | Rich | 2025 | 8 | Critically low | 8 | 70.4% | Low | 3 |
| 58 | 58 | AI-assisted | Robinson | 2024 | 9 | Critically low | NA | 77.8% | Low | 3 |
| 59 | 59 | AI-assisted | Ruksakulpiwat | 2024 | 6 | Critically low | NA | 63.0% | Low | 5 |
| 60 | 60 | AI-assisted | Schopow | 2023 | 3 | Critically low | NA | 55.6% | Low | 6 |
| 61 | 61 | AI-assisted | Shaffer | 2025 | 11 | Low | NA | 96.3% | Low | 3 |
| 62 | 62 | AI-assisted | Srinivasan | 2025 | 8 | Critically low | 8 | 55.6% | High | 5 |
| 63 | 63 | AI-assisted | Stafford | 2023 | 11 | Critically low | 14 | 96.3% | Low | 2 |
| 64 | 64 | AI-assisted | Steele | 2025 | 8 | Critically low | 10 | 66.7% | Low | 3 |
| 65 | 65 | AI-assisted | Talukdar | 2025 | 8 | Critically low | 10 | 74.1% | Low | 3 |
| 66 | 66 | AI-assisted | Teo | 2024 | 6 | Critically low | NA | 55.6% | Low | 2 |
| 67 | 67 | AI-assisted | Teperikidis | 2025 | 8 | Critically low | 11 | 87.0% | Low | 4 |
| 68 | 68 | AI-assisted | Trivedi | 2024 | 13 | High | 16 | 92.6% | Low | 2 |
| 69 | 69 | AI-assisted | Urru | 2024 | 11 | Low | 14 | 96.3% | Low | 2 |
| 70 | 70 | AI-assisted | Visser | 2025 | 7 | Critically low | NA | 68.5% | Low | 3 |
| 71 | 71 | AI-assisted | Vizcarra | 2024 | 5 | Critically low | NA | 50.0% | Low | 4 |
| 72 | 72 | AI-assisted | Warren | 2023 | 3 | Critically low | NA | 44.4% | High | 4 |
| 73 | 73 | AI-assisted | Weerakoon | 2025 | 9 | Critically low | NA | 66.7% | Low | 4 |
| 74 | 74 | AI-assisted | Wen | 2025 | 10 | Low | 13 | 85.2% | Low | 5 |
| 75 | 75 | AI-assisted | Zhu | 2025 | 10 | Low | 13 | 77.8% | Low | 5 |
| 76 | 76 | AI-assisted | de Zwart | 2024 | 2 | Critically low | NA | 40.7% | High | 3 |
| 77 | 77 | AI-assisted | van Dijk | 2024 | 8 | Low | NA | 66.7% | Low | 5 |
| 78 | 1 | Non-AI | Riley | 2024 | 11 | Low | 13 | 92.6% | Low | NA |
| 79 | 1 | Non-AI | Bakhshimoghaddam | 2024 | 11 | Low | 14 | 94.4% | Low | NA |
| 80 | 2 | Non-AI | Khawaja | 2025 | 6 | Critically low | NA | 48.1% | High | NA |
| 81 | 2 | Non-AI | Alizadeh | 2024 | 10 | Critically low | 12 | 88.9% | Low | NA |
| 82 | 3 | Non-AI | Ngo | 2025 | 13 | High | 16 | 96.3% | Low | NA |
| 83 | 3 | Non-AI | Ramah | 2025 | 10 | Low | NA | 72.2% | Low | NA |
| 84 | 4 | Non-AI | Akdemir | 2024 | 9 | Critically low | 11 | 83.3% | Low | NA |
| 85 | 4 | Non-AI | Mestre-Ferrandiz | 2024 | 4 | Critically low | NA | 37.0% | High | NA |
| 86 | 5 | Non-AI | Arnautovska | 2024 | 11 | Low | 14 | 100.0% | Low | NA |
| 87 | 5 | Non-AI | Mitra | 2024 | 7 | Critically low | NA | 44.4% | High | NA |
| 88 | 6 | Non-AI | Contreras-Ruston | 2025 | 3 | Critically low | NA | 44.4% | High | NA |
| 89 | 6 | Non-AI | Triolo | 2025 | 9 | Critically low | NA | 77.8% | Low | NA |
| 90 | 7 | Non-AI | Adoberg | 2025 | 10 | Critically low | NA | 68.5% | Low | NA |
| 91 | 7 | Non-AI | Kyriakoulis | 2025 | 9 | Critically low | 11 | 85.2% | Low | NA |
| 92 | 8 | Non-AI | Alexandrou | 2025 | 10 | Low | 11 | 72.2% | High | NA |
| 93 | 8 | Non-AI | Zhang | 2025 | 11 | Low | NA | 81.5% | Low | NA |
| 94 | 9 | Non-AI | Zhang | 2024 | 11 | Low | 12 | 75.9% | Low | NA |
| 95 | 9 | Non-AI | Sangalli | 2023 | 11 | Low | NA | 77.8% | Low | NA |
| 96 | 10 | Non-AI | MacLehose | 2025 | 7 | Critically low | 10 | 77.8% | High | NA |
| 97 | 10 | Non-AI | Zhang | 2024 | 11 | Low | 13 | 83.3% | Low | NA |
| 98 | 11 | Non-AI | Lee | 2025 | 12 | High | 15 | 92.6% | Low | NA |
| 99 | 11 | Non-AI | Rostami Haji Abadi | 2025 | 11 | Low | NA | 88.9% | Low | NA |
| 100 | 12 | Non-AI | Larouche | 2023 | 8 | Critically low | NA | 88.9% | Low | NA |
| 101 | 12 | Non-AI | Weng | 2023 | 12 | Low | 15 | 100.0% | Low | NA |
| 102 | 13 | Non-AI | He | 2024 | 11 | Low | 12 | 85.2% | Low | NA |
| 103 | 13 | Non-AI | Shao | 2024 | 9 | Critically low | 11 | 90.7% | Low | NA |
| 104 | 14 | Non-AI | Holanda da Cunha | 2025 | 12 | Low | NA | 88.9% | Low | NA |
| 105 | 14 | Non-AI | Cassioli | 2024 | 5 | Critically low | 6 | 72.2% | High | NA |
| 106 | 15 | Non-AI | Firouzabadi | 2025 | 4 | Critically low | NA | 42.6% | High | NA |
| 107 | 15 | Non-AI | Oyowvi | 2025 | 6 | Critically low | NA | 51.9% | High | NA |
| 108 | 16 | Non-AI | Miake-Lye | 2023 | 9 | Critically low | NA | 63.0% | High | NA |
| 109 | 16 | Non-AI | Bertocchi | 2024 | 6 | Critically low | NA | 55.6% | Unclear | NA |
| 110 | 17 | Non-AI | Elfios | 2024 | 10 | Critically low | 13 | 87.0% | Low | NA |
| 111 | 17 | Non-AI | Shetty | 2023 | 10 | Critically low | 12 | 83.3% | Low | NA |
| 112 | 18 | Non-AI | Karamitros | 2025 | 11 | Low | NA | 77.8% | Low | NA |
| 113 | 18 | Non-AI | Acosta | 2025 | 9 | Low | NA | 72.2% | Low | NA |
| 114 | 19 | Non-AI | Nair | 2025 | 7 | Critically low | NA | 74.1% | Low | NA |
| 115 | 19 | Non-AI | Padurean | 2025 | 10 | Low | NA | 77.8% | Low | NA |
| 116 | 20 | Non-AI | de Souza Gaio | 2025 | 11 | Low | 13 | 88.9% | Low | NA |
| 117 | 20 | Non-AI | Maguire | 2025 | 8 | Critically low | 9 | 87.0% | Low | NA |
| 118 | 21 | Non-AI | Chiang | 2025 | 12 | High | 15 | 92.6% | Low | NA |
| 119 | 21 | Non-AI | Stockdale | 2025 | 11 | Low | 13 | 88.9% | Low | NA |
| 120 | 22 | Non-AI | Guimaraes | 2023 | 11 | Low | 14 | 92.6% | Low | NA |
| 121 | 22 | Non-AI | Wang | 2023 | 11 | Low | 14 | 92.6% | Low | NA |
| 122 | 23 | Non-AI | Meszaros | 2025 | 11 | Low | 12 | 79.6% | High | NA |
| 123 | 23 | Non-AI | Chen | 2025 | 9 | Critically low | 11 | 83.3% | High | NA |
| 124 | 24 | Non-AI | Bali | 2024 | 6 | Critically low | NA | 63.0% | High | NA |
| 125 | 24 | Non-AI | Sutarsa | 2024 | 8 | Critically low | NA | 81.5% | Unclear | NA |
| 126 | 25 | Non-AI | Diaz | 2023 | 3 | Critically low | NA | 46.3% | High | NA |
| 127 | 25 | Non-AI | Seffah | 2023 | 7 | Critically low | NA | 53.7% | High | NA |
| 128 | 26 | Non-AI | Muxunov | 2026 | 13 | High | 16 | 92.6% | Low | NA |
| 129 | 26 | Non-AI | Chen | 2025 | 11 | Low | 14 | 87.0% | High | NA |
| 130 | 27 | Non-AI | Hou | 2025 | 7 | Critically low | NA | 63.0% | High | NA |
| 131 | 27 | Non-AI | Rohani | 2024 | 12 | High | NA | 77.8% | Low | NA |
| 132 | 28 | Non-AI | Casey | 2023 | 7 | Critically low | NA | 64.8% | High | NA |
| 133 | 28 | Non-AI | Ahmed | 2023 | 3 | Critically low | NA | 46.3% | High | NA |
| 134 | 29 | Non-AI | Li | 2024 | 11 | Low | 13 | 85.2% | Low | NA |
| 135 | 29 | Non-AI | Wang | 2025 | 12 | High | 15 | 92.6% | Low | NA |
| 136 | 30 | Non-AI | Gandy | 2024 | 11 | Low | 14 | 92.6% | Low | NA |
| 137 | 30 | Non-AI | Felisbino | 2025 | 12 | High | NA | 75.9% | Low | NA |
| 138 | 31 | Non-AI | Rehim | 2024 | 11 | Low | 14 | 87.0% | Low | NA |
| 139 | 31 | Non-AI | Wang | 2024 | 12 | Low | 15 | 100.0% | Low | NA |
| 140 | 32 | Non-AI | Couppoussamy | 2026 | 10 | Low | 13 | 96.3% | Low | NA |
| 141 | 32 | Non-AI | Tejada | 2025 | 11 | Low | 14 | 100.0% | Low | NA |
| 142 | 33 | Non-AI | Geravandi | 2024 | 8 | Critically low | 11 | 87.0% | Low | NA |
| 143 | 33 | Non-AI | Ghozy | 2025 | 10 | Critically low | 12 | 81.5% | Low | NA |
| 144 | 34 | Non-AI | Karimi | 2025 | 7 | Critically low | NA | 68.5% | Low | NA |
| 145 | 34 | Non-AI | Kumar | 2025 | 10 | Critically low | 11 | 87.0% | High | NA |
| 146 | 35 | Non-AI | Jayawardane | 2025 | 10 | Low | NA | 92.6% | Low | NA |
| 147 | 35 | Non-AI | Topala | 2025 | 11 | Low | 13 | 96.3% | Low | NA |
| 148 | 36 | Non-AI | Mukoka | 2025 | 12 | High | 15 | 90.7% | Low | NA |
| 149 | 36 | Non-AI | Strauss | 2025 | 11 | Low | 14 | 92.6% | High | NA |
| 150 | 37 | Non-AI | Goh | 2025 | 10 | Critically low | 13 | 88.9% | Low | NA |
| 151 | 37 | Non-AI | Valentine | 2024 | 9 | Low | NA | 72.2% | Low | NA |
| 152 | 38 | Non-AI | Hong | 2023 | 10 | Critically low | 13 | 92.6% | Low | NA |
| 153 | 38 | Non-AI | Lee | 2024 | 9 | Critically low | NA | 72.2% | High | NA |
| 154 | 39 | Non-AI | Fonseca | 2024 | 12 | High | 15 | 94.4% | Low | NA |
| 155 | 39 | Non-AI | Lechien | 2023 | 11 | Low | NA | 70.4% | High | NA |
| 156 | 40 | Non-AI | Qu | 2023 | 8 | Critically low | 11 | 92.6% | Low | NA |
| 157 | 40 | Non-AI | Lu | 2023 | 10 | Critically low | 13 | 81.5% | High | NA |
| 158 | 41 | Non-AI | Rostampour | 2025 | 11 | Low | 14 | 100.0% | Low | NA |
| 159 | 41 | Non-AI | Flores-Hernández | 2024 | 6 | Critically low | NA | 51.9% | High | NA |
| 160 | 42 | Non-AI | Wen | 2024 | 4 | Critically low | NA | 51.9% | High | NA |
| 161 | 42 | Non-AI | Khalid | 2025 | 9 | Critically low | NA | 74.1% | High | NA |
| 162 | 43 | Non-AI | Gilbert | 2024 | 9 | Critically low | NA | 61.1% | High | NA |
| 163 | 43 | Non-AI | Dadon | 2024 | 6 | Critically low | NA | 55.6% | Unclear | NA |
| 164 | 44 | Non-AI | Dumbuya | 2025 | 11 | Low | NA | 74.1% | Low | NA |
| 165 | 44 | Non-AI | Han | 2025 | 8 | Critically low | NA | 61.1% | High | NA |
| 166 | 45 | Non-AI | Wan Mohd Johari | 2025 | 11 | Moderate | NA | 81.5% | Low | NA |
| 167 | 45 | Non-AI | Caut | 2025 | 11 | Moderate | NA | 77.8% | Low | NA |
| 168 | 46 | Non-AI | Abdallah | 2024 | 10 | Critically low | 12 | 85.2% | High | NA |
| 169 | 46 | Non-AI | Hwang | 2024 | 10 | Critically low | 11 | 85.2% | Low | NA |
| 170 | 47 | Non-AI | Hanafi | 2025 | 11 | Moderate | 14 | 88.9% | Low | NA |
| 171 | 47 | Non-AI | Meillan | 2025 | 8 | Critically low | NA | 70.4% | Low | NA |
| 172 | 48 | Non-AI | Liu | 2024 | 13 | High | 16 | 88.9% | Low | NA |
| 173 | 48 | Non-AI | Altuhafy | 2024 | 13 | High | NA | 75.9% | High | NA |
| 174 | 49 | Non-AI | Lima | 2025 | 6 | Critically low | NA | 61.1% | High | NA |
| 175 | 49 | Non-AI | Zgambo | 2025 | 10 | Critically low | 13 | 83.3% | Low | NA |
| 176 | 50 | Non-AI | Clack | 2025 | 10 | Low | NA | 61.1% | High | NA |
| 177 | 50 | Non-AI | Seddik | 2025 | 10 | Critically low | NA | 70.4% | Low | NA |
| 178 | 51 | Non-AI | Xie | 2026 | 12 | High | 15 | 88.9% | Low | NA |
| 179 | 51 | Non-AI | Liu | 2025 | 11 | Moderate | 14 | 85.2% | Low | NA |
| 180 | 52 | Non-AI | Long | 2024 | 8 | Critically low | 10 | 68.5% | Unclear | NA |
| 181 | 52 | Non-AI | Frisira | 2025 | 10 | Low | 11 | 83.3% | Low | NA |
| 182 | 53 | Non-AI | Dejakaisaya | 2025 | 11 | Low | 14 | 100.0% | Low | NA |
| 183 | 53 | Non-AI | Carollo | 2025 | 5 | Critically low | NA | 40.7% | High | NA |
| 184 | 54 | Non-AI | N Alaqil | 2025 | 10 | Critically low | NA | 77.8% | Low | NA |
| 185 | 54 | Non-AI | Abram | 2025 | 12 | High | NA | 74.1% | High | NA |
| 186 | 55 | Non-AI | Russo | 2025 | 11 | Low | 13 | 92.6% | Low | NA |
| 187 | 55 | Non-AI | Huygen | 2024 | 10 | Critically low | 12 | 90.7% | Low | NA |
| 188 | 56 | Non-AI | Zhao | 2023 | 11 | Low | NA | 74.1% | Low | NA |
| 189 | 56 | Non-AI | Narimani | 2025 | 12 | High | NA | 92.6% | Low | NA |
| 190 | 57 | Non-AI | Gonzalez | 2025 | 12 | High | 14 | 90.7% | Low | NA |
| 191 | 57 | Non-AI | Lagden | 2025 | 12 | High | 15 | 81.5% | Low | NA |
| 192 | 58 | Non-AI | Collonnaz | 2024 | 7 | Critically low | NA | 66.7% | High | NA |
| 193 | 58 | Non-AI | Xu | 2023 | 8 | Critically low | NA | 63.0% | High | NA |
| 194 | 59 | Non-AI | Teti | 2025 | 10 | Critically low | NA | 85.2% | Low | NA |
| 195 | 59 | Non-AI | Pannunzio | 2024 | 9 | Critically low | 9 | 57.4% | High | NA |
| 196 | 60 | Non-AI | Jiao | 2024 | 11 | Critically low | NA | 74.1% | Low | NA |
| 197 | 60 | Non-AI | Samartha | 2024 | 6 | Critically low | NA | 50.0% | High | NA |
| 198 | 61 | Non-AI | Abdelkhader | 2025 | 11 | Low | 14 | 92.6% | Low | NA |
| 199 | 61 | Non-AI | Ou | 2025 | 7 | Critically low | 9 | 79.6% | Low | NA |
| 200 | 62 | Non-AI | Erqou | 2025 | 9 | Critically low | 11 | 90.7% | Low | NA |
| 201 | 62 | Non-AI | Soma | 2025 | 11 | Low | 14 | 100.0% | Low | NA |
| 202 | 63 | Non-AI | Hay | 2023 | 12 | High | 15 | 100.0% | Low | NA |
| 203 | 63 | Non-AI | Zhuang | 2024 | 12 | High | 15 | 87.0% | Low | NA |
| 204 | 64 | Non-AI | Pastre | 2025 | 11 | Moderate | 14 | 87.0% | Low | NA |
| 205 | 64 | Non-AI | Pellegrini | 2025 | 7 | Critically low | 8 | 48.1% | Unclear | NA |
| 206 | 65 | Non-AI | Chen | 2025 | 12 | Low | 14 | 96.3% | Low | NA |
| 207 | 65 | Non-AI | Korsa | 2025 | 11 | Low | NA | 81.5% | Low | NA |
| 208 | 66 | Non-AI | Pigoni | 2024 | 9 | Critically low | NA | 70.4% | Low | NA |
| 209 | 66 | Non-AI | Issaiy | 2023 | 11 | Low | NA | 79.6% | High | NA |
| 210 | 67 | Non-AI | Akankwasa | 2025 | 11 | Low | NA | 74.1% | High | NA |
| 211 | 67 | Non-AI | Siargkas | 2025 | 11 | Moderate | 14 | 88.9% | Low | NA |
| 212 | 68 | Non-AI | Seidler | 2023 | 12 | Low | 14 | 94.4% | Low | NA |
| 213 | 68 | Non-AI | Zeng | 2023 | 11 | Low | 14 | 92.6% | Low | NA |
| 214 | 69 | Non-AI | Poon | 2024 | 12 | High | NA | 81.5% | Low | NA |
| 215 | 69 | Non-AI | Hu | 2024 | 10 | Critically low | 13 | 87.0% | High | NA |
| 216 | 70 | Non-AI | Zhao | 2025 | 12 | High | 15 | 92.6% | High | NA |
| 217 | 70 | Non-AI | Liu | 2025 | 4 | Critically low | NA | 33.3% | High | NA |
| 218 | 71 | Non-AI | Davydow | 2025 | 7 | Critically low | NA | 59.3% | High | NA |
| 219 | 71 | Non-AI | Panda | 2025 | 8 | Critically low | 10 | 92.6% | Low | NA |
| 220 | 72 | Non-AI | Mendelsohn | 2022 | 8 | Critically low | NA | 66.7% | Low | NA |
| 221 | 72 | Non-AI | Huang | 2023 | 12 | High | 14 | 81.5% | Low | NA |
| 222 | 73 | Non-AI | Olufadewa | 2025 | 8 | Critically low | NA | 92.6% | High | NA |
| 223 | 73 | Non-AI | Almutawif | 2025 | 9 | Critically low | 12 | 88.9% | High | NA |
| 224 | 74 | Non-AI | Yang | 2025 | 10 | Critically low | 12 | 83.3% | High | NA |
| 225 | 74 | Non-AI | He | 2025 | 10 | Critically low | 13 | 79.6% | Low | NA |
| 226 | 75 | Non-AI | Tomescu-Stachie | 2025 | 13 | High | 16 | 100.0% | Low | NA |
| 227 | 75 | Non-AI | Dewi | 2025 | 10 | Critically low | 12 | 92.6% | Low | NA |
| 228 | 76 | Non-AI | Sanchez-Escudero | 2025 | 11 | Low | NA | 83.3% | Low | NA |
| 229 | 76 | Non-AI | Bao | 2024 | 11 | Low | 14 | 88.9% | Low | NA |
| 230 | 77 | Non-AI | de Macedo | 2024 | 11 | Moderate | 12 | 77.8% | Low | NA |
| 231 | 77 | Non-AI | Oriecuia | 2023 | 9 | Critically low | 11 | 75.9% | Low | NA |

Note. AMSTAR-2 13-item scores range from 0 to 13 and 16-item scores from 0 to 16, with higher scores indicating greater methodological adherence; the prespecified noninferiority margin for the 13-item score was -1.0 point, and no established minimally important difference is available.[3] PRISMA adherence ranges from 0 to 1, with higher values indicating more complete reporting.[4] ROBIS ratings are Low, High, Unclear, or Not applicable; Low is favorable.[5] AITDI scores range from 0 to 6, with higher scores indicating more complete AI-use reporting, and apply only to AI-assisted reviews.[1] NA denotes not applicable. Complete machine-readable study-level data are provided in Multimedia Appendix 2.

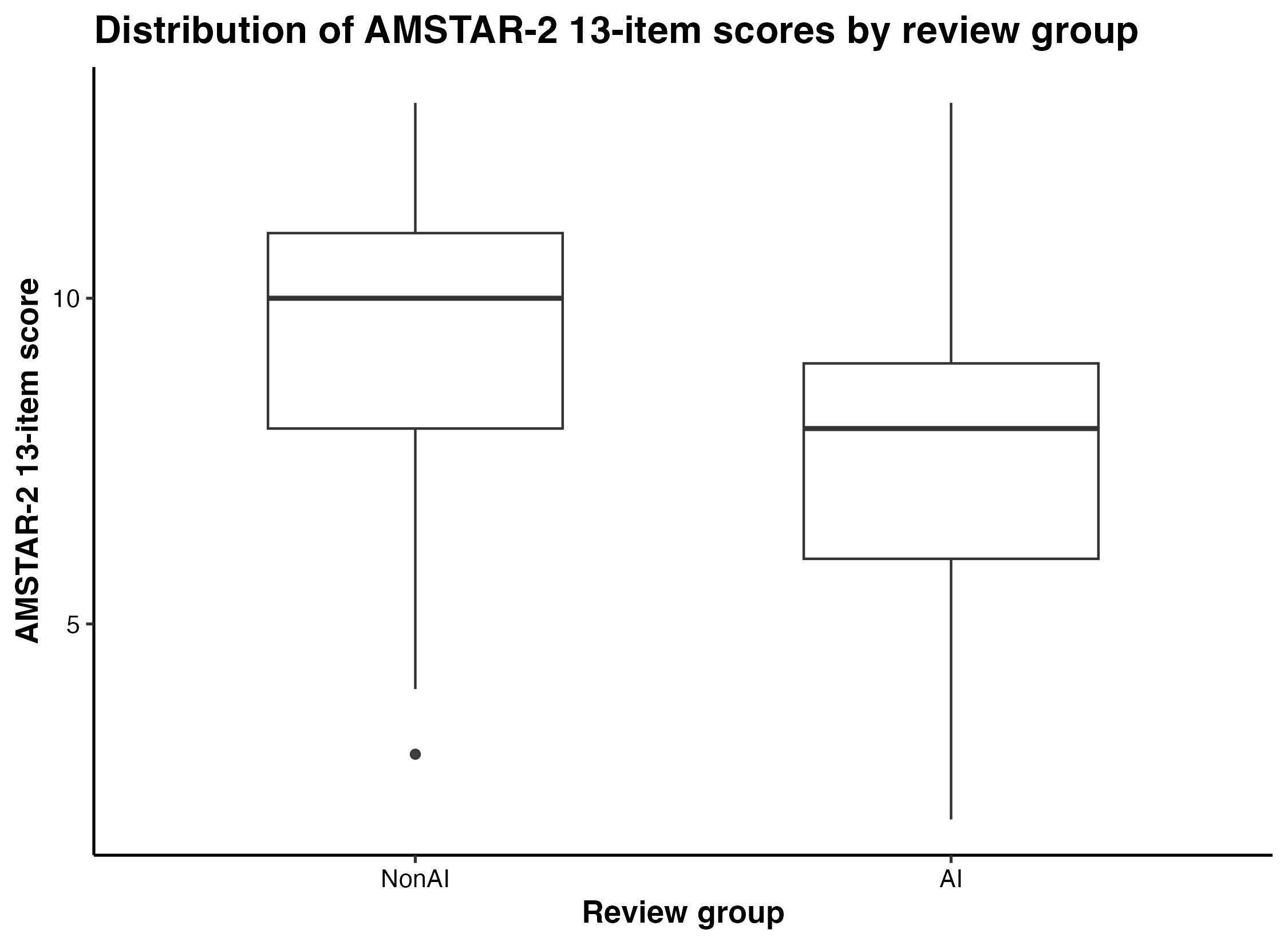

Figure S1. Distribution of AMSTAR-2 13-Item Scores by Review Group

Legend. Boxes show the interquartile range, horizontal lines show medians, whiskers extend to 1.5 times the interquartile range, and points indicate outliers. The modified AMSTAR-2 adherence score ranges from 0 to 13, with higher scores indicating greater methodological adherence.[3] The prespecified noninferiority margin for the AI-minus-non-AI difference was -1.0 point; no validated minimally important difference is established for this modified score. AI denotes artificial intelligence; AMSTAR-2, A Measurement Tool to Assess Systematic Reviews, version 2.

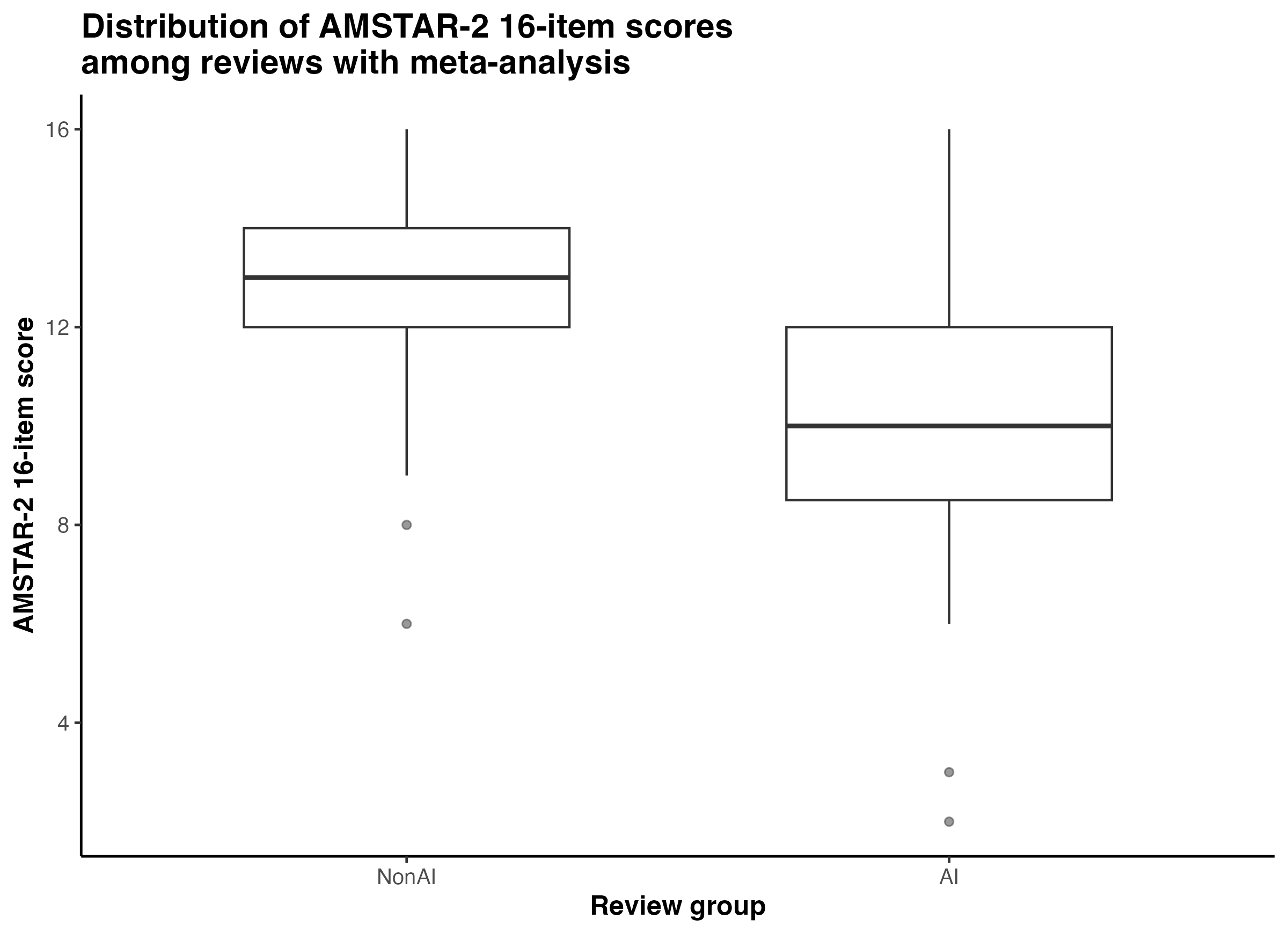

Figure S2. Distribution of AMSTAR-2 16-Item Scores Among Reviews with Meta-Analysis

Legend. Boxes show the interquartile range, horizontal lines show medians, whiskers extend to 1.5 times the interquartile range, and points indicate outliers. The AMSTAR-2 adherence score ranges from 0 to 16, with higher scores indicating greater methodological adherence.[3] This analysis was restricted to reviews with meta-analysis; no noninferiority margin or validated minimally important difference was specified for this secondary outcome. AI denotes artificial intelligence; AMSTAR-2, A Measurement Tool to Assess Systematic Reviews, version 2.

Table S4. Adapted 13-Item AMSTAR-2 Confidence Categories by Review Group

| Adapted 13-item confidence category | **AI-assisted reviews, n/N (%)** | **Non-AI reviews, n/N (%)** |
| --- | --- | --- |
| High | 2/77 (2.6%) | 22/154 (14.3%) |
| Moderate | 0/77 (0.0%) | 7/154 (4.5%) |
| Low | 21/77 (27.3%) | 50/154 (32.5%) |
| Critically low | 54/77 (70.1%) | 75/154 (48.7%) |

Note. Confidence categories were recalculated descriptively from the reconciled modified 13-item ratings using an adapted AMSTAR-2 framework. Critical items were 2, 4, 7, 9, and 13; items 11, 12, and 15 were excluded. Only a full Yes satisfied an item; Partial Yes and No counted as weaknesses. High indicated no critical weaknesses and at most one noncritical weakness; moderate, no critical weaknesses and more than one noncritical weakness; low, one critical weakness; and critically low, more than one critical weakness. These adapted categories are not original AMSTAR-2 overall confidence ratings. Percentages use all reviews in each group as the denominator.

Table S5. Descriptive AMSTAR-2 Critical- and Noncritical-Domain Scores by Review Group

| **Outcome** | **Non-AI n** | **Non-AI mean (SD)** | **Non-AI median [IQR]** | **AI-assisted n** | **AI-assisted mean (SD)** | **AI-assisted median [IQR]** |
| --- | --- | --- | --- | --- | --- | --- |
| AMSTAR-2 13-item score | 154 | 9.63 (2.27) | 10 [8–11] | 77 | 7.58 (2.60) | 8 [6–9] |
| Critical-domain score, 5 items | 154 | 3.34 (1.28) | 4 [3–4] | 77 | 2.55 (1.28) | 3 [1–4] |
| Noncritical-domain score, 8 items | 154 | 6.29 (1.26) | 7 [6–7] | 77 | 5.04 (1.74) | 5 [4–6] |
| Critical-domain adherence proportion | 154 | 0.67 (0.26) | 0.80 [0.60–0.80] | 77 | 0.51 (0.26) | 0.60 [0.20–0.80] |
| Noncritical-domain adherence proportion | 154 | 0.79 (0.16) | 0.88 [0.75–0.88] | 77 | 0.63 (0.22) | 0.62 [0.50–0.75] |

Note. The 5-item critical-domain score ranges from 0 to 5 and the 8-item noncritical-domain score from 0 to 8; higher values indicate greater adherence.[3] The corresponding adherence proportions range from 0 to 1. No validated minimally important difference is established for these exploratory domain scores.

Table S6. Adjusted AMSTAR-2 Critical- and Noncritical-Domain Analyses

| **Outcome** | **Model** | **AI-assisted reviews, n** | **Non-AI reviews, n** | **Difference, AI minus non-AI (95% CI)** | **P value** |
| --- | --- | --- | --- | --- | --- |
| Critical-domain score, 5 items | Adjusted | 77 | 154 | −0.58 (−0.96 to −0.19) | 0.004 |
| Noncritical-domain score, 8 items | Adjusted | 77 | 154 | −1.06 (−1.53 to −0.58) | <0.001 |
| Critical-domain adherence proportion | Adjusted | 77 | 154 | −11.6 percentage points (−19.2 to −3.9) | 0.004 |
| Noncritical-domain adherence proportion | Adjusted | 77 | 154 | −13.2 percentage points (−19.2 to −7.3) | <0.001 |
| Critical-domain score, 5 items | Matched-set fixed-effects adjusted | 77 | 154 | −0.57 (−0.95 to −0.19) | 0.004 |
| Noncritical-domain score, 8 items | Matched-set fixed-effects adjusted | 77 | 154 | −1.16 (−1.61 to −0.71) | <0.001 |
| Critical-domain adherence proportion | Matched-set fixed-effects adjusted | 77 | 154 | −11.3 percentage points (−18.9 to −3.7) | 0.004 |
| Noncritical-domain adherence proportion | Matched-set fixed-effects adjusted | 77 | 154 | −14.5 percentage points (−20.1 to −8.9) | <0.001 |

Note. Critical-domain score included AMSTAR-2 items 2, 4, 7, 9, and 13; noncritical-domain score included items 1, 3, 5, 6, 8, 10, 14, and 16.[3] Higher scores indicate greater adherence; negative differences indicate lower adherence among AI-assisted reviews. Adjusted models included journal impact tier, funding reporting, review-group affiliation, review-team size, and review-question type. Matched-set fixed-effects models additionally included matched-set indicators. No validated minimally important difference is established for these exploratory domain outcomes.

Table S7. AMSTAR-2 Critical- and Noncritical-Domain Analyses Among Reviews with Meta-Analysis

| **Outcome** | **Model** | **AI n** | **Comparator n** | **Difference, AI minus comparator (95% CI)** | **P value** |
| --- | --- | --- | --- | --- | --- |
| Critical-domain score, 7 items | Adjusted | 31 | 86 | −1.09 (−1.72 to −0.46) | 0.001 |
| Noncritical-domain score, 9 items | Adjusted | 31 | 86 | −1.62 (−2.24 to −0.99) | <0.001 |
| Critical-domain adherence proportion | Adjusted | 31 | 86 | −15.6 percentage points (−24.6 to −6.6) | 0.001 |
| Noncritical-domain adherence proportion | Adjusted | 31 | 86 | −18.0 percentage points (−24.9 to −11.0) | <0.001 |
| Critical-domain score, 7 items | Matched-set fixed-effects adjusted | 31 | 86 | −1.16 (−2.12 to −0.19) | 0.022 |
| Noncritical-domain score, 9 items | Matched-set fixed-effects adjusted | 31 | 86 | −1.40 (−2.15 to −0.65) | 0.001 |
| Critical-domain adherence proportion | Matched-set fixed-effects adjusted | 31 | 86 | −16.5 percentage points (−30.3 to −2.8) | 0.022 |
| Noncritical-domain adherence proportion | Matched-set fixed-effects adjusted | 31 | 86 | −15.6 percentage points (−23.9 to −7.2) | 0.001 |

Note. The 7-item critical-domain score ranges from 0 to 7 and includes items 2, 4, 7, 9, 11, 13, and 15; the 9-item noncritical-domain score ranges from 0 to 9 and includes items 1, 3, 5, 6, 8, 10, 12, 14, and 16.[3] Higher values indicate greater adherence; negative differences indicate lower adherence among AI-assisted reviews. No validated minimally important difference is established for these exploratory domain outcomes.

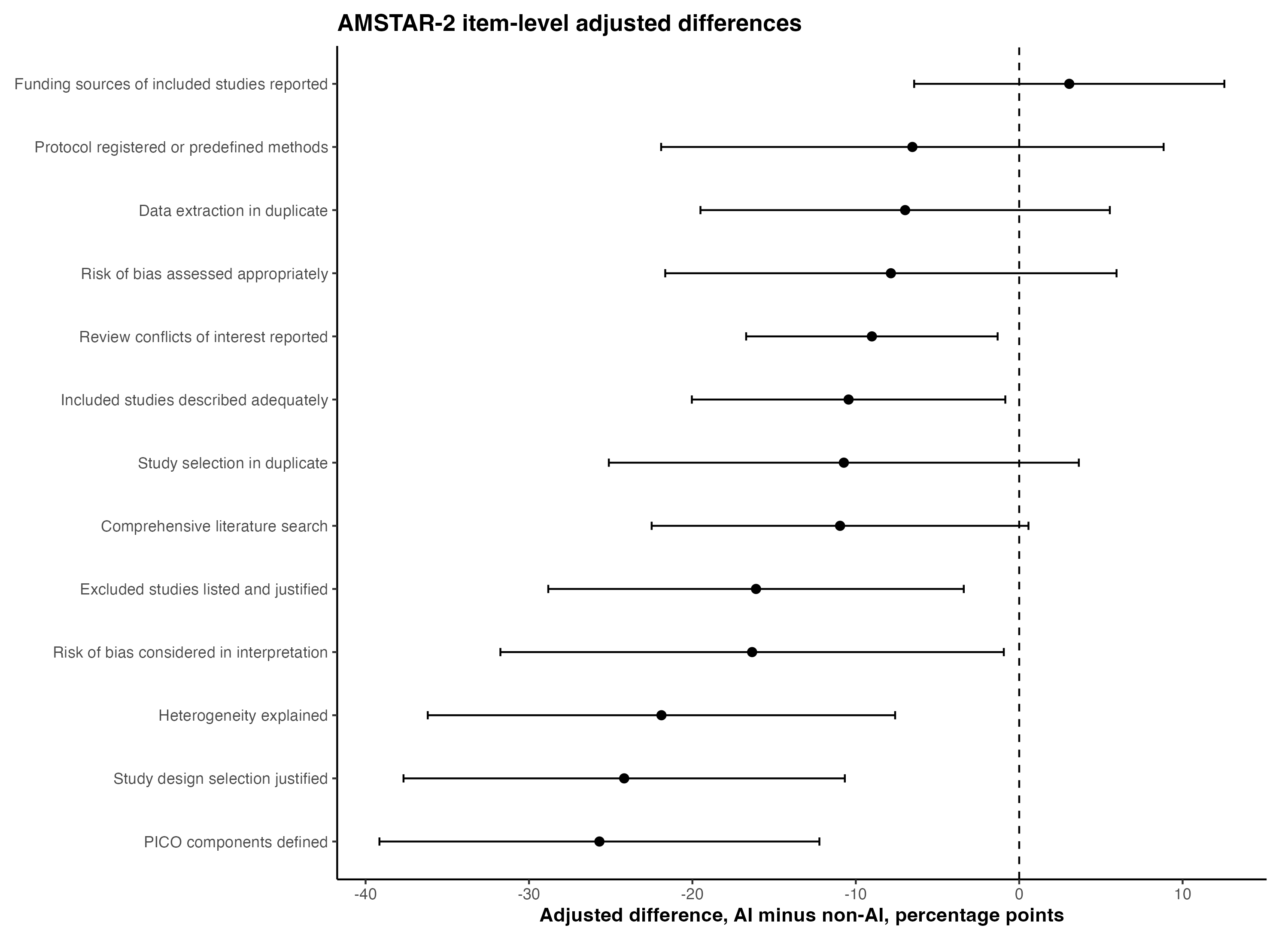

Figure S3. AMSTAR-2 Item-Level Adjusted Differences

Legend. Points show adjusted percentage-point differences in full item adherence (AI-assisted minus non-AI); horizontal lines show 95% confidence intervals, and the vertical dashed line indicates no difference. Negative values indicate lower adherence among AI-assisted reviews. A full “Yes” rating was classified as adherent; “Partial Yes” and “No” were not.[3] No validated minimally important difference is established for item-level contrasts. AI denotes artificial intelligence.

**Table S8. Item-Level AMSTAR-2 Adherence in the Modified 13-Item Assessment**

| **AMSTAR-2 item** | **Domain type** | **Reviews with no reported AI use (n=154), n (%)** | **AI-assisted reviews (n=77), n (%)** | **Adjusted difference, percentage points (95% CI)** | **P value** | **Matched-set fixed-effects adjusted difference, percentage points (95% CI)** | **P value** |
| --- | --- | --- | --- | --- | --- | --- | --- |
| AM1. PICO components defined | Noncritical | 140 (90.9) | 51 (66.2) | −25.7 (−39.2 to −12.2) | <0.001 | −24.1 (−37.4 to −10.8) | <0.001 |
| AM2. Protocol registered or methods predefined | Critical | 98 (63.6) | 42 (54.5) | −6.5 (−21.9 to 8.8) | 0.398 | −8.2 (−23.4 to 7.1) | 0.288 |
| AM3. Selection of study designs justified | Noncritical | 133 (86.4) | 49 (63.6) | −24.2 (−37.7 to −10.7) | <0.001 | −23.8 (−38.4 to −9.2) | 0.002 |
| AM4. Comprehensive literature search | Critical | 142 (92.2) | 62 (80.5) | −11.0 (−22.5 to 0.6) | 0.062 | −13.0 (−24.6 to −1.3) | 0.030 |
| AM5. Study selection performed in duplicate | Noncritical | 135 (87.7) | 53 (68.8) | −10.7 (−25.1 to 3.6) | 0.141 | −12.2 (−27.0 to 2.6) | 0.105 |
| AM6. Data extraction performed in duplicate | Noncritical | 120 (77.9) | 50 (64.9) | −7.0 (−19.5 to 5.5) | 0.269 | −9.1 (−20.8 to 2.6) | 0.124 |
| AM7. Excluded studies listed and exclusions justified | Critical | 44 (28.6) | 12 (15.6) | −16.1 (−28.8 to −3.4) | 0.014 | −14.4 (−28.2 to −0.6) | 0.041 |
| AM8. Included studies described adequately | Noncritical | 149 (96.8) | 65 (84.4) | −10.4 (−20.0 to −0.8) | 0.033 | −12.5 (−22.3 to −2.6) | 0.015 |
| AM9. Risk of bias assessed appropriately | Critical | 121 (78.6) | 45 (58.4) | −7.9 (−21.7 to 6.0) | 0.260 | −7.3 (−22.5 to 8.0) | 0.342 |
| AM10. Funding sources of included studies reported | Noncritical | 17 (11.0) | 9 (11.7) | 3.1 (−6.4 to 12.6) | 0.521 | 0.6 (−10.2 to 11.4) | 0.906 |
| AM13. Risk of bias considered when interpreting results | Critical | 109 (70.8) | 35 (45.5) | −16.3 (−31.7 to −0.9) | 0.038 | −13.9 (−29.9 to 2.2) | 0.089 |
| AM14. Heterogeneity satisfactorily explained and discussed | Noncritical | 124 (80.5) | 42 (54.5) | −21.9 (−36.2 to −7.6) | 0.003 | −25.3 (−40.3 to −10.3) | 0.001 |
| AM16. Review authors’ conflicts of interest reported | Noncritical | 151 (98.1) | 69 (89.6) | −9.0 (−16.7 to −1.3) | 0.022 | −9.8 (−17.7 to −1.8) | 0.017 |

**Note.** Only a “Yes” response was classified as adherent.[3] AMSTAR-2 item ratings were available for all 231 reviews. Adjusted differences were estimated using linear probability models with CR2 cluster-robust standard errors clustered by matched set and Satterthwaite small-sample corrections, and are expressed as AI-assisted reviews minus reviews with no reported AI use. Negative differences indicate lower adherence among AI-assisted reviews. Models adjusted for journal impact tier, funding reporting, formal review-group affiliation, review-team size, and review-question type. Matched-set fixed-effects models additionally included matched-set indicators. Items 11, 12, and 15 were excluded because they apply only to reviews with meta-analysis. Item-level analyses were exploratory, were not adjusted for multiple comparisons, and have no established minimally important difference. AI denotes artificial intelligence; AMSTAR-2, A Measurement Tool to Assess Systematic Reviews, version 2; CI, confidence interval; PICO, population, intervention, comparator, and outcome.

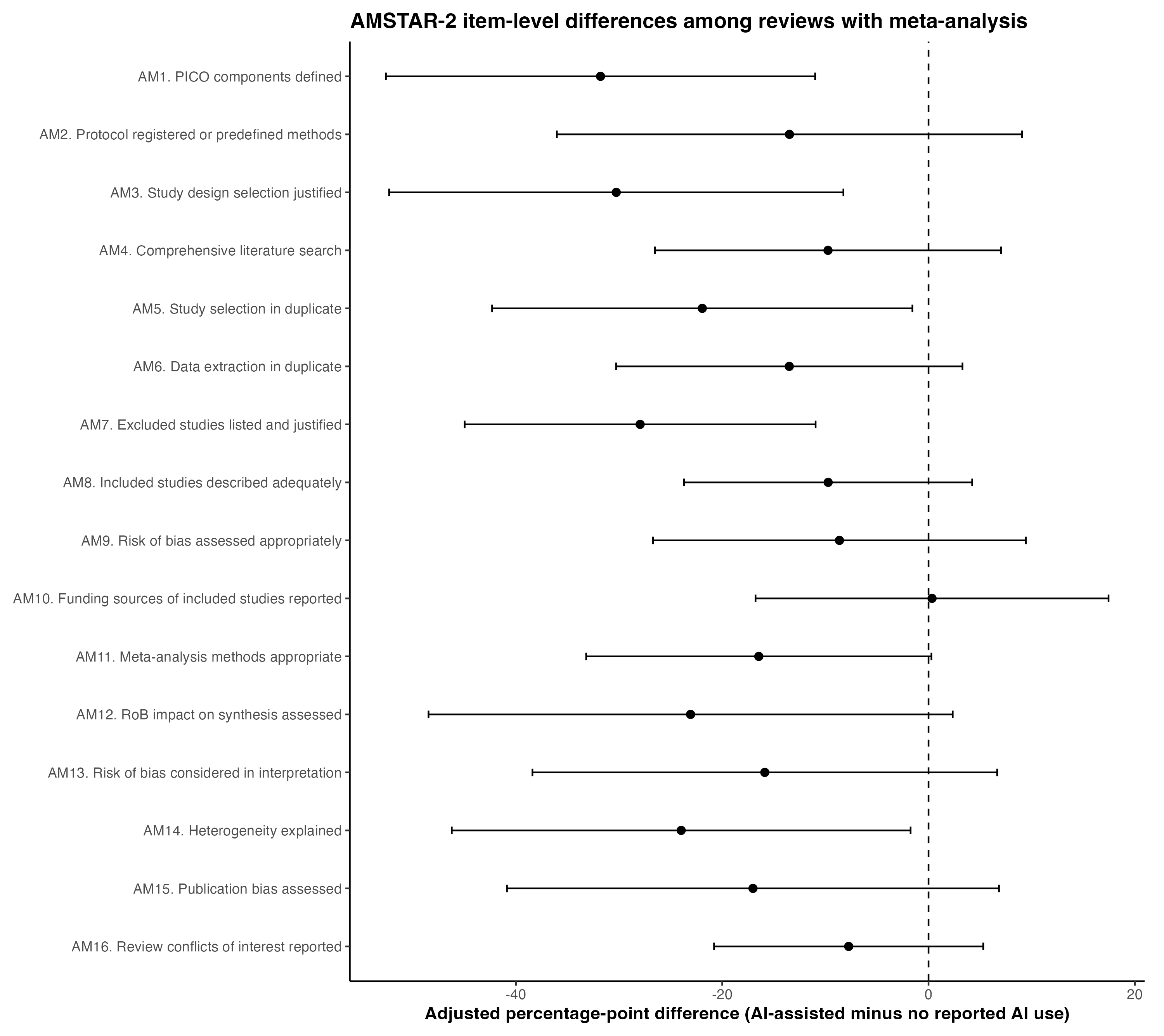

Figure S4. AMSTAR-2 Item-Level Adjusted Differences Among Reviews with Meta-Analysis

Legend. Points show adjusted percentage-point differences in full item adherence (AI-assisted minus non-AI); horizontal lines show 95% confidence intervals, and the vertical dashed line indicates no difference. Negative values indicate lower adherence among AI-assisted reviews. A “Yes” rating was classified as adherent.[3] This exploratory analysis was restricted to reviews with meta-analysis and has no established minimally important difference. AI denotes artificial intelligence.

**Table S9. Item-Level AMSTAR-2 Adherence Among Reviews with Meta-Analysis**

| **AMSTAR-2 item** | **Domain type** | **Item type** | **Reviews with no reported AI use (n=86), n (%)** | **AI-assisted reviews (n=31), n (%)** | **Adjusted difference, percentage points (95% CI)** | **P value** |
| --- | --- | --- | --- | --- | --- | --- |
| **AM1. PICO components defined** | Noncritical | General item | 80 (93.0) | 20 (64.5) | −31.8 (−52.6 to −11.0) | 0.004 |
| **AM2. Protocol registered or methods predefined** | Critical | General item | 65 (75.6) | 19 (61.3) | −13.5 (−36.0 to 9.1) | 0.231 |
| **AM3. Selection of study designs justified** | Noncritical | General item | 78 (90.7) | 20 (64.5) | −30.3 (−52.3 to −8.3) | 0.009 |
| **AM4. Comprehensive literature search** | Critical | General item | 81 (94.2) | 27 (87.1) | −9.7 (−26.5 to 7.0) | 0.244 |
| **AM5. Study selection performed in duplicate** | Noncritical | General item | 83 (96.5) | 21 (67.7) | −21.9 (−42.3 to −1.6) | 0.036 |
| **AM6. Data extraction performed in duplicate** | Noncritical | General item | 77 (89.5) | 22 (71.0) | −13.5 (−30.3 to 3.3) | 0.111 |
| **AM7. Excluded studies listed and exclusions justified** | Critical | General item | 25 (29.1) | 4 (12.9) | −28.0 (−45.0 to −10.9) | 0.002 |
| **AM8. Included studies described adequately** | Noncritical | General item | 84 (97.7) | 28 (90.3) | −9.7 (−23.7 to 4.2) | 0.164 |
| **AM9. Risk of bias assessed appropriately** | Critical | General item | 80 (93.0) | 24 (77.4) | −8.6 (−26.7 to 9.4) | 0.336 |
| **AM10. Funding sources of included studies reported** | Noncritical | General item | 13 (15.1) | 5 (16.1) | 0.3 (−16.8 to 17.5) | 0.968 |
| **AM11. Meta-analysis methods appropriate** | Critical | Meta-analysis–specific item | 82 (95.3) | 23 (74.2) | −16.5 (−33.2 to 0.3) | 0.054 |
| **AM12. Effect of risk of bias on synthesis assessed** | Noncritical | Meta-analysis–specific item | 61 (70.9) | 13 (41.9) | −23.1 (−48.5 to 2.3) | 0.073 |
| **AM13. Risk of bias considered when interpreting results** | Critical | General item | 68 (79.1) | 17 (54.8) | −15.9 (−38.4 to 6.7) | 0.160 |
| **AM14. Heterogeneity satisfactorily explained and discussed** | Noncritical | General item | 83 (96.5) | 22 (71.0) | −24.0 (−46.2 to −1.7) | 0.036 |
| **AM15. Publication bias assessed** | Critical | Meta-analysis–specific item | 63 (73.3) | 17 (54.8) | −17.0 (−40.9 to 6.8) | 0.155 |
| **AM16. Review authors’ conflicts of interest reported** | Noncritical | General item | 85 (98.8) | 29 (93.5) | −7.7 (−20.8 to 5.3) | 0.234 |

**Note.** Only a full “Yes” response was classified as adherent.[3] The analysis was restricted to reviews with meta-analysis, and AMSTAR-2 item ratings were available for all 117 reviews. Adjusted differences were estimated using linear probability models with CR2 cluster-robust standard errors clustered by matched set and Satterthwaite small-sample corrections, and are expressed as AI-assisted reviews minus reviews with no reported AI use. Negative differences indicate lower adherence among AI-assisted reviews. Models adjusted for journal impact tier, funding reporting, formal review-group affiliation, review-team size, and review-question type. Items 11, 12, and 15 apply specifically to reviews with meta-analysis. Item-level analyses were exploratory, were not adjusted for multiple comparisons, and have no established minimally important difference. AI denotes artificial intelligence; AMSTAR-2, A Measurement Tool to Assess Systematic Reviews, version 2; CI, confidence interval; PICO, population, intervention, comparator, and outcome.

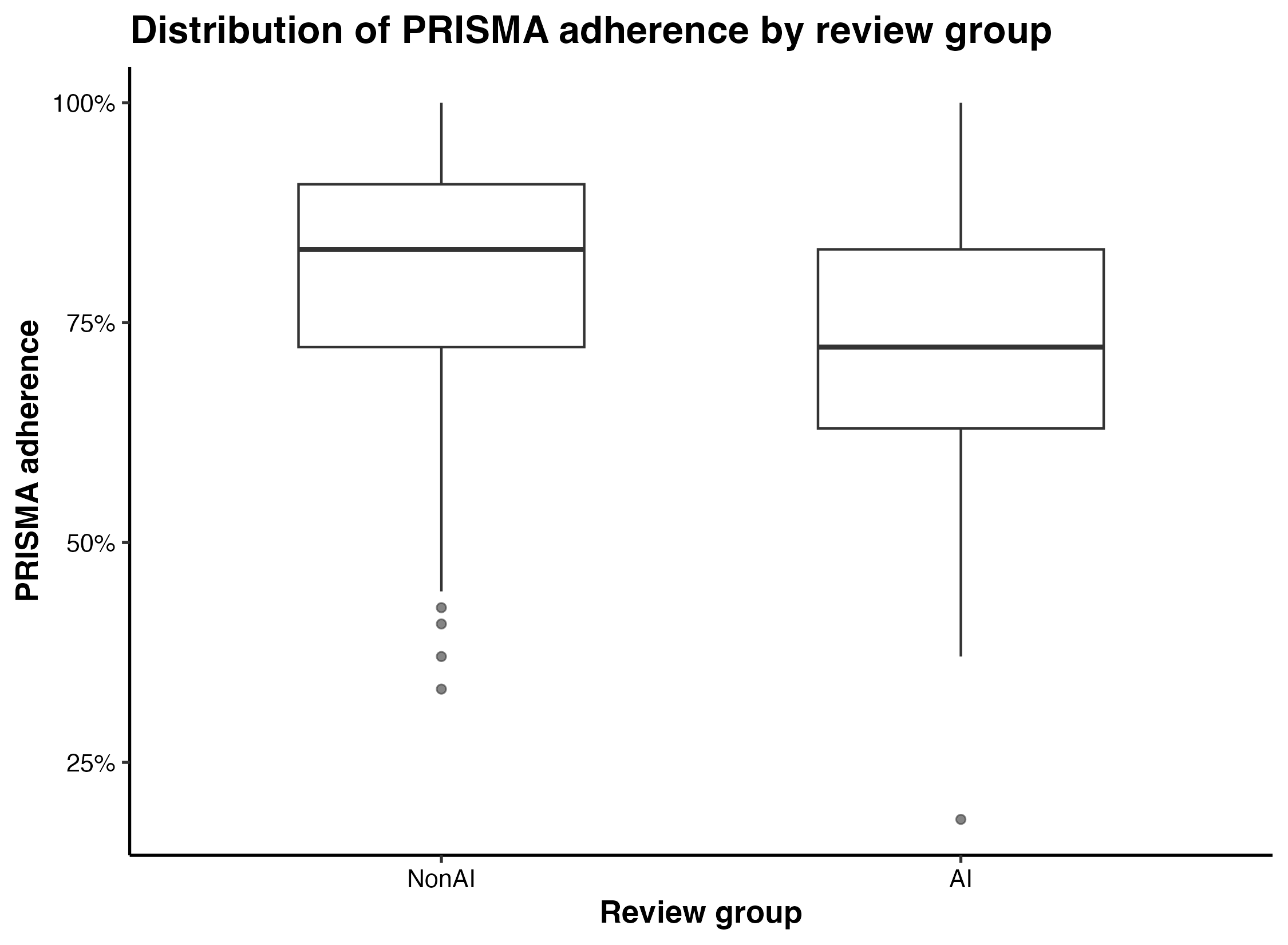

**Figure S5. Distribution of PRISMA 2020 Adherence by Review Group**

Legend. Boxes show the interquartile range, horizontal lines show medians, whiskers extend to 1.5 times the interquartile range, and points indicate outliers. PRISMA adherence ranges from 0 to 1 and is displayed as a percentage; higher values indicate more complete reporting.[4] Partial reporting received half credit. No validated minimally important difference is established for this adherence measure. AI denotes artificial intelligence; PRISMA, Preferred Reporting Items for Systematic Reviews and Meta-Analyses.

Table S10. PRISMA 2020 Domain-Level Analyses

| **PRISMA domain** | **Items** | **Non-AI mean adherence, mean (SD)** | **AI mean adherence, mean (SD)** | **Adjusted difference, AI minus non-AI** | **P value** | **Matched-set FE difference** | **FE P value** |
| --- | --- | --- | --- | --- | --- | --- | --- |
| Overall PRISMA adherence | 27 | 0.79 (0.15) | 0.71 (0.17) | −4.7 pp (−8.9 to −0.6) | 0.027 | −6.4 pp (−10.7 to −2.1) | 0.004 |
| Title/abstract | 2 | 0.95 (0.13) | 0.86 (0.24) | −7.20 pp (−13.88 to −0.51) | 0.035 | −7.38 pp (−14.06 to −0.70) | 0.031 |
| Introduction | 2 | 0.99 (0.06) | 0.99 (0.07) | −0.51 pp (−2.46 to 1.44) | 0.605 | 0.22 pp (−2.19 to 2.64) | 0.852 |
| Methods | 11 | 0.75 (0.20) | 0.68 (0.22) | −2.83 pp (−8.15 to 2.49) | 0.291 | −5.27 pp (−10.70 to 0.15) | 0.057 |
| Results | 7 | 0.71 (0.22) | 0.59 (0.25) | −7.99 pp (−14.56 to −1.43) | 0.018 | −11.10 pp (−17.80 to −4.39) | 0.002 |
| Discussion | 1 | 0.98 (0.13) | 0.95 (0.17) | −4.46 pp (−9.13 to 0.21) | 0.061 | −2.70 pp (−7.46 to 2.07) | 0.261 |
| Other information | 4 | 0.82 (0.22) | 0.74 (0.21) | −5.07 pp (−11.27 to 1.13) | 0.107 | −5.11 pp (−11.45 to 1.24) | 0.112 |

Note. Adherence proportions range from 0 to 1, with higher values indicating more complete reporting.[4] Partial reporting received half credit. Differences are AI-assisted minus non-AI and are reported in percentage points; negative values indicate lower adherence among AI-assisted reviews. The overall row reflects the final comparative models, and the domain rows are exploratory decompositions. No validated minimally important difference is established. FE denotes fixed effects, pp, percentage points.

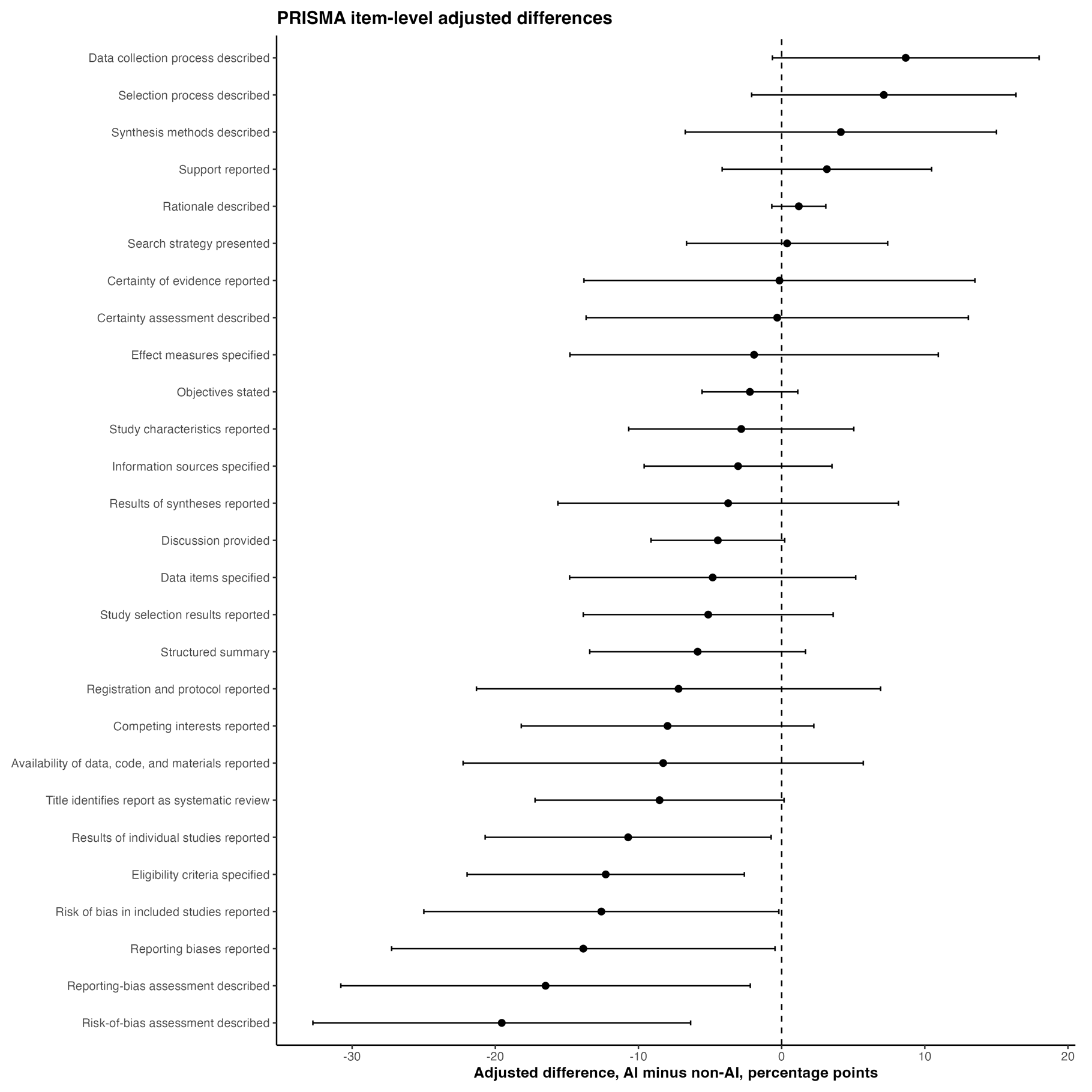

Figure S6. PRISMA 2020 Item-Level Adjusted Differences

Legend. Points show adjusted percentage-point differences in numeric item scores (AI-assisted minus non-AI); horizontal lines show 95% confidence intervals, and the vertical dashed line indicates no difference. Numeric scores were 1 for complete, 0.5 for partial, and 0 for absent reporting.[4] Negative values indicate less complete reporting among AI-assisted reviews. Item-level analyses were exploratory and have no established minimally important difference. AI denotes artificial intelligence; PRISMA, Preferred Reporting Items for Systematic Reviews and Meta-Analyses.

Table S11. Selected PRISMA 2020 Item-Level Contributors Using the Numeric Item Score

| **Item** | **Domain** | **Non-AI mean score (SD)** | **AI mean score (SD)** | **Adjusted score difference, AI minus non-AI** | **P value** |
| --- | --- | --- | --- | --- | --- |
| PR14. Reporting-bias assessment described | Methods | 0.49 (0.49) | 0.27 (0.44) | −16.5 pp (−30.8 to −2.2) | 0.025 |
| PR18. Risk of bias in included studies reported | Results | 0.75 (0.43) | 0.46 (0.50) | −19.5 pp (−32.7 to −6.4) | 0.004 |
| PR27. Availability of data, code, and materials reported | Other information | 0.72 (0.43) | 0.64 (0.48) | −8.3 pp (−22.3 to 5.7) | 0.241 |
| PR5. Eligibility criteria specified | Methods | 0.96 (0.15) | 0.84 (0.34) | −12.3 pp (−22.0 to −2.6) | 0.014 |
| PR19. Results of individual studies reported | Results | 0.90 (0.27) | 0.71 (0.42) | −10.7 pp (−20.7 to −0.7) | 0.036 |
| PR11. Study risk-of-bias assessment described | Methods | 0.79 (0.40) | 0.55 (0.49) | −12.6 pp (−25.0 to −0.2) | 0.047 |
| PR26. Competing interests reported | Other information | 0.95 (0.21) | 0.86 (0.35) | −8.0 pp (−18.2 to 2.2) | 0.124 |
| PR21. Reporting biases reported | Results | 0.42 (0.48) | 0.27 (0.44) | −13.9 pp (−27.2 to −0.5) | 0.043 |
| PR23. Discussion provided | Discussion | 0.98 (0.13) | 0.95 (0.17) | −4.5 pp (−9.1 to 0.2) | 0.061 |
| PR24. Registration and protocol reported | Other information | 0.69 (0.46) | 0.58 (0.48) | −7.2 pp (−21.3 to 6.9) | 0.311 |
| PR25. Support reported | Other information | 0.91 (0.28) | 0.90 (0.28) | 3.2 pp (−4.2 to 10.5) | 0.391 |

Note. Numeric item scores range from 0 to 1: complete reporting=1, partial reporting=0.5, and absent reporting=0.[4] Higher scores indicate more complete reporting. Adjusted differences are AI-assisted minus non-AI and are reported in percentage points; negative values indicate lower reporting completeness among AI-assisted reviews. Item-level analyses were exploratory, were not adjusted for multiple comparisons, and have no established minimally important difference. pp denotes percentage points.

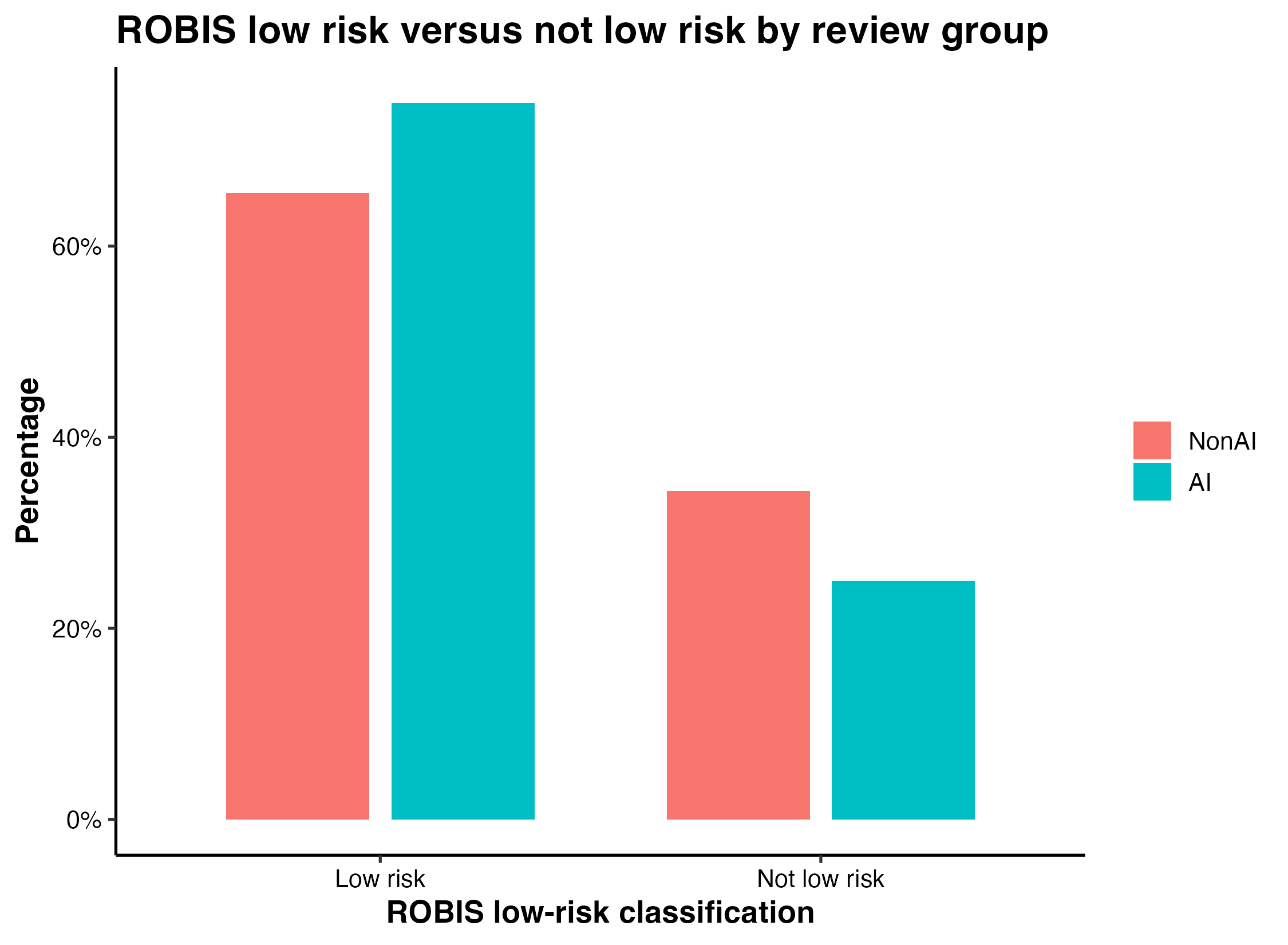

Figure S7. ROBIS Overall Low-Risk Classification by Review Group

Legend. Bars show the percentage of assessable reviews classified as low risk or not low risk. Low risk is the favorable outcome; high and unclear judgments were classified as not low risk, and not-applicable or missing judgments were excluded.[5] One AI-assisted review had a not-applicable overall judgment. ROBIS is categorical and has no minimally important difference. AI denotes artificial intelligence; ROBIS, Risk of Bias in Systematic Reviews.

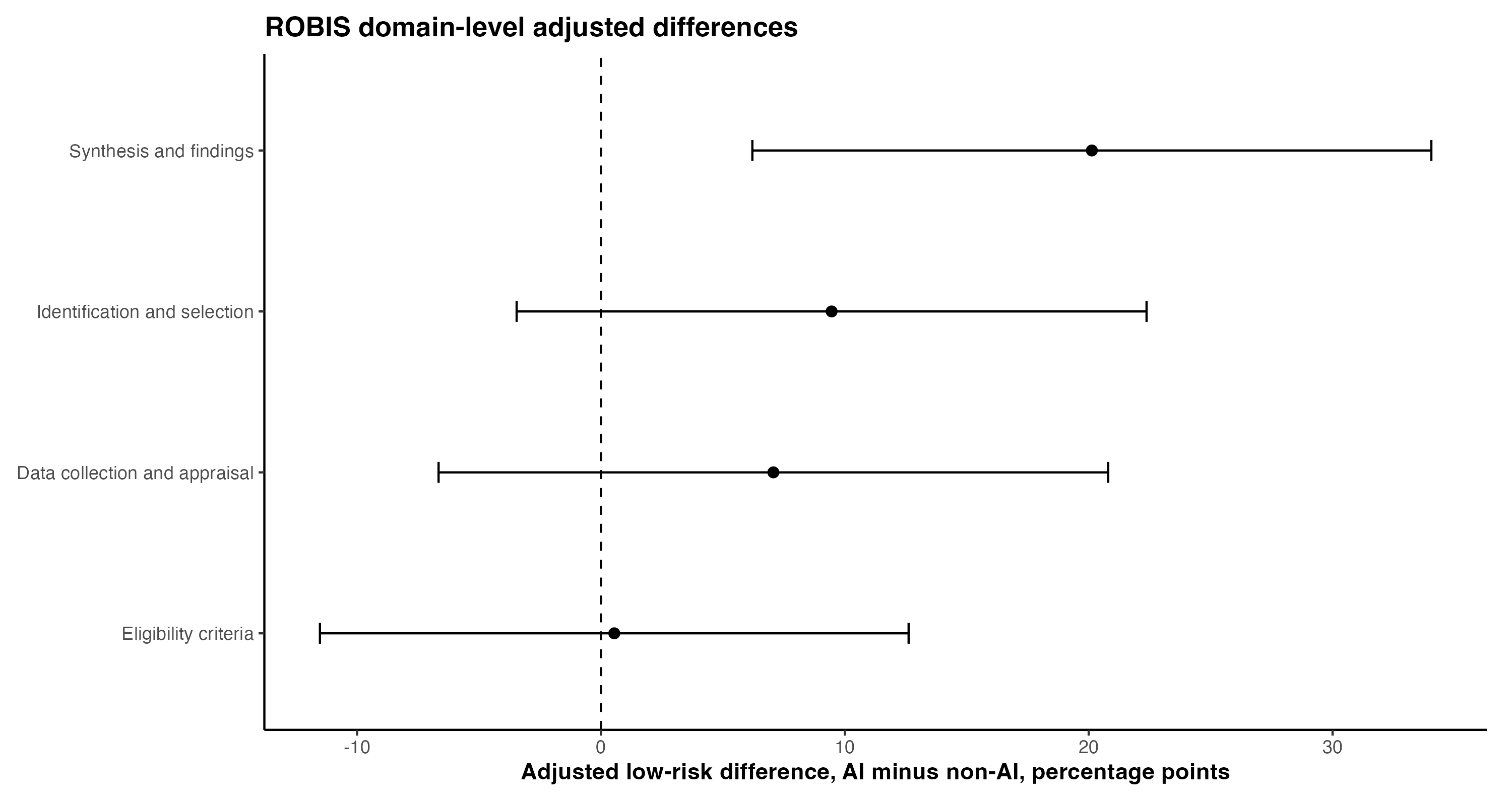

Figure S8. ROBIS Domain-Level Adjusted Differences in Low-Risk Ratings

Legend. Points show adjusted percentage-point differences in the proportion rated low risk (AI-assisted minus non-AI); horizontal lines show 95% confidence intervals and the vertical dashed line indicates no difference. Positive values indicate more favorable low-risk ratings among AI-assisted reviews. High and unclear judgments were classified as not low risk; not-applicable and missing judgments were excluded.[5] Domain-level analyses were exploratory and have no established minimally important difference.

Table S12. ROBIS Domain-Level Analyses: Low Risk of Bias in AI-Assisted and Non-AI Reviews

| **ROBIS component** | **Non-AI low risk, n/N (%)** | **AI-assisted low risk, n/N (%)** | **Unadjusted difference** | **Adjusted difference, AI minus non-AI (95% CI)** | **Adjusted P value** | **Matched-set FE difference (95% CI)** | **FE P value** |
| --- | --- | --- | --- | --- | --- | --- | --- |
| Domain 1: Eligibility criteria | 134/154 (87.0%) | 66/76 (86.8%) | −0.2 pp | 0.5 pp (−11.5 to 12.6) | 0.928 | −1.1 pp (−13.6 to 11.4) | 0.862 |
| Domain 2: Identification and selection | 120/153 (78.4%) | 63/76 (82.9%) | 4.5 pp | 9.5 pp (−3.5 to 22.4) | 0.148 | 10.5 pp (−3.4 to 24.4) | 0.134 |
| Domain 3: Data collection and appraisal | 107/154 (69.5%) | 50/76 (65.8%) | −3.7 pp | 7.1 pp (−6.7 to 20.8) | 0.307 | 8.1 pp (−6.2 to 22.4) | 0.259 |
| Domain 4: Synthesis and findings | 100/154 (64.9%) | 60/76 (78.9%) | 14.0 pp | 20.1 pp (6.2 to 34.0) | 0.005 | 18.3 pp (2.1 to 34.4) | 0.027 |

Note. Values represent the proportion of assessable reviews rated low risk.[5] High and unclear judgments were classified as not low risk; not applicable and missing judgments were excluded. Positive percentage-point differences indicate a higher proportion rated low risk among AI-assisted reviews. Domain-level analyses were exploratory and have no established minimally important difference. FE denotes fixed effects, pp, percentage points.

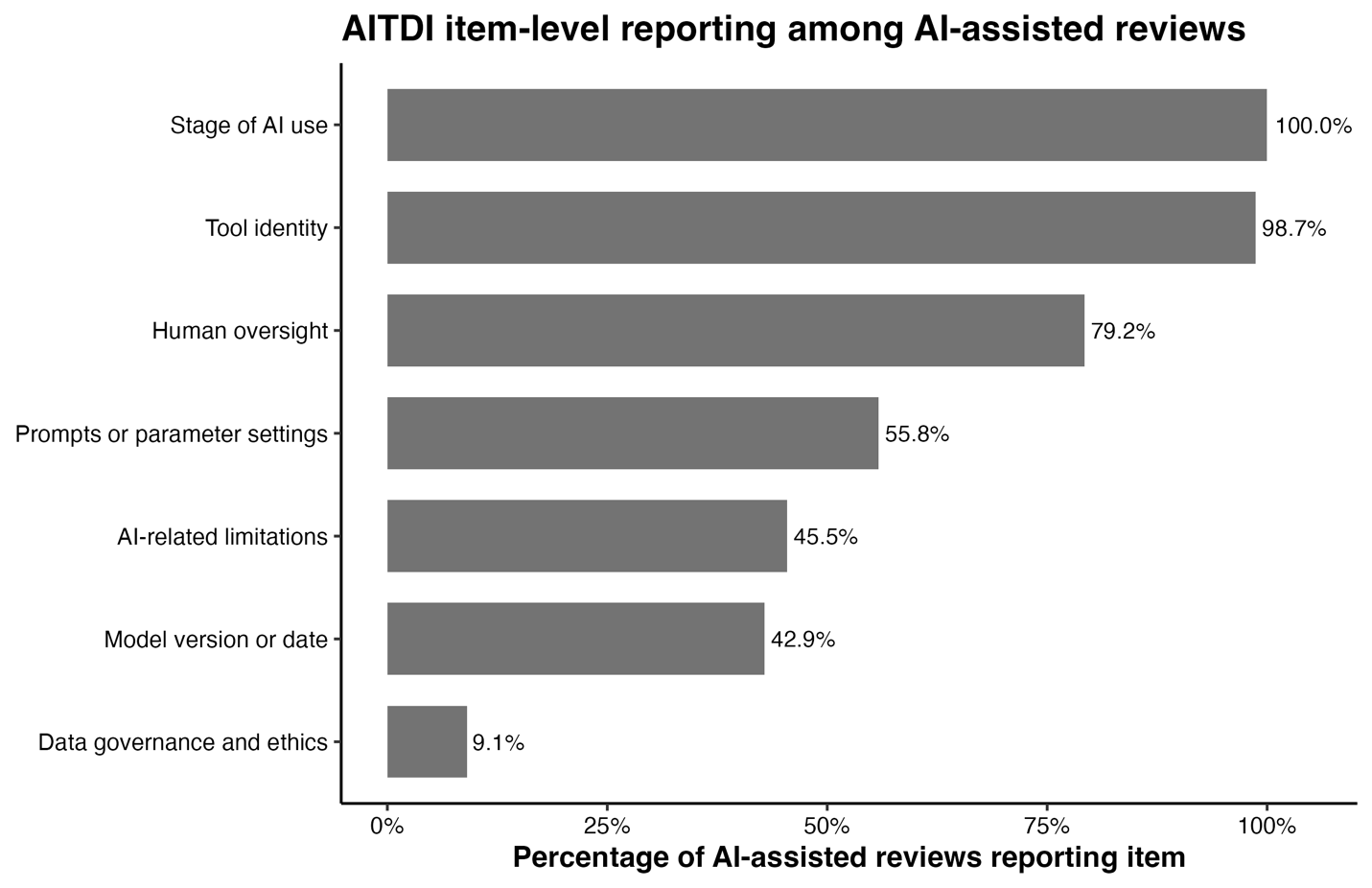

Figure S9. AITDI Item-Level Reporting Among AI-Assisted Reviews

Legend. Bars show the percentage of AI-assisted reviews reporting each transparency item. AITDI contains six scored items with a total range of 0 to 6, with higher scores indicating more complete AI-use reporting; AI-related limitations are summarized as an additional nonscored item.[1] AITDI is preliminary and has no validated minimally important difference. AI denotes artificial intelligence; AITDI, Artificial Intelligence Transparency and Disclosure Index.

Table S13. AITDI Item-Level Reporting Among AI-Assisted Reviews

| **AITDI item** | **Domain** | **Reported, n/N (%)** | **Interpretation** |
| --- | --- | --- | --- |
| Data governance and ethics considerations | Data governance/ethics | 7/77 (9.1%) | Rarely reported |
| Model version or date | Transparency/reporting | 33/77 (42.9%) | Infrequently reported |
| AI-related limitations | Limitations | 35/77 (45.5%) | Infrequently reported |
| Parameter settings or prompts | Transparency/reporting | 43/77 (55.8%) | Moderately reported |
| Human oversight | Transparency/reporting | 61/77 (79.2%) | Commonly reported |
| Tool identity, specific AI system used | Transparency/reporting | 76/77 (98.7%) | Nearly universally reported |
| Stage of use, review stages involving AI | Transparency/reporting | 77/77 (100%) | Universally reported |

Note. AITDI contains six scored binary items; AI-related limitations are shown as an additional nonscored item.[1] Items are ordered from lowest to highest reporting frequency. Higher reporting frequency indicates more complete disclosure but does not establish methodological quality. AITDI was not assessed among non-AI reviews and has no validated minimally important difference. AI denotes artificial intelligence; AITDI, Artificial Intelligence Transparency and Disclosure Index.

Table S14. AITDI Total Scores Among AI-Assisted Reviews by Selected Study and AI-Use Characteristics

| **Characteristic** | **Level** | **AI reviews, n** | **Mean (SD)** | **Median [IQR]** | **Range** |
| --- | --- | --- | --- | --- | --- |
| Overall | All AI-assisted reviews | 77 | 3.86 (1.06) | 4 [3–5] | 2–6 |
| Journal impact tier | Q1 | 52 | 3.73 (1.09) | 3 [3–5] | 2–6 |
| Journal impact tier | Q2 | 21 | 4.19 (0.98) | 4 [4–5] | 2–6 |
| Journal impact tier | Unknown | 4 | 3.75 (0.96) | 3.5 [3–4.2] | 3–5 |
| Funding reporting | Not reported | 7 | 3.43 (0.98) | 3 [3–4] | 2–5 |
| Funding reporting | Reported | 70 | 3.90 (1.07) | 4 [3–5] | 2–6 |
| Review question type | Intervention | 15 | 4.00 (1.41) | 4 [3–5] | 2–6 |
| Review question type | Diagnostic | 7 | 3.86 (0.69) | 4 [3.5–4] | 3–5 |
| Review question type | Etiology/Association | 7 | 4.29 (0.95) | 5 [3.5–5] | 3–5 |
| Review question type | Prognostic | 5 | 3.00 (0.71) | 3 [3–3] | 2–4 |
| Review question type | Prevalence | 6 | 3.50 (0.55) | 3.5 [3–4] | 3–4 |
| Review question type | Qualitative | 6 | 4.83 (0.98) | 5 [5–5] | 3–6 |
| Review question type | Methodological | 20 | 3.90 (0.97) | 4 [3–5] | 2–5 |
| Review question type | Other/Mixed | 11 | 3.36 (0.92) | 3 [3–3.5] | 2–5 |
| Broad AI tool type | LLM plus screening/classification tool | 3 | 3.33 (0.58) | 3 [3–3.5] | 3–4 |
| Broad AI tool type | LLM-based tool only | 34 | 4.47 (0.90) | 5 [4–5] | 3–6 |
| Broad AI tool type | Other or mixed AI tool type | 5 | 3.60 (0.89) | 3 [3–4] | 3–5 |
| Broad AI tool type | Screening/classification tool only | 35 | 3.34 (0.97) | 3 [3–4] | 2–5 |
| AI stage of use | Data extraction only | 7 | 4.00 (1.15) | 4 [3–4.5] | 3–6 |
| AI stage of use | Multiple review stages | 11 | 4.27 (1.10) | 5 [3–5] | 3–6 |
| AI stage of use | Screening/selection only | 45 | 3.69 (1.06) | 4 [3–5] | 2–6 |
| AI stage of use | Search/retrieval only | 7 | 3.71 (0.95) | 3 [3–4.5] | 3–5 |
| AI stage of use | Synthesis/analysis only | 7 | 4.29 (0.95) | 5 [3.5–5] | 3–5 |

Note. AITDI total scores range from 0 to 6, with higher scores indicating more complete AI-use reporting.[1] Scores were summarized descriptively among AI-assisted reviews only; no formal hypothesis testing was performed. AITDI is preliminary and has no validated minimally important difference. Categories with fewer than 5 reviews included unknown journal impact tier and combined LLM plus screening/classification tools. LLM denotes large language model.

Table S15. Associations Between AITDI Total Score and Methodological or Reporting Quality Among AI-Assisted Reviews

| **Outcome** | **AITDI model** | **Estimate per 1-point higher AITDI (95% CI)** |
| --- | --- | --- |
| AMSTAR-2 13-item score | Simple | −0.16 (−0.72 to 0.40) |
| AMSTAR-2 13-item score | Adjusted | −0.10 (−0.64 to 0.43) |
| PRISMA adherence | Simple | −1.5 percentage points (−5.2 to 2.2) |
| PRISMA adherence | Adjusted | −1.3 percentage points (−4.6 to 2.1) |

Note. AITDI total scores range from 0 to 6, with higher scores indicating more complete AI-use reporting.[1] Estimates represent the difference associated with each 1-point higher AITDI score. PRISMA estimates are reported in percentage points. Analyses were restricted to AI-assisted reviews; adjusted models included selected review-level covariates. AITDI is preliminary and has no validated minimally important difference.

Table S16. Exploratory Timeliness and Dissemination Analyses

| **Outcome** | **Analysis population** | **AI-assisted reviews** | **Non-AI reviews** | **Adjusted estimate** | **P value** | **Additional notes** |
| --- | --- | --- | --- | --- | --- | --- |
| Receipt-to-publication interval | 193 reviews; 64 AI-assisted and 129 non-AI | Median 127.5 days (IQR 70.2–204.8) | Median 140 days (IQR 76–228) | Mean difference −2.6 days (95% CI −39.1 to 34.0) | 0.888 | Log-linear relative difference −0.1% (95% CI −22.5% to 28.7%); P=0.992 |
| 12-month citations | 208 reviews; 74 AI-assisted and 134 non-AI | Median 5 (IQR 1.2–12) | Median 3 (IQR 2–8.8) | Relative difference 11.1% (95% CI −10.7% to 38.2%) | 0.339 | Adjusted for journal impact tier and publication year |
| Altmetric Attention Score | 90 reviews; 34 AI-assisted and 56 non-AI | Median 6 (IQR 2.2–12.5) | Median 2 (IQR 1–8) | Relative difference 50.6% (95% CI −17.4% to 174.6%) | 0.176 | Substantial missingness and limited within-set overlap |

Note. Timeliness and dissemination analyses were exploratory. Receipt-to-publication interval reflects the journal receipt-to-publication process rather than the time required to conduct the systematic review. Relative differences for citation and Altmetric outcomes were derived from log(1+outcome) models and adjusted for journal impact tier and publication year. IQR denotes interquartile range.

Table S17. Matched-Set Paired Sensitivity Analysis for the AMSTAR-2 13-Item Score

| **Outcome** | **Analysis** | **Matched sets, n** | **Mean paired difference** | **SD** | **Median paired difference (IQR)** | **Range** | **95% CI** | **P value vs 0** | **P value vs margin −1.0** | **Noninferiority met** |
| --- | --- | --- | --- | --- | --- | --- | --- | --- | --- | --- |
| AMSTAR-2 13-item score | Paired matched-set analysis | 77 | −2.05 | 2.83 | −2.00 (−3.50 to 0.00) | −10.00 to 2.50 | −2.69 to −1.40 | <0.001 | 0.002 | No |

Note. For each matched set, the paired difference was the AI-assisted review’s modified AMSTAR-2 13-item score minus the mean score of its two matched non-AI comparators. The score ranges from 0 to 13, with higher scores indicating greater adherence.[3] The prespecified noninferiority margin was -1.0 point; noninferiority required the lower bound of the 95% confidence interval to exceed -1.0. No validated minimally important difference exists for the modified score. IQR denotes interquartile range; SD, standard deviation.

Table S18. Sensitivity Analysis Restricted to Standard Systematic Reviews

| **Outcome** | **Analysis population** | **Model** | **Reviews, n** | **Matched sets, n** | **AI reviews, n** | **Non-AI reviews, n** | **Mean difference, AI minus non-AI (95% CI)** | **P value** | **Noninferiority margin** | **Noninferiority met** |
| --- | --- | --- | --- | --- | --- | --- | --- | --- | --- | --- |
| AMSTAR-2 13-item score | Standard systematic reviews only | Adjusted | 227 | 77 | 74 | 153 | −1.75 (−2.47 to −1.03) | <0.001 | −1.0 | No |
| AMSTAR-2 13-item score | Standard systematic reviews only | Matched-set fixed-effects adjusted | 227 | 77 | 74 | 153 | −1.89 (−2.56 to −1.22) | <0.001 | −1.0 | No |

Note. The modified AMSTAR-2 13-item score ranges from 0 to 13, with higher scores indicating greater adherence.[3] The prespecified noninferiority margin was -1.0 point. Adjusted models included journal impact tier, funding reporting, review-group affiliation, review-team size, and review-question type; the fixed-effects model additionally included matched-set indicators. No validated minimally important difference exists for the modified score.

Table S19. Leave-One-Journal-Out Sensitivity Analysis for the AMSTAR-2 13-Item Score

| **Journal removed** | **Reviews from journal** | **Matched sets removed** | **Reviews removed** | **Remaining reviews** | **Estimate, AI minus non-AI** | **Shift from primary** | **P value** | **Noninferiority met** |
| --- | --- | --- | --- | --- | --- | --- | --- | --- |
| Journal of Medical Internet Research | 4 | 4 | 12 | 219 | −1.82 (−2.58 to −1.05) | −0.18 | <0.001 | No |
| International Journal of Molecular Sciences | 3 | 3 | 9 | 222 | −1.69 (−2.46 to −0.92) | −0.06 | <0.001 | No |
| Frontiers in Medicine | 3 | 3 | 9 | 222 | −1.70 (−2.44 to −0.95) | −0.06 | <0.001 | No |
| Cureus | 7 | 6 | 18 | 213 | −1.66 (−2.41 to −0.91) | −0.02 | <0.001 | No |
| Healthcare | 4 | 4 | 12 | 219 | −1.57 (−2.35 to −0.79) | +0.07 | <0.001 | No |
| PLOS ONE | 3 | 3 | 9 | 222 | −1.54 (−2.27 to −0.81) | +0.10 | <0.001 | No |

Note. Matched sets containing each journal that contributed at least 3 reviews were removed one at a time, and the adjusted modified AMSTAR-2 13-item model was refitted. The score ranges from 0 to 13, with higher scores indicating greater adherence.[3] Negative estimates indicate lower adherence among AI-assisted reviews. The noninferiority margin was -1.0 point; no validated minimally important difference exists for the modified score.

Table S20. Overlap-Weighted Sensitivity Analysis for the AMSTAR-2 13-Item Score

| **Component** | **Finding** |
| --- | --- |
| Analysis population | 231 reviews; 77 AI-assisted and 154 non-AI reviews; 77 matched sets |
| Outcome | AMSTAR-2 13-item score |
| Exposure | AI-assisted review vs non-AI review |
| Propensity score covariates | Q1 journal status, funding reporting, review team size |
| Propensity score range | 0.246 to 0.661 |
| Overlap weight range | 0.252 to 0.754 |
| Effective sample size, Non-AI | 151.1 |
| Effective sample size, AI | 76.2 |
| Largest unweighted SMD | Review team size, SMD 0.203 |
| Weighted SMDs after overlap weighting | All approximately 0 |
| Overlap-weighted estimate | −2.09 AMSTAR-2 points |
| 95% CI | −2.73 to −1.44 |
| P value | <0.001 |
| Noninferiority margin | −1.0 AMSTAR-2 point |
| Noninferiority met? | No |

Note. Propensity scores were estimated using Q1 journal status, funding reporting, and review-team size. AI-assisted reviews received 1 minus the propensity score and non-AI reviews received the propensity score. The modified AMSTAR-2 score ranges from 0 to 13, with higher scores indicating greater adherence.[3] The noninferiority margin was -1.0 point; no validated minimally important difference exists for the modified score. SMD denotes standardized mean difference.

Table S21. Balance Diagnostics for the Overlap-Weighted Analysis

| **Covariate** | **Level** | **Unweighted SMD** | **Overlap-weighted SMD** |
| --- | --- | --- | --- |
| Review team size | Continuous | 0.203 | ~0.000 |
| Journal Q1 status | Q2/lower/unknown | 0.099 | ~0.000 |
| Journal Q1 status | Q1 | −0.099 | ~0.000 |
| Funding reported | Not reported | 0.000 | ~0.000 |
| Funding reported | Reported | 0.000 | ~0.000 |

Note. Standardized mean differences compare AI-assisted and non-AI reviews. Values closer to 0 indicate better balance; an absolute SMD below 0.10 is commonly interpreted as acceptable balance. AI denotes artificial intelligence; SMD, standardized mean difference.

Table S22. Factors Associated With AMSTAR-2 and PRISMA Scores in AI-Assisted and Non-AI Reviews

| **Outcome** | **Predictor** | **AI-assisted reviews, estimate (95% CI)** | **Non-AI reviews, estimate (95% CI)** |
| --- | --- | --- | --- |
| AMSTAR-2 13-item score | Clinical non-intervention vs intervention | −0.70 (−2.27 to 0.86) | −0.78 (−1.56 to 0.00) |
| AMSTAR-2 13-item score | Other/methodological/qualitative vs intervention | −2.20 (−3.67 to −0.73) | −1.19 (−2.32 to −0.06) |
| AMSTAR-2 13-item score | Review-team size, per additional member | 0.16 (0.03 to 0.29) | −0.03 (−0.16 to 0.11) |
| AMSTAR-2 13-item score | Q1 vs Q2/lower/unknown | 0.38 (−0.79 to 1.55) | 0.17 (−0.72 to 1.05) |
| PRISMA adherence | Clinical non-intervention vs intervention | −0.008 (−0.107 to 0.091) | −0.072 (−0.125 to −0.018) |
| PRISMA adherence | Other/methodological/qualitative vs intervention | −0.144 (−0.237 to −0.051) | −0.122 (−0.185 to −0.058) |
| PRISMA adherence | Review-team size, per additional member | 0.012 (0.004 to 0.021) | 0.000 (−0.009 to 0.009) |
| PRISMA adherence | Q1 vs Q2/lower/unknown | −0.010 (−0.083 to 0.064) | 0.039 (−0.026 to 0.103) |

Note. Estimates are mutually adjusted for review-question type, review-team size, and Q1 journal status. Intervention reviews and journals classified as Q2, lower, or unknown were reference categories. AMSTAR-2 estimates are point differences on the 0-to-13 modified score; PRISMA estimates are proportions and may be multiplied by 100 to obtain percentage points. [3,4] Conventional model-based standard errors were used for AI-assisted reviews; non-AI models used CR2 standard errors clustered by matched set with Satterthwaite corrections. Analyses were exploratory and were not formal interaction tests. No validated minimally important difference is established for either measure.

Table S23. Model-Complexity Assessment for Adjusted Models

| **Model** | **N** | **Estimated parameters, excluding intercept** | **Observations per parameter** | **Overfitting concern** |
| --- | --- | --- | --- | --- |
| AMSTAR13 adjusted, full cohort | 231 | 15 | 15.4 | Low concern |
| PRISMA adjusted, full cohort | 231 | 15 | 15.4 | Low concern |
| ROBIS adjusted LPM, full cohort | 230 | 15 | 15.3 | Low concern |
| AMSTAR16 adjusted, meta-analysis only | 117 | 15 | 7.8 | Moderate concern |

Note. Adjusted models included 15 estimated parameters excluding the intercept. The full-cohort AMSTAR-2 13-item, PRISMA, and ROBIS models had approximately 15 observations per estimated parameter. The AMSTAR-2 16-item model was restricted to reviews with meta-analysis. This table is a descriptive model-complexity diagnostic rather than a formal test of overfitting. LPM denotes linear probability model.

### **Appendix 3. References for Included Systematic Reviews**

References 6 through 236 correspond to the included reviews listed in Table S2. For each review, the supplementary reference number equals the manuscript ID plus 5.

#### **AI-Assisted Reviews**

68. Stafford IG, Lai MN, Tan K. Automated oxygen delivery for preterm infants with respiratory dysfunction. DOI: 10.1002/14651858.CD013294.pub2.

69. Steele DW, Kanaan G, Caputo EL, et al. Treatment of Obsessive-Compulsive Disorder in Children and Youth: A Meta-Analysis. Pediatrics 2025;155:1-20. DOI: 10.1542/peds.2024-068992.

70. Talukdar R, Ajayan R, Gupta S, et al. Chronic Kidney Disease Prevalence in India: A Systematic Review and Meta-Analysis From Community-Based Representative Evidence Between 2011 to 2023. Nephrology (Carlton, Vic.) 2025;30:e14420. DOI: 10.1111/nep.14420.

71. Teo ZL, Jin L, Li S, et al. Federated machine learning in healthcare: A systematic review on clinical applications and technical architecture. Cell reports. Medicine 2024;5:101419. DOI: 10.1016/j.xcrm.2024.101419.

72. Teperikidis L, Boulmpou A, Amin G, et al. Cardiac Remodeling in Preeclampsia: A Large-Language-Model-Assisted Meta-Analysis and Meta-Regression. Journal of cardiovascular pharmacology 2025. DOI: 10.1097/FJC.0000000000001774.

73. Trivedi A, Jatana V, Sinn KJ. Early versus late administration of amino acids in preterm infants receiving parenteral nutrition. DOI: 10.1002/14651858.CD008771.pub3.

74. Urru AS, Geist M, Carlinger R, Bodrero E, Bruschettini M. Strategies for cessation of caffeine administration in preterm infants. DOI: 10.1002/14651858.CD015802.pub2.

#### **Reviews With No Reported AI Use**

### **Appendix 4. Reporting Checklist for Meta-Epidemiological Methodology Research**

| **Item and topic** | **Reporting requirement** | **Location and response** |
| --- | --- | --- |
| 1 Title | Identify the study design. | Title page; Abstract, Methods. The title identifies a matched meta-research study and the abstract specifies a matched-cohort design. The exact term “meta-epidemiological” is not used. |
| 2 Structured summary | Summarise aims, sources, methods, findings, limitations and implications. | Partially reported. Abstract, Background, Objective, Methods, Results, and Conclusions. Provides rationale, objectives, design, eligibility, matching, outcomes, models, findings and implications; registration is within Methods. Database names and search dates, and explicit methodological limitations, are not summarized in the abstract. |
| 3 Rationale | Explain the knowledge gap. | Introduction, paragraphs 1–3. Explains the knowledge gap and the complementary role of evaluating completed reviews alongside task-level AI evaluations. |
| 4 Objectives | State objectives and the tested hypothesis. | Introduction, final paragraph; Methods, Outcomes and Statistical Analysis. Comparative objectives are explicit; the noninferiority hypothesis is operationalized using the −1-point margin. |
| 5 Protocol | Identify an accessible protocol. | Methods, Study Design; Appendix 1, S1 and S12. Provides OSF DOI 10.17605/OSF.IO/Q5JRW and published protocol reference 25; deviations and clarifications are documented. |
| 6 Eligibility criteria | Define eligibility and explain restrictions. | Methods, Eligibility Criteria and Exposure Classification; Appendix 1, S2–S4. Defines eligible review types, language, health focus, publication period and substantive reported AI use; the reason for the 2023–2025 period is stated. |
| 7 Information sources | Identify sources and search dates. | Methods, Information Sources and Search Strategy and Comparator Identification and Selection; Appendix 1, S2 and S5; internal Appendix 2, Detailed Search Strategies. Database platforms, coverage and main search dates are provided. Partially reported for the two additional PubMed replacement searches: exact dates are not provided. |
| 8 Search | Provide a reproducible electronic search. | Partially reported. Internal Appendix 2, Search Strategies A–C, gives complete database strategies and limits. Appendix 1, S5, describes individualized comparator searches and exceptions, but the exact terms and result lists for two additional PubMed replacement searches were unavailable; those searches cannot be fully reproduced. |
| 9 Study selection methods | Describe selection and reviewer involvement. | Methods, Eligibility Criteria and Exposure Classification and Comparator Identification and Selection; Appendix 1, S3 and S5. Describes duplicate screening, adjudication, reproducible random sampling, replacement procedures and exceptions. |
| 10 Data collection | Describe extraction and verification. | Methods, Data Extraction and Appraisal; Appendix 1, S6. Describes independent duplicate extraction using piloted forms, reconciliation, adjudication and consistency checks. |
| 11 Data items | Define variables, assumptions and missing-data handling. | Methods, Eligibility Criteria and Exposure Classification, Outcomes, Covariates, and Statistical Analysis; Appendix 1, S4 and S6–S10. Defines exposure, scores, covariates, operational coding and available-case analysis without imputation. Partially reported: detailed item-level rules distinguishing complete from partial PRISMA reporting are not provided, as acknowledged in Discussion. |
| 12 Risk of bias methods | Describe relevant appraisal and analytical use. | Methods, Data Extraction and Appraisal, Outcomes, and Statistical Analysis; Appendix 1, S7 and S9. ROBIS is a study outcome; low-risk versus high/unclear coding and not-applicable or missing exclusions are specified. |
| 13 Summary measures | Define estimates and their direction. | Methods, Statistical Analysis; Appendix 1, S9–S10. Defines AI-minus-comparator mean differences, percentage-point differences, transformed relative differences and the noninferiority rule. |
| 14 Synthesis methods | Describe models, assumptions and additional analyses. | Partially reported. Methods, Statistical Analysis; Appendix 1, S9–S12; Multimedia Appendix 3 (R code). Describes regression, covariates, cluster-robust variance estimation, fixed effects, available-case analyses and sensitivity analyses; S12 identifies modifications. Formal residual/model-fit diagnostics and their assessment are not reported in the manuscript or supplement. |
| 15 Study selection results | Report flow, exclusions and reviewer agreement. | Partially reported. Results, Study Sample; Figure 1; Appendix 1, S3. Reports screening and inclusion flow and exclusions in the flow diagram. Calibration targets and adjudication are described, but no observed numerical inter-reviewer agreement statistic is reported. |
| 16 Study characteristics | Provide study characteristics and citations. | Table 1; Tables S1–S2; internal Appendix 3, References for Included Systematic Reviews. Provides aggregate and individual-review characteristics and citations. |
| 17 Risk of bias results | Provide individual appraisal findings. | Results, Risk of Bias; Table S3; Figure S7–S8; Table S12; Multimedia Appendix 2 (dataset). Table S3 provides individual-review overall ROBIS judgments; Figure S8 and Table S12 summarize domain-level comparisons. The overview describes domain- and item-level appraisal scores in the separate dataset. |
| 18 Individual study data | Provide data underlying study-level analyses. | Tables S2–S3; Multimedia Appendix 2 (dataset). The tables provide individual-review characteristics and overall AMSTAR-2, PRISMA, ROBIS and AITDI outcomes, including adapted AMSTAR-2 confidence categories. The overview identifies the separate dataset as the source of complete domain- and item-level appraisal scores. |
| 19 Synthesis results | Report estimates, precision and relevant diagnostics. | Partially reported. Results; Table 2; Figure 2; Tables S4–S16 and S23. Reports estimates, confidence intervals, outcome-specific sample sizes and model complexity. Formal residual/model-fit diagnostics are not reported; between-trial treatment-effect heterogeneity is not the estimand. |
| 20 Additional analysis results | Report additional analyses. | Results, Sensitivity and Exploratory Analyses; Table 2 (matched-set fixed effects); Tables S17–S22; Results, Transparency of AI Use and Table S15. Reports paired, restricted, fixed-effects, journal-exclusion, overlap-weighted, stratified and AITDI association analyses. |
| 21 Summary of evidence | Interpret findings against existing evidence and uncertainty. | Discussion, paragraphs beginning “Reviews with reported AI assistance,” “These findings reflect,” “This study shifts,” and “The discordance,” together with the limitations paragraphs. Interprets findings against prior task-level evidence, distinguishes appraisal constructs, and qualifies causal interpretation and uncertainty. No formal GRADE assessment is reported. |
| 22 Limitations | Discuss methodological biases and limitations. | Partially reported. Discussion, paragraphs beginning “This study has limitations” and “AMSTAR-2 was not designed”; Appendix 1, S5 and S12. Discusses disclosure misclassification, residual confounding, judgment, instrument applicability, incomplete coding guidance, limited precision, generalizability and matching deviations. Publication bias is not separately assessed or explicitly discussed. |
| 23 Conclusions | Explain implications for research and practice. | Discussion, paragraphs beginning “The practical implications,” “Future work should,” and “In conclusion.” Explains implications for disclosure, human oversight, reproducibility, future validation and users of systematic reviews. |
| 24 Funding | Identify support and funder involvement. | Funding section, after Data availability statement and before Contributors. Reports no specific study funding, identifies author support and states that these funding sources had no role in the study or publication decisions. |

Based on Murad MH, Wang Z. Guidelines for reporting meta-epidemiological methodology research. Evid Based Med. 2017; 22:139–142. doi:10.1136/ebmed-2017-110713, Table 1. Requirements are paraphrased; item numbers are added for navigation. Locations refer to manuscript sections, supplementary sections, and tables. Partial reporting is stated explicitly.
